# SARS-CoV-2 infection fatality rates in India: systematic review, meta-analysis and model-based estimation

**DOI:** 10.1101/2021.09.08.21263296

**Authors:** Lauren Zimmermann, Subarna Bhattacharya, Soumik Purkayastha, Ritoban Kundu, Ritwik Bhaduri, Parikshit Ghosh, Bhramar Mukherjee

## Abstract

**Introduction:** Fervorous investigation and dialogue surrounding the true number of SARS-CoV-2 related deaths and implied infection fatality rates in India have been ongoing throughout the pandemic, and especially pronounced during the nation’s devastating second wave. We aim to synthesize the existing literature on the true SARS-CoV-2 excess deaths and infection fatality rates (IFR) in India, through a systematic search followed by viable meta-analysis. We then provide updated epidemiological model-based estimates of the wave 1, wave 2 and combined IFRs using an extension of the Susceptible-Exposed-Infected-Removed (SEIR) model, using data from April 1, 2020 to June 30, 2021.

**Methods:** Following PRISMA guidelines, the databases PubMed, Embase, Global Index Medicus, as well as BioRxiv, MedRxiv, and SSRN for preprints (accessed through iSearch), were searched on July 3, 2021 (with results verified through August 15, 2021). Altogether using a two-step approach, 4,765 initial citations were screened resulting in 37 citations included in the narrative review and 19 studies with 41 datapoints included in the quantitative synthesis. Using a random effects model with DerSimonian-Laird estimation, we meta-analyze IFR_1_ which is defined as the ratio of the total number of observed reported deaths divided by the total number of estimated infections and IFR_2_ (which accounts for death underreporting in the numerator of IFR_1_). For the latter, we provide lower and upper bounds based on the available range of estimates of death undercounting, often arising from an excess death calculation. The primary focus is to estimate pooled nationwide estimates of IFRs with the secondary goal of estimating pooled regional and state-specific estimates for SARS-CoV-2 related IFRs in India. We also try to stratify our empirical results across the first and the second wave. In tandem, we present updated SEIR model estimates of IFRs for waves 1, 2, and combined across the waves with observed case and death count data from April 1, 2020 to June 30, 2021.

**Results:** For India countrywide, underreporting factors (URF) for cases (sourced from serosurveys) range from 14.3-29.1 in the four nationwide serosurveys; URFs for deaths (sourced from excess deaths reports) range from 4.4-11.9 with cumulative excess deaths ranging from 1.79-4.9 million (as of June 2021). Nationwide pooled IFR_1_ and IFR_2_ estimates for India are 0.097% (95% confidence interval [CI]: 0.067 – 0.140) and 0.365% (95% CI: 0.264 – 0.504) to 0.485% (95% CI: 0.344 – 0.685), respectively, again noting that IFR_2_ changes as excess deaths estimates vary. Among the included studies in this meta-analysis, the IFR_1_ generally appear to decrease over time from the earliest study end date to the latest study end date (from 4 June 2020 to 6 July 2021, IFR_1_ changed from 0.199 to 0.055%), whereas a similar trend is not as readily evident for IFR_2_ due to the wide variation in excess death estimates (from 4 June 2020 to 6 July 2021, IFR_2_ ranged from (0.290-1.316) to (0.241-0.651) %).

Nationwide SEIR model-based combined estimates for IFR_1_ and IFR_2_ are 0.101% (95% CI: 0.097 – 0.116) and 0.367% (95% CI: 0.358 – 0.383), respectively, which largely reconcile with the empirical findings and concur with the lower end of the excess death estimates. An advantage of such epidemiological models is the ability to produce daily estimates with updated data with the disadvantages being that these estimates are subject to numerous assumptions, arduousness of validation and not directly using the available excess death data. Whether one uses empirical data or model-based estimation, it is evident that IFR_2_ is at least 3.6 times more than IFR_1._

**Conclusion:** When incorporating case and death underreporting, the meta-analyzed cumulative infection fatality rate in India varies from 0.36%-0.48%, with a case underreporting factor ranging from 25-30 and a death underreporting factor ranging from 4-12. This implies, by June 30, 2021, India may have seen nearly 900 million infections and 1.7-4.9 million deaths when the reported numbers stood at 30.4 million cases and 412 thousand deaths (*covid19india.org*) with an observed case fatality rate (CFR) of 1.35%. We reiterate the need for timely and disaggregated infection and fatality data to examine the burden of the virus by age and other demographics. Large degrees of nationwide and state-specific death undercounting reinforce the call to improve death reporting within India.

## INTRODUCTION

The second wave of SARS-CoV-2 in India–a country broaching 1/5^th^ of the world population, wrought a devastating toll of an astronomical 414 thousand daily reported infections and 4.5 thousand reported daily deaths from COVID-19 (*1*) at its peak in May of 2021, leading to a collapse of healthcare infrastructure (*2*). Concerns have been raised regarding the inadequacy of the healthcare systems of adjacent countries in this region (*3*), as well as tragically realized amidst recent surges of SARS-CoV-2 in some of these countries (*4*).

It is now well-known that only a fraction of SARS-CoV-2 infections are captured, stemming from a large degree of covert infections, access, willingness and availability of testing, and sometimes a desire to maintain public image (*5*) (*6*). Similarly, due to incomplete and lagged reporting, lack of medical certification and misclassification of the cause of deaths, not all COVID-19 related deaths have been captured correctly (*7*) (*8*) (*9*). Infection fatality rates (IFR) are measured as the ratio of the total number of deaths to the total number of infections over a given period. Serological surveys, with rigorous sampling design and cogent analysis, can provide an estimate of the prevalence of antibodies (Ab) formed in response to a past natural infection, and thereby an estimate of the total number of infected cases in an unvaccinated study population, which forms the basis for the denominator of the IFR measure. With introduction of vaccines, seropositivity could arise both from vaccines or from past infections and thus such studies are less informative about the denominator in an IFR calculation. Excess deaths calculation using the death data released during the pandemic period can give us an idea on the number of COVID-19 deaths we failed to report or the extent of death underreporting (*7*) (*9*) (*10*). Compartmental epidemiological models, including extended versions of the susceptible-exposed-infected-removed (SEIR) model and renewal process models can also estimate the total number of latent infections and deaths by making stringent assumptions needed to identify certain key parameters (*11*) (*12*) (*13*).

From a global lens, several systematic reviews have examined this integral measure of SARS-CoV-2 mortality, the IFR, at various points of the pandemic. As of 16 June 2020, a systematic review and meta-analysis of 24 studies estimated a pooled **worldwide** IFR of 0.68% (95% CI: 0.53 – 0.82), while noting considerable heterogeneity among the included studies and being unable to adjust for age, comorbidities, and other demographic factors (limited by the availability of data) (*14*). Other efforts to systematically investigate the infection fatality rates of SARS-CoV-2 include a review and meta-analysis of age-specific IFRs from 27 studies among OECD countries (as of 18 September 2020), which concluded that the IFR increases substantially with age estimating an age-specific IFR of 0.002% among children aged <10, 0.01% among younger adults aged ≥25, 0.4% among adults aged ≥55, 4.6% among adults aged ≥65, and up to 15% among adults aged ≥85 (*15*). Limited to non-developing countries, this review estimated that 90% of the variation in IFR estimates is explained by the age make-up of the underlying populations (*15*). Regarding reconciling SARS-CoV-2 seroprevalence, a key ingredient to know IFR estimates, a systematic review and meta-analysis (as of 14 August 2020) of 47 studies (encompassing 23 countries worldwide) found a worldwide seroprevalence estimate of 3.38% (95% CI: 3.05-3.72) with a range of 0.37% (Malaysia) to 22.1% (Iran) among the included studies, and reported considerable overall heterogeneity among the included studies as well as regional variation in the effect size with 1.45% (95% CI: 0.95-1.84) for South America to 5.27% (95% CI: 3.97-6.57%) for Northern Europe (*16*). India was not examined in these multi-country systematic reviews of SARS-CoV-2 IFR and seroprevalence, and nor were any of its neighboring countries.

Concerning India and its neighboring countries (i.e., Bangladesh, Nepal, Pakistan, and Sri Lanka), certain features common in this region are distinct from the countries considered in the above reviews and thereby beckon a comprehensive investigation of the SARS-CoV-2 fatality within these countries. First, the age structures of these countries are relatively young in comparison to higher income countries (as of 2020, the estimated median age in years are the following: India 28.4, Bangladesh 27.6, Nepal 24.6(*20*), Pakistan 22.8, Sri Lanka 34.0 versus the United Kingdom 40.5, Italy 47.3, Germany 45.7, and the United States 38.3 (*17*)). Considering this difference in the population age composition, a meta-analysis of SARS-CoV-2 infection fatality rates and seroprevalence would aid in discerning the values of these measures specific to South-East Asia. We note that, at the time of this report, no systematic review has examined the existent studies on IFRs within the region encompassing and surrounding India.

Here, we perform a systematic review of existent literature on SARS-CoV-2 infection fatality rates (IFRs) in India and India’s neighboring countries of Bangladesh, Nepal, Pakistan, and Sri Lanka, as of 3 July 2021, with verification of search thoroughness extending to 15 August 2021. Due to the paucity of data and availability of relevant studies in the adjacent countries and considering the intensity of the latest SARS-CoV-2 outbreak in India as detailed above, we focused on India for the quantitative synthesis. Through a meta-analysis of SARS-CoV-2 IFR estimates, we examine the nationwide, regional, and state-specific estimates and heterogeneity over the course of the pandemic within India. As an alternative approach to empirical evidence synthesis and meta-analysis, we use an extended susceptible-exposed-infected-removed (SEIR) model to present updated estimates of SARS-CoV-2 infection fatality rates with data through June 30, 2021, as well as underreporting factors for cases and deaths, corresponding to the first and second waves, as well as cumulative, for COVID-19 in India. This paper concludes with a discussion of takeaways from the systematic review and meta-analysis, as well as from the updated model-based results. We highlight some immediate and future considerations regarding revising the death reporting system in India and why capturing deaths are important for the health and future of the living.

## METHODS

### Terminology and Definitions

Below we provide the definitions, with formulas as needed, of the various terms used throughout this systematic review and meta-analysis.

***Seroprevalence survey (i.e., serosurvey)*** in the context of SARS-CoV-2 are large-scale studies aimed at estimating the true prevalence of SARS-CoV-2 (i.e., the cumulative percent infected over a period of time) within the target population utilizing serology testing for IgG or IgM antibody presence (*18*) scaled to the level of a geographic location (e.g., nationwide, statewide, citywide, or districtwide), a community, or a smaller group. Study designs of serosurveys commonly involve population-based sampling methods (e.g., multi-stage cluster sampling, probability proportionate to size, etc.) applied to obtain a randomly selected, representative sample.

***Case Underreporting Factor (URF (C))*** is defined as the estimated total cumulative infections divided by the reported (i.e., observed) cumulative cases at the indicated date.

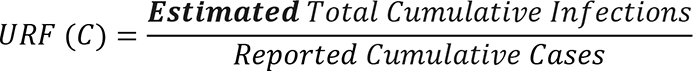

The numerator (estimated total cumulative infections) is estimated by the population of the target geographic region from which samples were selected, multiplied by the seroprevalence, as sourced from serosurveys. The population estimate is age-adjusted according to the age cut-off for the underlying study by multiplying the total population estimate of the study area by the proportion of the population in the age range included in the study sample.

***Excess Deaths*** is defined as the absolute difference between the observed all-cause mortality in a specific time period and the expected all-cause mortality in the same period. When calculating excess deaths, the expected all-cause mortality is commonly projected using prior years of data, an average of prior years, or an alternative benchmark, as it largely depends on the availability and continuity of all-cause mortality data within the location of study.

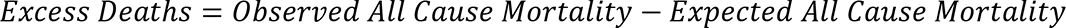

***Death Underreporting Factor (URF (D))*** is defined as the estimated total cumulative deaths divided by the reported (i.e., observed) cumulative deaths at the indicated date, as collected 14 days after the date indicated.

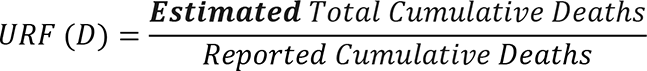

The numerator of the URF (D) (i.e., estimated total cumulative deaths) is the location and time period-specific excess deaths estimate (available at the time of this report), as sourced from media reports and studies on excess deaths. Using all-cause-mortality to calculate excess deaths over a long period has the limitation that the numerator often is not specific to COVID-19 disease. Some model-based studies have alternative ways of estimating this fraction using assumed values of COVID-19 fatality rates.

***Case Fatality Rate (CFR)*** is defined as the reported (i.e., observed) cumulative deaths divided by the reported/observed cumulative cases. Reported cumulative deaths are routinely collected 14 days after the date indicated to allow for fatality delay between symptom onset and death. Whereas reported cumulative cases are collected on the date indicated, as a conservative approach to capturing the cumulative infections up to the date indicated. Further details on the intuition behind the assumption of a 14-day fatality lag can be found in Supplementary Appendix F.

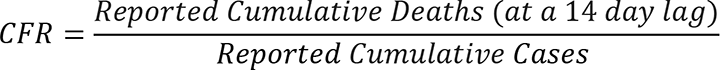

***Infection Fatality Rate_1_ (IFR_1_)*** is defined as the reported (i.e., observed) cumulative deaths (collected 14 days after the indicated date) divided by the estimated total cumulative infections. In other words, IFR_1_ is an estimate of the true fatality rate for which the uncertainty in the denominator is adjusted but not the uncertainty in the numerator, namely, death undercounting is not accounted for.

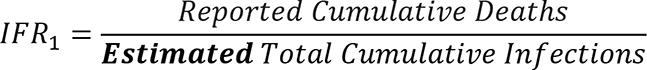

In our meta-analysis, for studies without precalculated IFR_1_, the denominator (estimated total cumulative infections) is computed as the seroprevalence multiplied by the population estimate of the study location. Again, the population estimate is age-adjusted, according to the age inclusion criterion of the study design, by multiplying the population number by the percent of the population in the specified age range.

***Infection Fatality Rate_2_ (IFR_2_)*** is defined as the estimated total cumulative deaths (14 days after the indicated date) divided by the estimated total cumulative infections. In other words, IFR_2_ is an estimate of the true fatality rate for which both the uncertainty in the denominator and the uncertainty in the numerator (i.e., death undercounting) are adjusted.

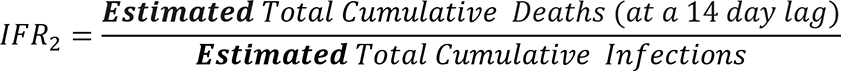

For studies without precalculated IFR_2_, the denominator (estimated total cumulative infections) is the same as the denominator in IFR_1_. The numerator (estimated total cumulative infections) is computed as the reported (i.e., observed) cumulative deaths (collected 14 days after the indicated date) multiplied by the death underreporting factor, as either model-derived or sourced directly from media reports and studies on excess deaths.

### Identification of Studies for Systematic Review and Meta-analysis

The present systematic review and meta-analysis was conducted following the Preferred Reporting Items for Systematic Reviews and Meta-analyses (PRISMA) statement and guidelines (*19*) (see Supplementary Appendix A for the completed PRISMA checklist). Two reviewers developed the search strategy with the expertise and guidance of information specialists from the University of Michigan Taubman Health Sciences Library. On July 3, 2020, we performed the search for relevant articles publicly disseminated from January 1, 2020 onwards and conducted in any language within the databases PubMed, Embase, World Health Organization’s database Global Index Medicus, as well as BioRxiv, MedRxiv, and SSRN through isearch, an interface that houses SARS-CoV-2 related reports curated by the National Institute of Health (a complete search strategy, including search terms, Boolean operators, and applied filters, for each database is published in Supplementary Appendix B). Up until August 15, we verified that no relevant studies were missed by manually combing through studies referenced in media reports, government press releases, and any additional preprints or reports published after July 3 and through August 15, 2021.

For the narrative review, eligible studies included a nationwide, regional (e.g., state, union territory, city, district, or slum designated area, etc.), or age/sex-specific measure of seroprevalence or infection fatality rate (IFR) for India or adjacent countries of Bangladesh, Nepal, Pakistan, and Sri Lanka. Consistent with a systematic review that examines international infection fatality rates of SARS-CoV-2 among OECD countries (i.e. non-developing countries) (*15*), studies were excluded in the full-text screening (a) if the studies examined narrower subpopulations of the location of interest (e.g., healthcare workers, HIV positive adults, or industrial workers, etc.), (b) if the studies recruited participants from certain facilities (e.g., blood donors, dialysis centers), (c) if the studies considered individuals at hospitals, urgent care centers, intensive care units (ICUs), or tertiary care centers, (d) if the studies performed active recruitment of subjects (i.e. participants volunteered), and (e) if the studies limited their cohort to asymptomatic or symptomatic individuals. These criteria are used to identify studies both with representative samples of the general population of interest and with measurements of the outcomes that reflect seropositivity of SARS-CoV-2 regardless of virus manifestation (or irrespective of clinical presentation and presence of symptoms).

For the meta-analysis of infection fatality rates (IFRs), studies were included if (in addition to meeting the eligibility criteria for inclusion to the qualitative review) (a) 95% confidence interval was available for either the seroprevalence and/or the precalculated IFR, when available, and (b) data on COVID-19 confirmed fatalities was available for the study location from *covid19india.org* (*20*) at the time of this review. Additionally, for studies that do not provide a precalculated IFR_2_ (i.e., do not provide an IFR estimate adjusted for death underreporting), a study was excluded from IFR_2_ examination if no excess deaths or death underreporting factors were available (at the time of this review) for the location of interest (e.g., district or city) in media reports or excess deaths focused publications. Namely, this report utilizes excess deaths from three sources for the nationwide meta-analysis of IFR_2_ (*21*) (*22*) (*23*) and the following sources for the regional and state-level IFR_2_ analysis: for cities and districts Ahmedabad (*22*), Bangalore Rural District (*22*), Chennai (*22*) (*24*), Indore (*22*), Mumbai (*25*) (*22*) (*26*), Ujjain (*22*), and for states Delhi (*27*) (*13*) (*22*), Karnataka (*28*) (*29*), Tamil Nadu (*22*) (*28*). Table A below summarizes the sources used in nationwide excess deaths studies (*21*) (*22*) (*30*) (*23*), as well as in the Data Development Lab (*31*), an interactive open source data platform recently made available for India.

**Table A.** Summary of data sources for nationwide excess deaths for India, as of June 2021, used in key

| Ref | Sources |
| --- | --- |
| Leffler et al., 2021 (21) | 1. state-specific excess deaths estimates (see **Table 3**) |
| Guilmoto, 2021 (32) | 1. Age and sex-specific deaths from Kerala sample (https://dashboard.kerala.gov.in/deaths.php)<br>2. Deaths of elected officials from Member of the Legislative Assembly sample (https://tcpd.ashoka.edu.in/lok-dhaba/)<br>3. Deaths of railway workers from the Indian Railways sample (https://indianrailways.gov.in/railwayboard/uploads/directorate/stat_econ/Annual-Reports-2019-2020/Year-Book-2019-20-English_Final_Web.pdf)<br>4. Deaths of schoolteachers from Karnataka sample (*Not available*) |
| Deshmukh et al., 2021 (22) | 1. Civil Registration System (https://crsorgi.gov.in/about-us.html),<br>2. MoHFW facility-based reported deaths,<br>3. Cvoter India OmniBus telephone survey (https://teamcvoter.com/) |
| Ramachandran et al., 2021 (30) | 1. Consumer Pyramids Household Survey (https://consumerpyramidsdx.cmie.com/) |
| Anand et al., 2021 (23) | 1. Consumer Pyramids Household Survey (https://consumerpyramidsdx.cmie.com/),<br>2. Civil Registration System (https://crsorgi.gov.in/about-us.html) |
| Data<br>Development Lab<br>(devdatalab.org/covid)<br>(31) | 1. state-specific excess deaths estimates (see the mortality data documentation on the Data Development Lab website, devdatalab.org/covid) |

### Two-Step Screening and Data Abstraction

Citation titles and abstracts were screened independently by two reviewers for verifying the inclusion and exclusion criteria. For articles not satisfying the exclusion criterion, the full text was accessed, and the two screeners separately reviewed the methods and results sections of these studies for final inclusion into this systematic review and meta-analysis. The two reviewers subsequently extracted data from the included articles on the following items: (a) study design, location, and time period, as well as inclusion criteria, (b) sample size denoting the number of participants in the study, (c) descriptive statistics of the proportion of the study cohort male versus female and the mean age of the study cohort, (d) estimate of infection fatality ratio (IFR) or seroprevalence for SARS-CoV-2 and corresponding 95% confidence intervals, when available, (e) any statistical sensitivity tests performed.

### Statistical Meta-analysis

A detailed description of the meta-analysis framework is presented in Supplementary Appendix F. To perform the meta-analyses presented herein, the R package *meta* (*33*) was utilized and the R package *metafor* (*34*) was employed to perform the included sensitivity analyses.

In preparation for the meta-analysis, we extract the IFR or seroprevalence estimates, as well as the provided 95% CI, from each study corresponding to the study’s intended overall population. For seroprevalence studies without precalculated IFR_1_, we collect data on **(a)** COVID-19 confirmed fatalities from *covid19india.org* (*20*) 14 days after the end date of the study period (as previously reasoned), **(b)** 2019 projected population estimates from the World Bank for India (*35*), 2019 projected population estimates from the 2011 census for states (*36*), and 2011 census population estimates for districts and cities (limited by available projections) (*36*), **(c)** 2011 census information on age demographics for the study location (*37*), and **(d)** underreporting factors, as derived or directly reported from media reports and studies on excess deaths for the nation, state, district, or city were collected nearest to the end date of when the study was conducted. With these collected data items, for studies without a precalculated fatality rate, we compute IFR_1_ and IFR_2_ following the definitions provided in the ***Terminology and Definitions*** section above. We note that for smaller regional denominations (e.g., cities or districts) for which data on excess deaths were unavailable, we use the statewide death underreporting factor (if available) in computing IFR_2_.

For the meta-analysis itself, we estimate three versions of SARS-CoV-2 pooled IFRs– namely (a) IFR_1_, (b) IFR_2_ with lower limit of the reported estimates of URF (D), and (c) IFR_2_ with higher limit of the reported estimates of URF (D)– for each sub-analysis. First, using a random effects model with inverse variance method and DerSimonian-Laird (DL) estimation, we estimate the nationwide SARS-CoV-2 pooled IFRs, as well as 95% confidence intervals, for India across all available national IFR datapoints (both precalculated and computed herein) included in this review. Next, a regional analysis of IFRs is performed considering India partitioned into the following regions: North, Northeast, East, South, West, and Central. Regions with at least one available eligible study are examined. To compute the regional SARS-CoV-2 pooled IFRs, we first pool the IFR estimates from statewide, citywide, and districtwide studies associated with each state, using the same random effects estimation approach. Then, the regional SARS-CoV-2 pooled IFRs are approximated with the statewide pooled IFR estimates, using the analogous random effects model with the inverse variance approach and DL estimator.

In both the nationwide and regional analyses, **heterogeneity** is assessed using the *I^2^* statistic, where a *I^2^* of 50% to 90% denotes substantial heterogeneity and *I^2^* ≥ 75% (and up to 100%) indicates considerable heterogeneity among the included studies, as recommended by Cochrane guidelines (*38*). For the regional analysis, we interpret the *I^2^* with caution as we do expect there to be variation in the true effect size among the included studies, as these studies cover varying states and IFRs have been found to vary by state (*13*).

To assess **publication bias,** we produce a funnel plot to examine the distribution of p-values and 95% confidence intervals, as well as perform the Egger’s and Begg’s tests to formally test for funnel plot asymmetry. To assess the **risk of bias** in the seroprevalence estimates across the included studies, we utilized the Joanna Briggs Institute (JBI) approach (*39*) (*40*). Upon completing the series of questions for each study, the tool provides a rating of risk of bias (low/ moderate/ high) based on the sampling design and data collection methods. We answer each question according to the provided values (“Yes”/ “No”/ “Unclear”/ “Not applicable”). Remarks are included with respect to the IFR estimates within the returned strata of the risk of bias.

### Model-based Estimation of Fatality Rates in India during Wave 1, Wave 2, and combined over waves from April 1, 2020 to June 30, 2021

The present study follows the same methodology as the previous versions (*13*). In brief, we employ a compartmental epidemiologic model, the extended susceptible-exposed-infected-removed (SEIR-*fansy*), to model the transmission dynamics of SARS-CoV-2. Accounting for the false negative rates of the SARS-CoV-2 antigen test type (e.g., RT-PCR and rapid antigen tests) observed in India, we include unascertained cases and deaths as a compartment. As depicted in Supplementary Appendix K Figure S1, the population is further divided into 10 disjoint compartments: S (Susceptible), E (Exposed), T (Tested), U (Untested), P (Tested Positive), F (Tested False Negative), RR (Reported Recovered), RU (Unreported Recovered), DR (Reported Deaths) and DU (Unreported Deaths). A full description of the methodological framework is available in Supplementary Appendix K. Nine differential equations form the basis for imitating the spread of SARS-CoV-2. Parameters and 95% credible intervals (CrI) are estimated using Bayesian techniques by performing Metropolis–Hastings algorithm-based random sampling from the posterior distribution with a Gaussian proposal density.

Considering the inherent differences between the first and second waves of the pandemic (e.g., the strain on the healthcare infrastructure and dearth of medical resources in wave 2), we examine the entailed epidemiological measures for wave 1 and wave 2 separately, with wave 1 as April 1 2020–January 31, 2021 and wave 2 as February 1, 2021–June 30, 2021 (latest until this report). The start date of wave 2 of February 1, 2021 is reasoned as two weeks before the national effective reproduction number crossed unity on February 14, 2021 to allow for time between virus exposure and symptom onset. Daily infected cases, recovered cases, and deaths are collected from *covid19india.org* (*20*). We compute nationwide and state-wise fatality rates and case–as well as death– underreporting factors for both wave 1 and wave 2. Additionally, we estimate cumulative infected case and death counts across waves 1 and 2 adjusting for the respective underreporting factors. To obtain the combined number of cases across both waves, we multiply the wave-specific infections with the underreporting estimate and sum across the two waves. Similarly, to obtain the combined number of deaths across both waves, we compute the combined number of deaths by multiplying the wave-specific deaths with the underreporting estimate and sum across the two waves. Lastly, an overall infection fatality rate (IFR) for India as of 30 June 2021 is provided as the estimated cumulative number of deaths divided by the estimated cumulative number of infections.

## RESULTS

### Search Results and Included Studies

As shown in Figure 1 below, the search resulted in a total of 5174 citations across the four search engines: PubMed (2940), Embase (1119), WHO Global Index Medicus (Southeast Asia Region: 5, Eastern Mediterranean Region: 2), and isearch (1078). After removing duplicate citations across databases, 4765 articles were screened in the initial level of title/abstract, and we reviewed 137 articles in the second level of full-text screening. Of these studies, 61 studies were excluded as no seroprevalence, nor infection fatality rate estimates were presented. Moreover, 18 studies were excluded as the cohort consisted of healthcare workers, 15 studies were excluded as the cohort consisted of patients from hospitals, urgent care units, or tertiary care centers, 5 studies were excluded because the cohorts were limited to either asymptomatic (4 studies) or symptomatic (1 study) individuals, 4 studies were excluded as the cohort consisted of working individuals, 2 studies were excluded as the cohort consisted of blood donors, 2 studies were excluded as the cohort was sampled from testing centers, 1 study was excluded because the cohort consisted of members from elderly care centers, 1 study was excluded as the participants were actively recruited. Overall, across these subpopulations, 48 studies were collectively excluded for the reason of having narrow or non-representative samples, which in turn impedes generalizability to the broader target population within the location of study.

**Figure 1.**
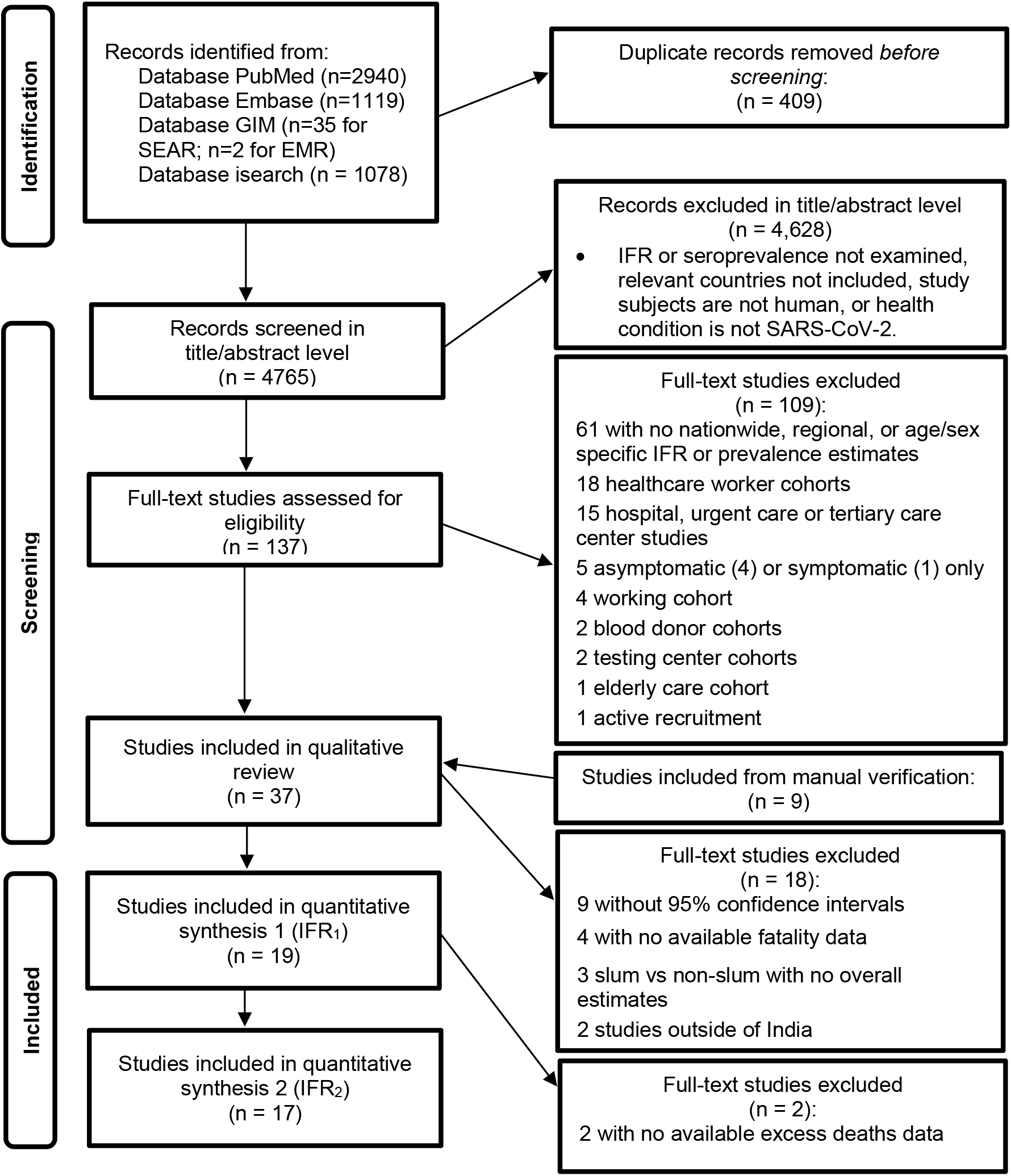
PRISMA flow diagram (41)

### Qualitative Review

#### Locations of included articles

Of the 37 articles included in the narrative review, 2/5 countries originally in the search (namely, India and Pakistan) were present in these articles. In other words, no eligible studies were available for Bangladesh, Nepal, nor Sri Lanka. The predominant country studied was India (35/37 included articles) with Pakistan being the other country (2/37 included articles). In the initial screening, nearly all citations were written in English with 1 citation written in French. In the full-text screening, all articles were written in English.

#### Study designs, populations, and objectives

As shown in Supplementary Appendix C Table 1, among the 37 included articles, 27/37 were serosurvey studies (25 serosurveys for India (*42*) (*43*) (*44*) (*45*) (*46*) (*47*) (*48*) (*49*) (*50*) (*51*) (*52*) (*53*) (*54*) (*55*) (*56*) (*57*) (*58*) (*59*) (*60*) (*61*) (*62*) (*63*) (*64*) (*65*) (*66*) and 2 serosurveys for Pakistan (*67*) (*68*)) and 10/37 were other study designs (*69*) (*70*) (*71*) (*72*) (*73*) (*74*) (*13*) (*12*) (*75*) (*76*), such as extended compartmental epidemiological model-based approaches.

The age distributions of the study populations reflect the younger populations in India and Pakistan with 6/11 studies (that reported the study age composition) having a mean age of ≤24 to <40 years and 5/11 studies with a mean age of ≤40 to <50 years (Supplementary Appendix C Table 1). Regarding the participation rates by gender, out of the 19 studies (that reported the study gender composition) 5 studies had a considerably larger proportion of male than female participants, whereas 3 studies had a heavier percentage of female than male participants (Supplementary Appendix C Table 1). The remaining 11/19 had a comparable percentage of male versus female participants.

A nationwide target population accounted for 10/37 studies, namely, 8 for India (*59*) (*60*) (*61*) (*62*) (*74*) (*13*) (*12*) (*75*) (*76*) and 1 for Pakistan (*68*). The predominant population of interest was cities with 19/37 studies focusing on municipal target populations and with the following counts of studies per city: 6 Delhi–India (*47*) (*48*) (*49*) (*50*) (*70*) (*71*), 4 Mumbai–India (*56*) (*57*) (*58*) (*73*), 1 Ahmedabad–India (*42*), 2 Bhubaneswar–India (*44*) (*45*), 2 Chennai–India (*46*) (*69*), 1 Indore–India (*55*), 1 Pimpri-Chinchwad–India (*63*), 1 Ujjain–India (*66*), and 1 Karachi–Pakistan (*67*). The next highest population of interest was state and union territories with 6/37 studies focusing on statewide generalizations and the following counts of studies per state or union territory: 3 Karnataka–India (*52*) (*53*) (*72*), 1 Kashmir–India (*54*), 1 Puducherry–India (*64*), and 1 Tamil Nadu–India (*65*). The prevalence of SARS-CoV-2 in slum dwellings was considered in 8/37 studies with the following counts of studies per slum area: 3 slum sections within Mumbai–India (*56*) (*57*) (*73*), 1 slum sections within Matunga, Chembur West, and Dahisar wards in Mumbai– India (*58*), 1 Devarajeevana Halli (DJ Halli) slum in Bengaluru–India (*51*), 1 slum sections within Pimpri-Chinchiwad–India (*63*) and 2 nationwide slum prevalence approximations from the 2^nd^ and 3^rd^ nationwide serosurveys for India (*60*) (*61*). Seroprevalence of SARS-CoV-2 within rural districts versus urban was examined in 9/37 included studies and with the following counts per area: 1 Bangalore Rural District (with subdistricts–otherwise referred to as taluks of Devanahalli, Doddaballapura, Hoskote, Nelamangala)–India (*43*), 1 Delhi–India (*50*), 1 Karnataka–India (*52*), 1 Kashmir–India (*54*), 1 Puducherry–India (*64*), 1 Tamil Nadu–India (*65*) and 3 nationwide–India (*59*) (*60*) (*61*).

#### SARS-CoV-2 surveillance measurements and levels of granularity

As presenting either an IFR or seroprevalence estimate was an inclusion criterion, all 37 included studies estimated one or both of these measures. The breakdown of measures examined was as follows: 8/37 studies presented only IFR estimates without estimating seroprevalence, 15/37 studies presented only seroprevalence estimates without computing IFRs, and 14/37 studies reported both estimated seroprevalence and IFRs. Among the included studies, seroprevalence was estimated at the following levels: overall region, subregion (e.g., district/ward), type of residence (e.g., urban/rural/slum/non-slum), age, sex, age-sex, and for select comorbidities (e.g., diabetes and hypertension). In contrast, IFRs were estimated largely at the overall level with just 2 studies presenting age-sex stratified estimates (1 serosurvey for Tamil Nadu (*65*) and 1 other study design for Karnataka, Mumbai and Bihar migrants (*72*)).

#### Remarks on excluded articles

The prevailing reason for exclusion of studies in the full-text screening was that the article did not provide concurrent prevalence estimate nor an IFR (61/109). Of these 61 studies presenting other epidemiological measures, a large number examined CFRs. Additionally, a considerable number of studies (18/109) were excluded that examined the subpopulation of healthcare workers and with the following counts of healthcare worker studies per country: India (16) and Pakistan (2). The third notable exclusion criterion was studies that examined hospital, urgent care, or tertiary care centers (15/109), which resulted in the following counts of patient or hospitalized studies per country: India (11), Pakistan (2), Bangladesh (1), and Nepal (1). A complete list of studies excluded with reasonings is presented in the Supplementary Appendix D, as well as a summary table synthesizing the total number of studies excluded per eligibility criterion by country (see Supplementary Appendix E).

### Quantitative Summary for India: Meta-analysis of Infection Fatality Rates (IFR)

Of the 37 eligible studies in the narrative review, 19 and 17 studies were included in the meta-analysis of IFR_1_ and IFR_2_, respectively. Supplementary Appendix G lists the studies that were not able to be included in the meta-analysis and each respective reason for exclusion. To summarize, the reasons for exclusion include: (a) did not provide a 95% confidence interval for the seroprevalence and/or IFR estimate, (b) fatality data were unavailable for the study location (e.g., city or select districts) from *covid19india.org* (*20*), (b) excess deaths data were unavailable at the state level for the study location in order to derive the death underreporting factor, (d) studies in Pakistan as previously discussed the meta-analysis focuses on India, and (e) studies that stratify by slum versus non-slum and do not provide an overall estimates are excluded, as individuals living in slum areas tend to be of low socioeconomic status relative to persons in non-slum areas.

#### Summary of Nationwide Underreporting of Infections

As shown in Table 1 below, the breadth of case underreporting countrywide has remained high in wave 2 compared to wave 1, as is evidenced by the dip in case underreporting factors (URF) from 29.1 to 14.3 in the 1^st^ to the 2^nd^ nationwide serosurveys compared to the consistency in high case URFs from 25.7 to 24.9 in the 3^rd^ and 4^th^ nationwide serosurveys, respectively, in India (*59*) (*60*) (*61*) (*62*). Please note that the 4^th^ serosurvey did include vaccinated individuals, but by June 30, only roughly 5% of the Indian population was fully vaccinated and under 20% were partially vaccinated, so the estimates reflect largely antibodies from past infections but are likely overestimates.

**Table 1.**
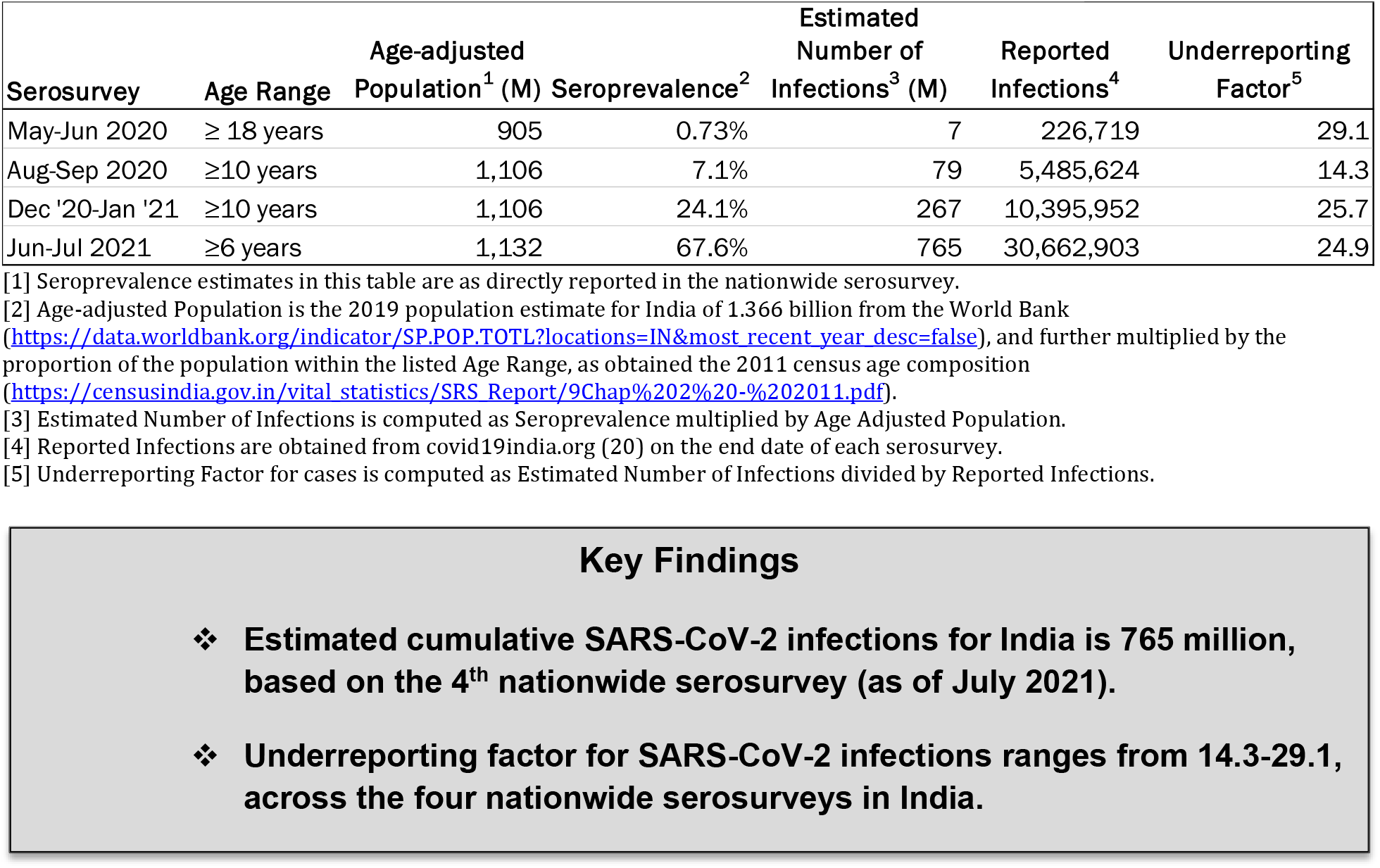
Nationwide seroprevalence and case underreporting factors for India.

| Serosurvey | Age Range | Age-adjusted Population<sup>1</sup> (M) | Seroprevalence<sup>2</sup> | Estimated Number of Infections<sup>3</sup> (M) | Reported Infections<sup>4</sup> | Underreporting Factor<sup>5</sup> |
| --- | --- | --- | --- | --- | --- | --- |
| May-Jun 2020 | ≥ 18 years | 905 | 0.73% | 7 | 226,719 | 29.1 |
| Aug-Sep 2020 | ≥10 years | 1,106 | 7.1% | 79 | 5,485,624 | 14.3 |
| Dec '20-Jan '21 | ≥10 years | 1,106 | 24.1% | 267 | 10,395,952 | 25.7 |
| Jun-Jul 2021 | ≥6 years | 1,132 | 67.6% | 765 | 30,662,903 | 24.9 |

[1] Seroprevalence estimates in this table are as directly reported in the nationwide serosurvey.
[2] Age-adjusted Population is the 2019 population estimate for India of 1.366 billion from the World Bank (https://data.worldbank.org/indicator/SP.POP.TOTL?locations=IN&most_recent_year_desc=false), and further multiplied by the proportion of the population within the listed Age Range, as obtained the 2011 census age composition (https://censusindia.gov.in/vital_statistics/SRS_Report/9Chap%20%20-%202011.pdf).
[3] Estimated Number of Infections is computed as Seroprevalence multiplied by Age Adjusted Population.
[4] Reported Infections are obtained from covid19india.org (20) on the end date of each serosurvey.
[5] Underreporting Factor for cases is computed as Estimated Number of Infections divided by Reported Infections.

#### Background on Nation and State Level Excess Deaths

As shown in Table 2, five recent studies have provided nationwide excess deaths estimates for India, as of June 2021 (*21*) (*32*) (*22*) (*30*) (*23*). Of the three studies that examine excess deaths in India broken out by waves 1 and 2 (*21*) (*22*) (*23*), excess deaths are generally found to be considerably higher in wave 2 than wave 1 with the exception of the Anand et al., 2021 study (*23*), where excess deaths decrease from 3.4 (wave 1) to 1.5 million (wave 2). We note that the time period of the second wave in this study (*23*) extends into wave 2, as the national effective reproductive number crossed unity on February 14, 2021 in India, which may in part explain the high excess deaths estimate comparable to the other two studies. The degree of nationwide death underreporting has remained high from waves 1 to 2 with the Wave 1 URF (D) ranging from 3.8-19.6 and the Wave 2 URF (D) ranging from 4.0-11.2 (Table 2). Up to June 2021, the cumulative excess death estimates across waves 1 and 2 for India range from 1.79 million (*21*) to 4.9 million (*23*), suggesting a combined nationwide URF (D) of 4.4-11.9.

**Table 2.** Nationwide excess deaths and death underreporting factors for India in waves 1, 2, and combined.

| Ref/First Author | Wave 1<br>(Apr 2020-Jan 2021) |  |  |  | Wave 2<br>(Feb-Aug 2021) |  |  |  | Combined across Waves 1 and 2<br>(Apr 2020-Aug 2021) |  |  |  |
| --- | --- | --- | --- | --- | --- | --- | --- | --- | --- | --- | --- | --- |
|  | Time Period | Excess Deaths (M) | COVID-19 Reported Deaths<sup>1</sup> | Under Reporting Factor<sup>2</sup> | Time Period | Excess Deaths (M) | COVID-19 Reported Deaths<sup>1</sup> | Under Reporting Factor<sup>2</sup> | Time Period | Excess Deaths (M) | COVID-19 Reported Deaths<sup>1</sup> | Under Reporting Factor<sup>2</sup> |
| (21) Leffler | Jan-Dec 2020 | Low: 736 th<br>High: 1.04 | 151,954 | Low: 4.8<br>High: 6.9 | Jan-Jun 2021 | Low: 1.06<br>High: 1.58 | 263,001 | Low: 4.0<br>High: 6.0 | Jan '20-Jun '21 | Low: 1.79<br>High: 2.62 | 412,019 | Low: 4.4<br>High: 6.4 |
| (32) Guilmoto | - | - | - | - | - | - | - | - | Mar '20-May '21 | Low: 2.16<br>High: 2.45 | 353,558 | Low: 6.1<br>High: 6.9 |
| (22) Deshmukh | Jun '20-Jan '21 | Low: 595 th<br>High: 658 th | 155,769 | Low: 3.8<br>High: 4.2 | Apr-Jun 2021 | Low: 2.51<br>High: 2.77 | 249,059 | Low: 10.1<br>High: 11.2 | Jun '20-Jun '21 | Low: 3.11<br>High: 3.43 | 412,019 | Low: 7.5<br>High: 8.3 |
| (30)<sup>3</sup> Ramachandran | - | - | - | - | - | - | - | - | Jan '20-Jun '21 | 3.36 | 404,211 | 8.4 |
| (23) Anand | Apr '20-Mar '21 | 3.4 | 173,153 | 19.6 | Apr-Jun 2021 | 1.5 | 249,059 | 6 | Apr '20-Jun '21 | 4.9 | 412,019 | 11.9 |

[1] COVID-19 Reported Deaths are obtained 14 days after the end date of the time period from *covid19india.org* (20), unless otherwise noted.
[2] Underreporting Factor is computed as Excess Deaths divided by COVID-19 Reported Deaths, unless otherwise noted.
[3] Underreporting Factor (URF), as well as COVID-19 Reported Deaths, are directly reported in this study. Hence, the URF in this table is the precalculated estimate provided.

As shown below in Table 3, the statewide excess deaths available at the time of this report largely show a similar story of increased excess deaths in wave 2 relative to wave 1, corroborating the severe toll of the second wave of the pandemic on India. In parallel, the state-specific death URFs are consistently higher in wave 2 compared to wave 1 (Table 3), actualizing the widely discussed absence and deterioration of timely and accurate death reporting in wave 2 in various states in India. The exception to this assertion is Kerala where the wave-specific death URFs are not meaningful different and excess deaths have remained low or albeit negative (Table 3). In wave 1, the two states with the highest death URFs are Assam (14.1-19) and Andhra Pradesh (9.1-11.1), whereas the two states with the lowest death underreporting that in fact exhibited negative excess deaths compared to prior year(s) are Gujarat (−70 thousand deaths) and Uttar Pradesh (−37 thousand deaths). In wave 2, the two states with the largest death underreporting are Madhya Pradesh (23.8-42.4) and again Andhra Pradesh (24.6-36.2). The two states with the lowest death URFs in wave 2, where excess deaths were lower than the number of reported COVID-19 deaths, are Kerala and Delhi. It is important to recognize that the timing of the onset of the second wave differs by state and city, wherein Table 3 the categorization of waves is based on the timing of the second wave nationwide in India.

**Table 3.**
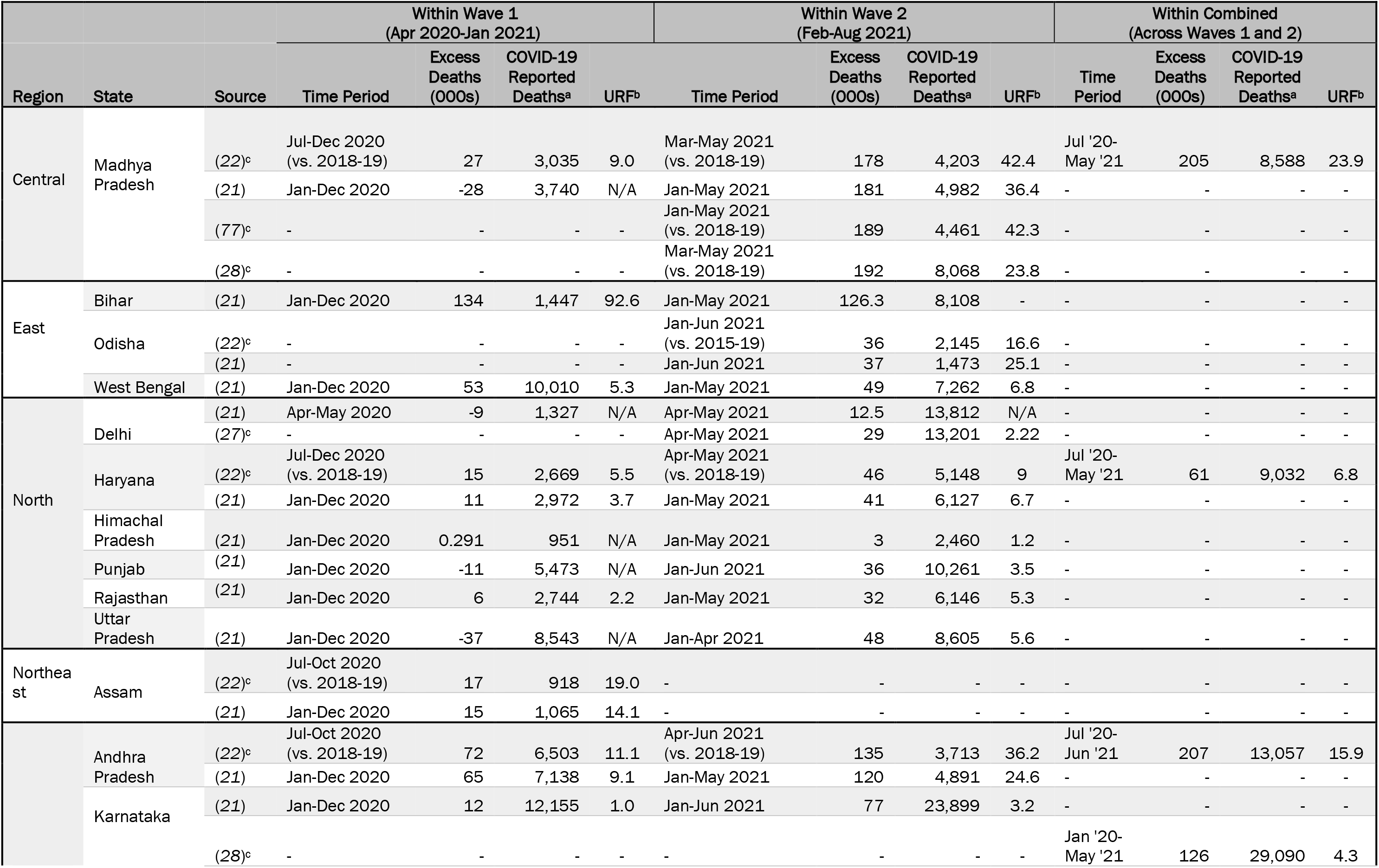
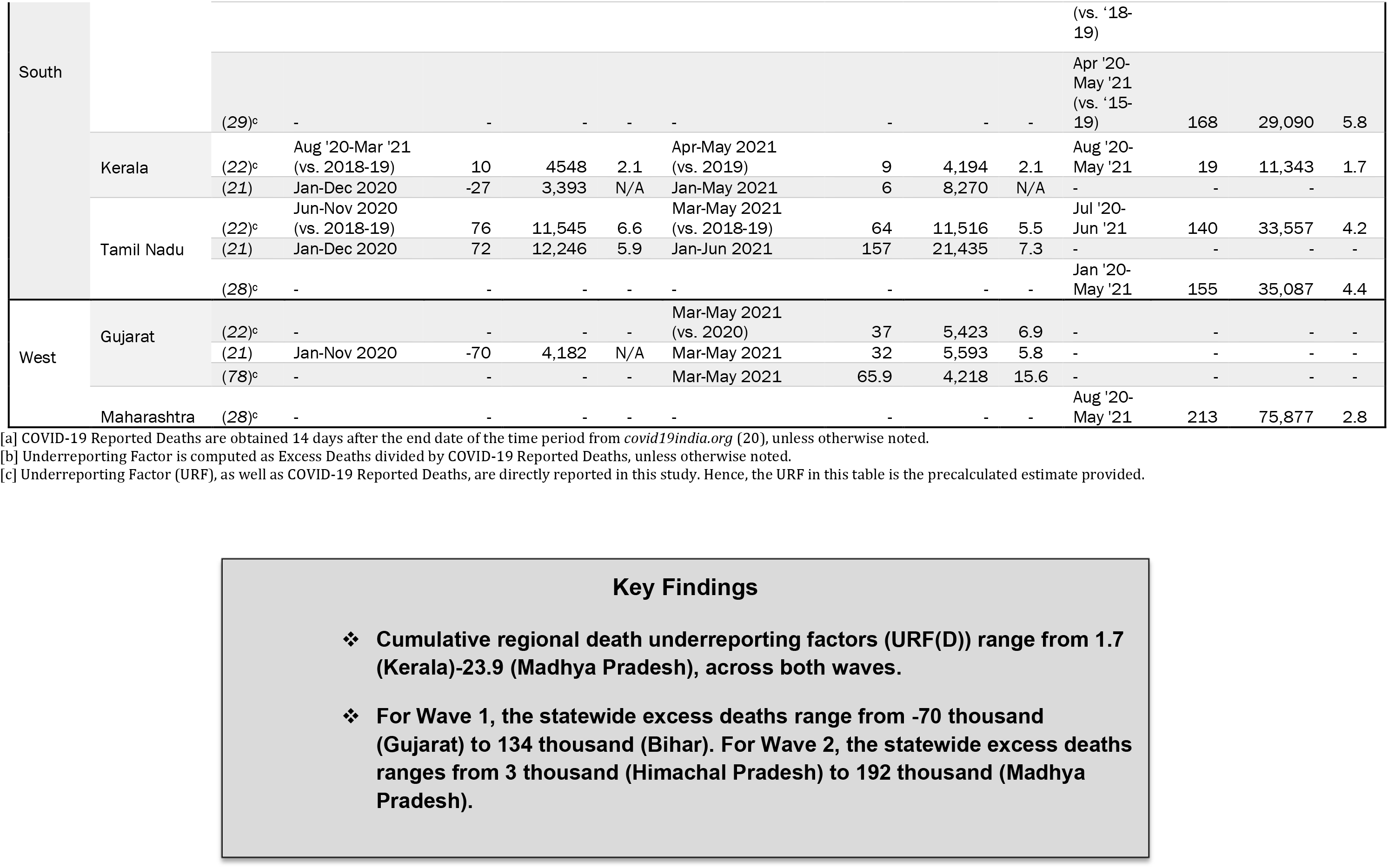
Statewide excess deaths and death underreporting factors for India in waves 1, 2, and combined.

#### Nationwide IFR_1_ and IFR_2_

Figure 2 presents the results of the nationwide meta-analysis of IFR_1_ for India. Using a random effects approach with DL estimation, the estimated nationwide pooled SARS-CoV-2 IFR_1_ for India is 0.097% (95% CI: 0.067 – 0.140). This suggests a nationwide SARS-CoV-2 fatality ratio for India of 97 deaths per 100,000 infections, when not accounting for underreporting of deaths. As seen in Figure 2, there is considerable variation in the time periods encompassed in the studies belying this overall estimate. The range of IFR_1_ (from highest to lowest) among the included studies is 0.199% (4 June 2020) to 0.055% (6 July 2021).

**Figure 2.**
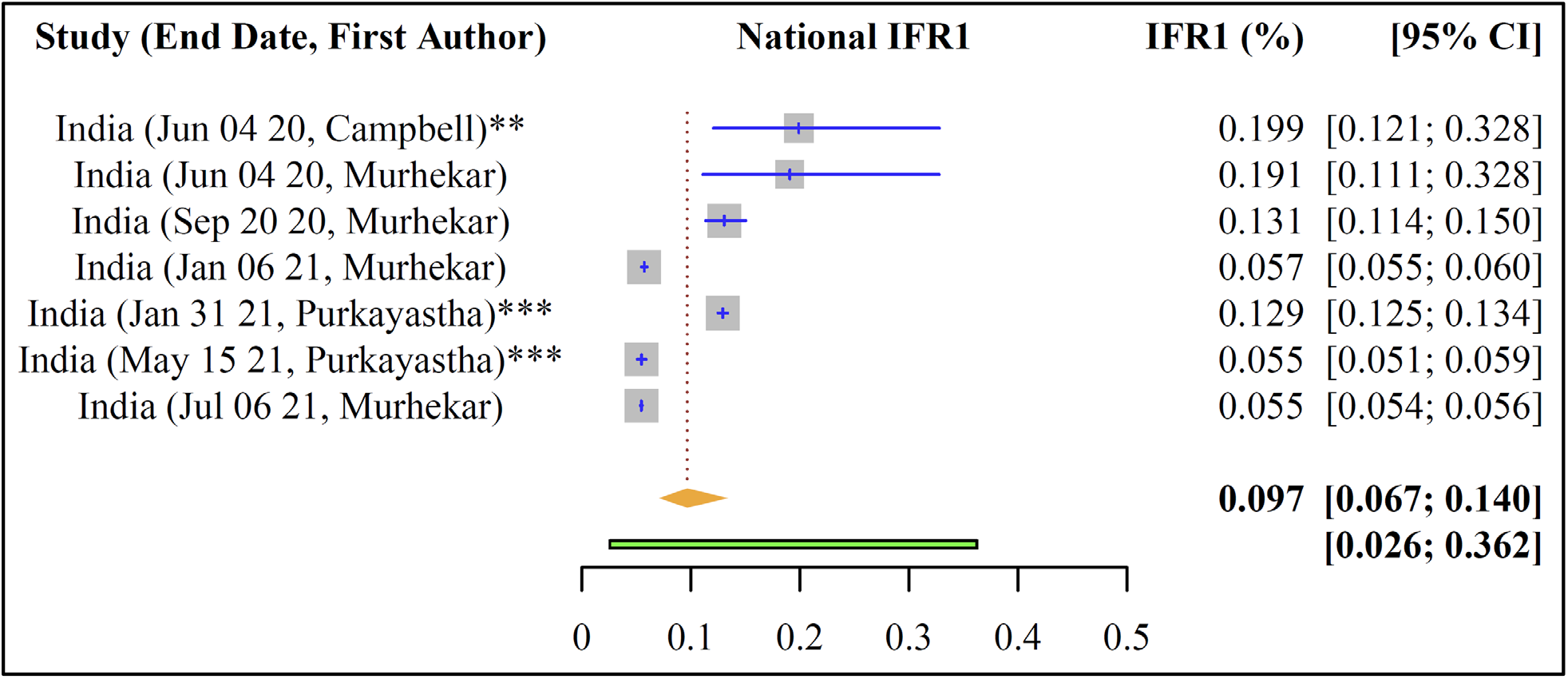
Nationwide estimated pooled IFR_1_ of SARS-CoV-2 for India. Note: * = IFR_1_ precalculated. ** = IFR_2_ precalculated. *** = both IFR_1_ and IFR_2_ precalculated. For those with no stars, IFR_1_ and IFR_2_ as well as 95% CIs were computed.

Figure 3 shows the results of the nationwide meta-analysis of IFR_2_ for India. Using a random effects approach with DL estimation, the estimated nationwide pooled SARS-CoV-2 IFR_2_ for India ranges from 0.365% (95% CI: 0.264 – 0.504) to 0.485% (95% CI: 0.344 – 0.685), when applying the lower and higher ends of the uncertainty bracket for death underreporting, respectively. This can be interpreted as a nationwide SARS-CoV-2 fatality ratio for India of 365-485 deaths per 100,000 infections, when further adjusting for underreporting of deaths.

**Figure 3.**
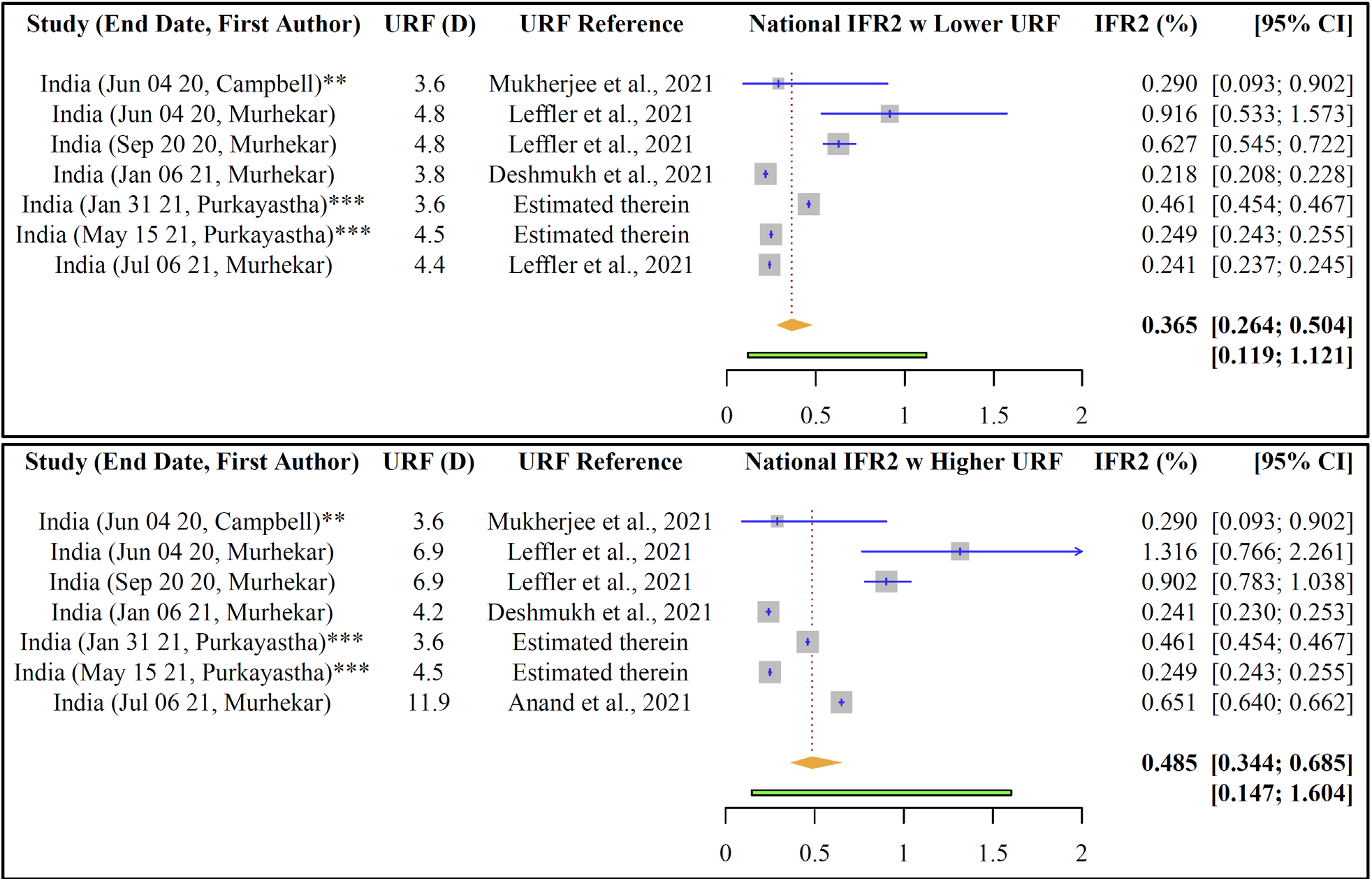
Nationwide estimated pooled IFR_2_ of SARS-CoV-2 for India, with lower and higher URF uncertainty. Note: * = IFR_1_ precalculated. ** = IFR_2_ precalculated. *** = both IFR_1_ and IFR_2_ precalculated. For those with no stars, IFR_1_ and IFR_2_ as well as 95% CIs were computed.

A detailed forest plot of the nationwide seroprevalences of SARS-CoV-2 in India, as well as the accompanying 95% confidence interval, underlying the computed nationwide IFRs, as directly reported in the included studies, can be found in Supplementary Appendix H Figure 1.

With respect to observed time trends from this meta-analysis, Table 4 includes a comparison of the case fatality rate (CFR) to IFR_1_ and IFR_2_ in waves 1 versus combined across waves 1 and 2. Both the CFR (i.e., not accounting for case nor death underreporting) and IFR_1_ (i.e., not accounting for death underreporting) have decreased overtime. Specifically, the nationwide CFR has decreased from 5.56% in June 2020 to 1.36% in July 2021 (Table 4). The nationwide IFR_1_ has also largely been decreasing over time from 0.199% (4 June 2020) to 0.055% (6 July 2021), based on estimates (both precalculated and computed) from included studies (Table 4). Whereas for IFR_2_ (i.e., when adjusting for both case and death undercounting) this trend is not readily apparent. As a whole, IFR_2_ has changed overtime between 0.290% (4 June 2020) to 0.241-0.651% (6 July 2021), oscillating between 0.241% (6 Jan 2021) to 0.916-1.316% (4 June 2020), as detailed in Table 4. The absence of this trend for IFR_2_ reconciles with the large excess deaths estimates observed for wave 2 compared to wave 1 (Table 2) and, in particular, the increase in the nationwide death URFs of 3.8-4.2 (wave 1) to 10.1-11.2 (wave 2) (Table 2), as derived from the excess deaths estimated in Deshmukh et al., 2021 (*22*).

**Table 4.**
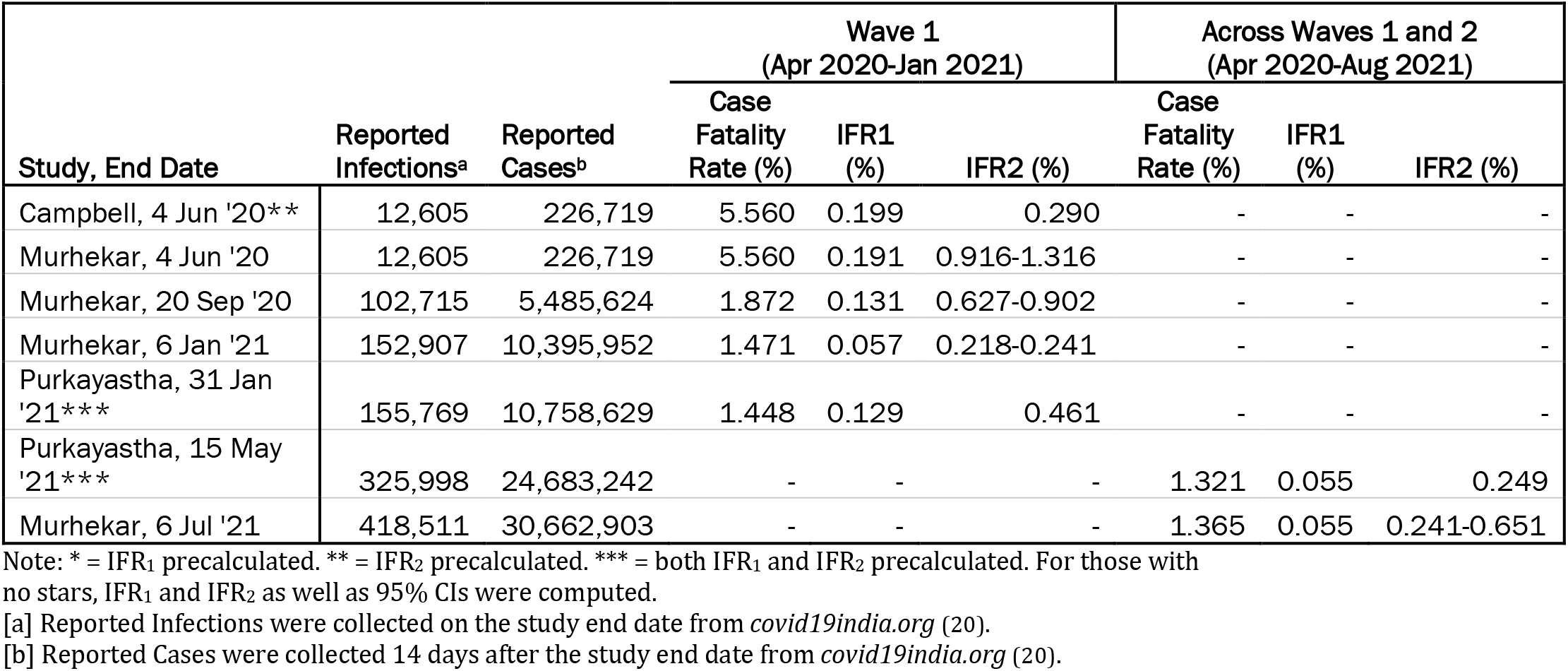
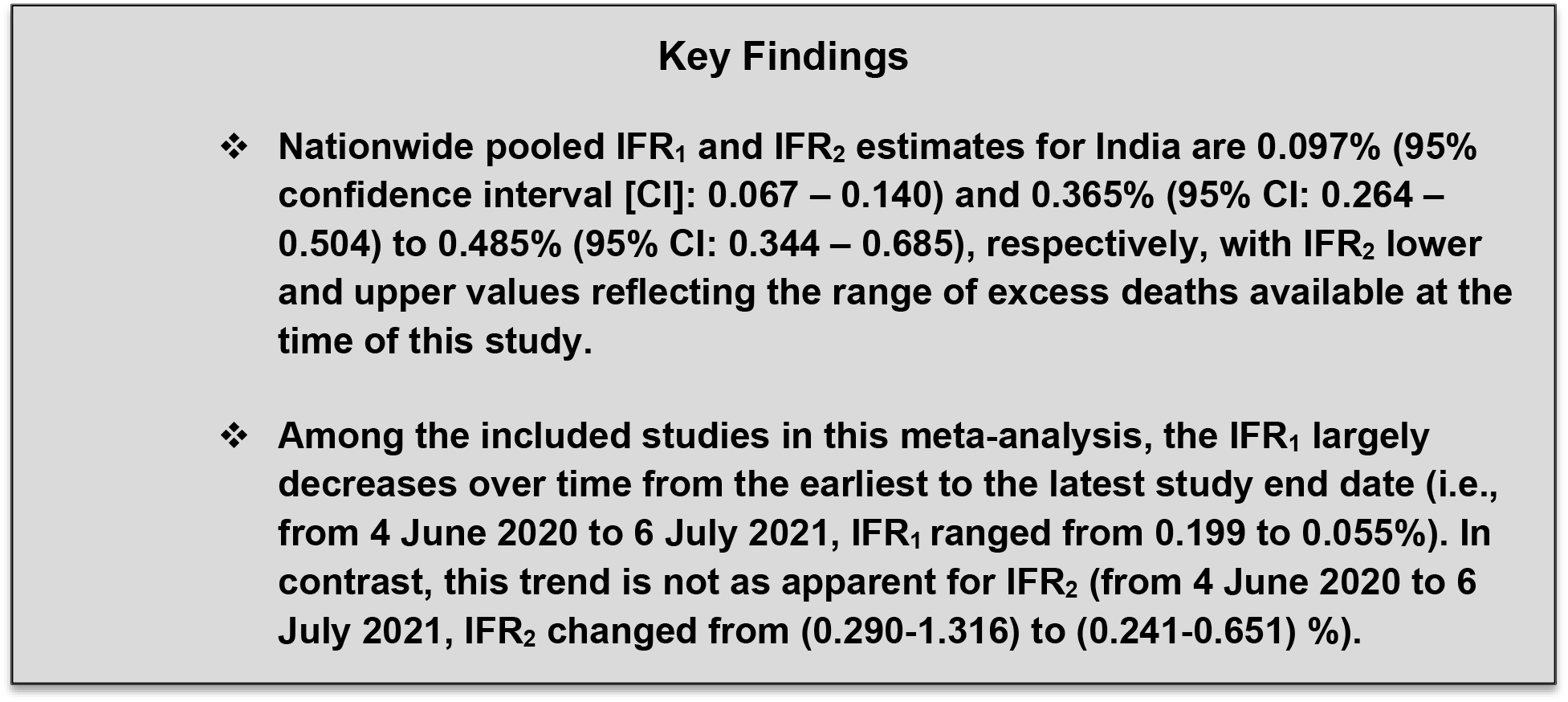
Comparison of nationwide CFR to IFR_1_ and IFR_2_ from included studies for wave 1 and across waves.

##### Regional and Statewide IFR_1_ and IFR_2_

Detailed estimates of regional pooled IFR_1_ and IFR_2_ of SARS-CoV-2 in India and 95% confidence intervals (CI), as well as the statewide pooled IFR_1_ and IFR_2_ estimates inputted for each region, are presented in Figures 4 and 5, respectively. Using the same random effects model with DerSimonian-Laird estimation as in the nationwide analysis, the regional pooled IFR_1_ is estimated to be the following (as ordered from highest to lowest pooled effect size and reporting only regions with more than one state): East 0.136% (95% CI: 0.062 – 0.302), West 0.125% (95% CI: 0.052 – 0.303), North 0.109% (95% CI: 0.037 – 0.325), and South 0.062% (95% CI: 0.023 – 0.164) (Figure 4). Hence, East and West India have the largest estimated IFR_1_ of 0.136% and 0.125%, respectively, suggesting estimated infection fatality rates (when not adjusting for underreporting) for SARS-CoV-2 of 136 deaths per 100,000 persons and 125 deaths pers 100,000 persons in these regions, respectively. The estimate for the East is largely driven by the comparatively high statewide IFR_1_ estimate for West Bengal of 0.322% (95% CI: 0.290 – 0.357), whereas the estimate for the West is driven by Maharashtra (0.244%) (Figure 4). The lowest IFR_1_ estimate is for South India of 0.062 (95% CI: 0.023 – 0.164), backed by the Andhra Pradesh and Karnataka which have the lowest statewide pooled IFR_1_ of 0.019% and 0.019%, respectively (Figure 4). Also considering regions with 1 state, pooled regional IFR_1_ estimates range from 0.062 for South to 0.174% for Central.

**Figure 4.**
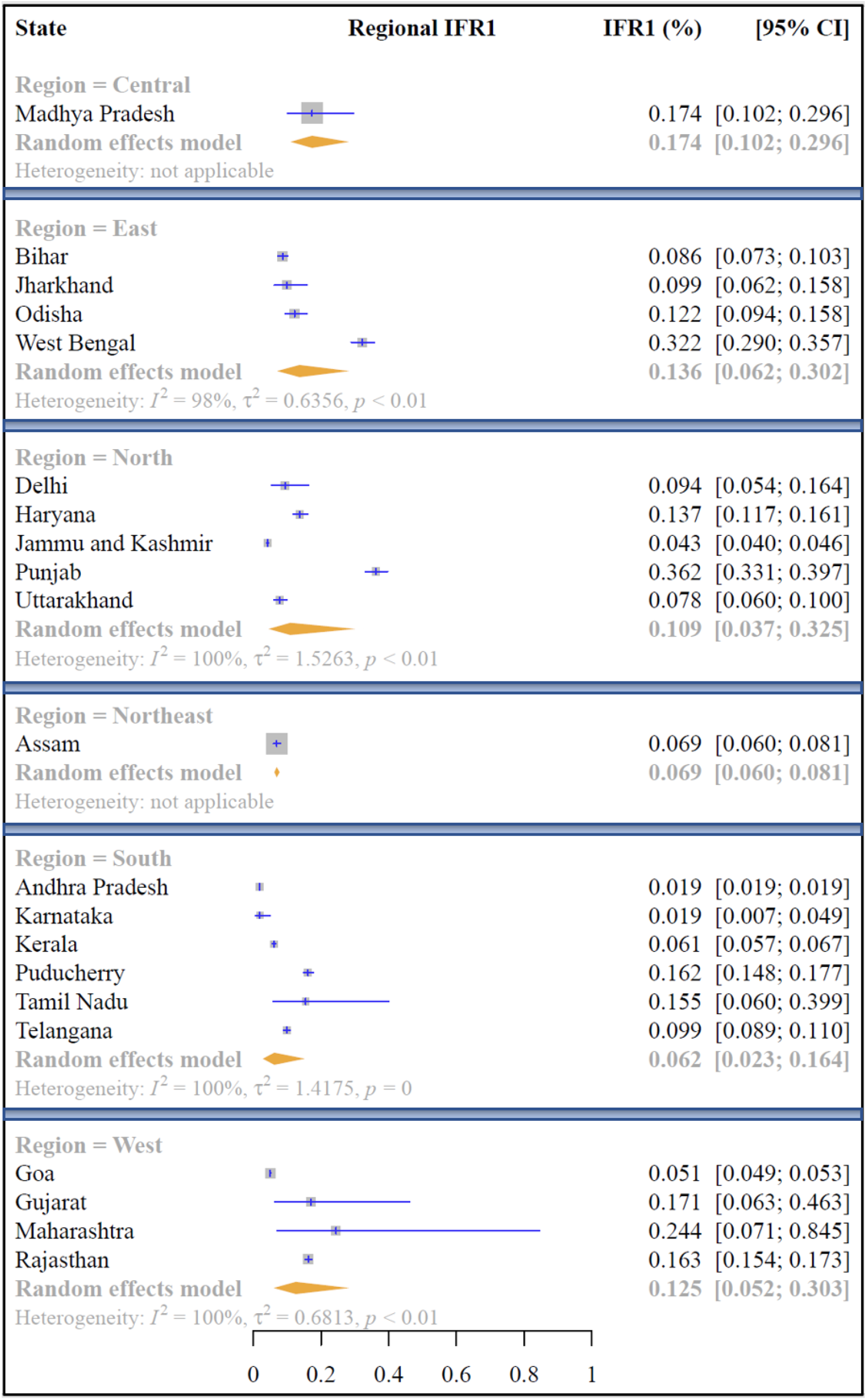
Regional estimated pooled IFR_1_ of SARS-CoV-2 for India. Note: * = IFR_1_ precalculated. ** = IFR_2_ precalculated. *** = both IFR_1_ and IFR_2_ precalculated. For those with no stars, IFR_1_ and IFR_2_ as well as 95% CIs were computed.

**Figure 5.**
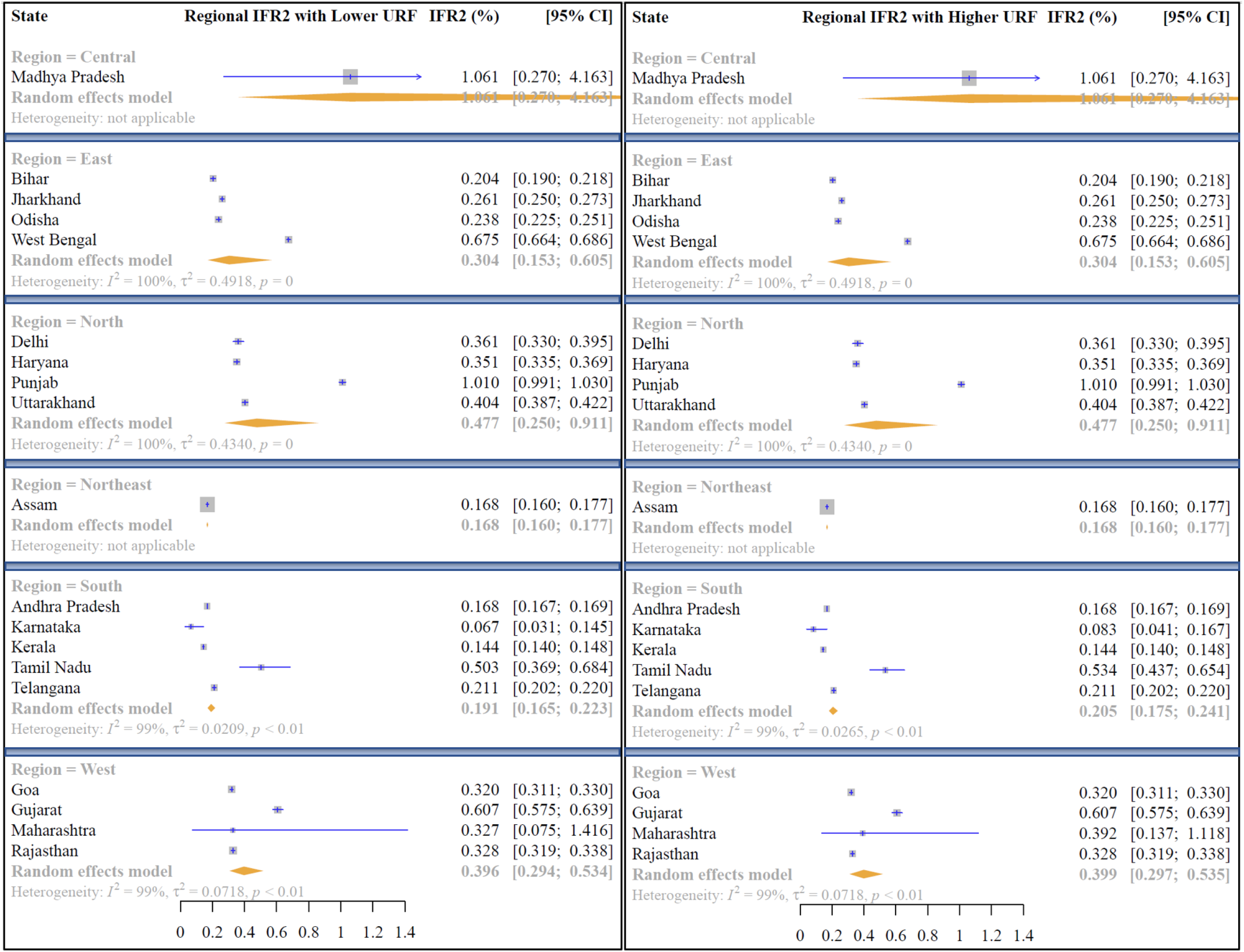
Regional estimated pooled IFR_2_ of SARS-CoV-2 for India, with lower and higher URF uncertainty. Note: State-specific IFRs in this forest plot are pooled estimates across included studies encompassed in each state.

We also find considerable heterogeneity exists even within each respective region in India. As examples, the statewide pooled IFR_1_ varies in the North from 0.043% (Jammu and Kashmir) to 0.362% (Punjab), in the South from 0.019% (Andhra Pradesh and Karnataka, respectively) to 0.162% (Puducherry), and in the West from 0.051% (Goa) to 0.244% (Maharashtra).

Using the same random effects model with DL estimation, the range of the regional pooled IFR_2_ is estimated to be the following (as ordered from highest to lowest pooled effect size and considering only regions with >1 state): North 0.477% (95% CI: 0.250 – 0.911) to 0.477% (95% CI: 0.250 – 0.911), West 0.396% (95% CI: 0.294 – 0.534) to 0.399% (95% CI: 0.297 – 0.535), East 0.304% (95% CI: 0.153 – 0.605) to 0.304% (95% CI: 0.153 – 0.605), and South 0.191% (95% CI: 0.165 – 0.223) to 0.205% (95% CI: 0.175 – 0.241) (Figure 5). Hence, North and West India have the largest estimated IFR_2_ of 0.477-0.477% and 0.396-0.399%, respectively, indicating estimated infection fatality rates (when adjusting for death underreporting) for SARS-CoV-2 of 477 deaths per 100,000 persons and 396-399 deaths per 100,000 persons in these regions, respectively. The Northern IFR_2_ estimate is largely driven by the comparatively high statewide IFR_2_ estimate for Punjab (1.010%) which has the second highest statewide pooled IFR_2_ (after Madhya Pradesh at 1.061%) of all states encompassed among the included studies, whereas the estimate for the West is driven by Gujarat (0.607%) (Figure 5). The lowest IFR_2_ estimate is again for South India of 0.191-0.205% (among regions with >1 state), backed by Karnataka and Kerala which have the lowest statewide pooled IFR_2_ of 0.067-0.083% and 0.144%, respectively (Figure 5). In sum, when considering regions with 1 state, pooled regional IFR_2_ estimates range from 0.168% for Northeast to 1.061% for Central.

A detailed forest plot of the state, city and district-specific seroprevalences of SARS-CoV-2 in India, as well as the accompanying 95% confidence interval, belying the computed regional and statewide IFRs, as directly reported in the included studies, can be found in Supplementary Appendix H Figure 1.

###### Key Findings

✤ **Regional pooled IFR1 estimates within India range from a low of 0.062% in the South to a high of 0.136% in the East. Regional pooled IFR2 estimates range from [0.191 – 0.205%] in the South to [0.477%] in the North.**
✤ **Considerable within region heterogeneity is also observed, as pooled state specific IFR1 and IFR2 estimates are evidenced to vary within regions.**

We present a funnel plot of the standard error of the effect sizes of the included studies in Figure 6 as a standard method for examining publication bias. Although the results show asymmetry, in the context of seroprevalence proportions from serosurveys, which comprise the bulk of the study designs, this does not necessarily indicate the presence of publication bias (*79*). Instead, we suspect that the asymmetry is due to heterogeneity in the actual effect sizes of the prevalence of SARS-CoV-2 among the populations captured by the included studies. A detailed assessment of publication bias is presented in Supplemental Appendix J. The results of the Joanna Briggs Institute (JBI) assessment for risk of bias within each included study can be found in Supplemental Appendix I.

**Figure 6.**
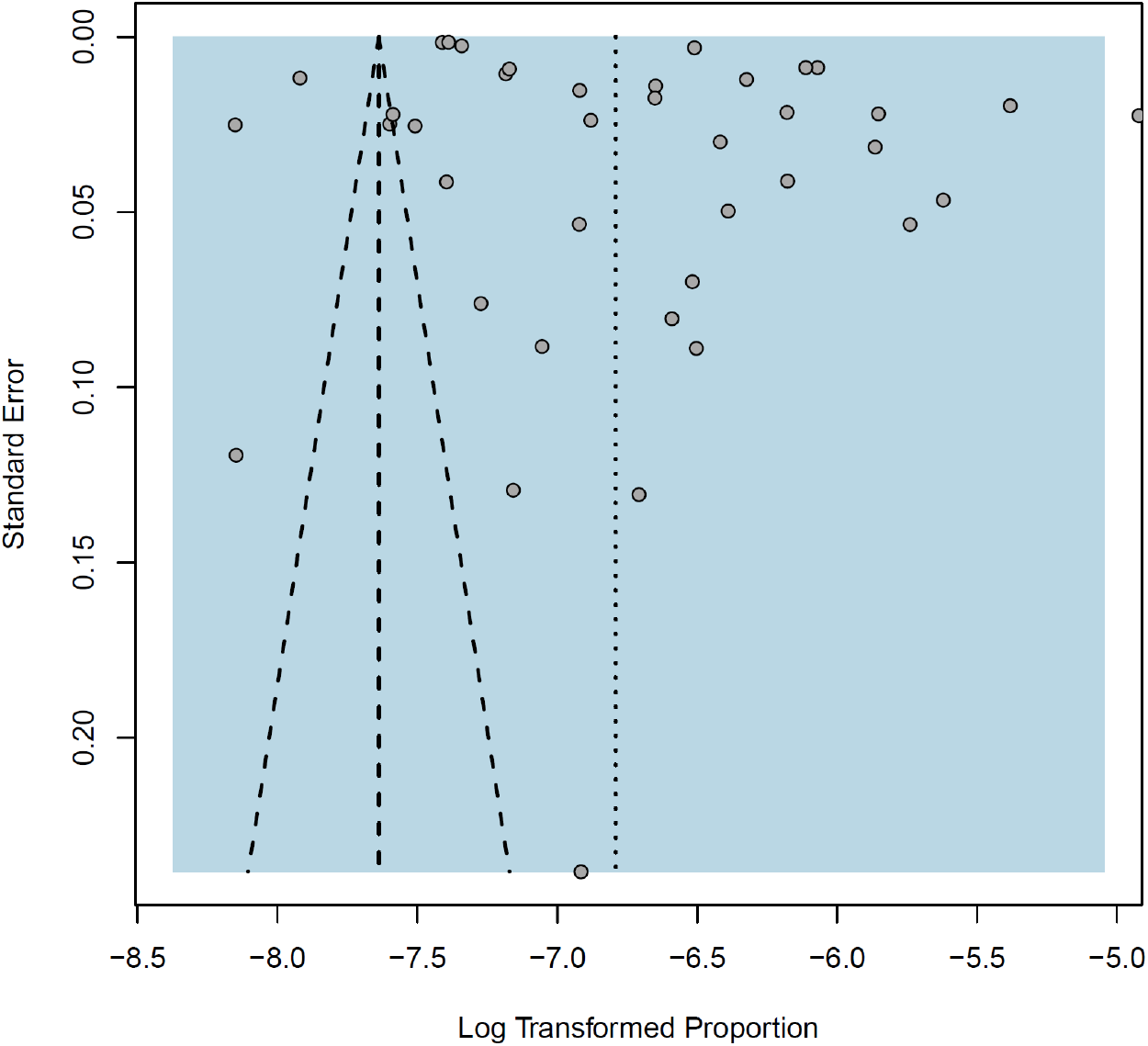
Funnel plot for examination of publication bias across included studies.

Data files and code used to produce the above results are available on GitHub at the following repository: https://github.com/lzimmermann4/india_c19_ifrs.

### Updated Model-based estimates of Wave 1 and Wave 2 Fatalities

Since wave 1 is defined analogously as in the latest report (*13*) and the underlying data are the same, the wave 1 estimates presented herein are unchanged. Briefly, when adjusting the denominator by the estimated cases underreporting factor of 11.11 (95% CrI 10.71 – 11.47), the nationwide IFR_1_ estimate (defined as observed cumulative deaths/estimated total cumulative infections) for wave 1 is 0.129% (95% CrI 0.125 – 0.134). When further adjusting the numerator by the estimated death underreporting factor of 3.56 (95% CrI 3.48 – 3.64), we obtain a nationwide IFR_2_ estimate (i.e., estimated cumulative total deaths/estimated cumulative total infections) for wave 1 of 0.461% (95% CrI 0.455 – 0.468). The wave 1 IFR_2_ estimate is largely congruent to the estimated effect sizes from the latest nationwide serosurvey in India (*59*) in that it falls within the bounds of the computed IFR_2_ herein for the 4^th^ nationwide serosurvey of 0.241-0.651%, when applying a death underreporting factor range of 4.4 (1.79 million excess deaths (*21*)) to 11.9 (4.9 million excess deaths (*23*)).

Since wave 2 data extend to June 30, 2021, using the same compartmental epidemiological model we estimate case and death underreporting factors of 13.3 (95% CrI 11.4 – 14.6) and 3.46 (95% CrI 3.13 – 3.69), respectively. As such, the nationwide IFR_1_ estimate for wave 2 is 0.095% (95% CrI 0.086 – 0.109), which is slightly higher than the previously reported estimate of 0.032% (95% CrI 0.029% – 0.035%) as of May 15, 2021 (i.e. near the peak of wave 2 in India) (*13*). The nationwide IFR_2_ estimate for wave 1 is 0.326% (95% CrI 0.316 – 0.342), which is higher than our latest reported estimate of 0.183% (95% CrI 0.180 – 0.186) as of May 15, 2021 (*13*). The current estimates of the IFR for wave 2 have increased in magnitude in comparison to than past estimates due to having a time period that better reflects the trajectory of the second wave and data that encompass both the ascent and descent of cases and deaths. Moreover, the updated underreporting factors for cases and deaths are half the size of previous underreporting factors for wave 2 (i.e. 26.73 (95% CrI 24.26 – 28.81) and 5.77 (95% CrI 5.34 – 6.15), respectively, as of May 15) (*13*) and have largely converged in magnitude to the underreporting factors for wave 1.

Figure 6 contains the case and death underreporting factors and estimated number of infections and deaths for waves 1 and 2, respectively, for India. State-level estimates of IFR_1_ and IFR_2_ for waves 1 and 2 are reported in ***Supplementary Appendix N*** Figure 2, which shows the between state variation in these measures for wave 1 and wave 2, respectively, for the 20 states with high cases-deaths within India.

**Figure 6.**
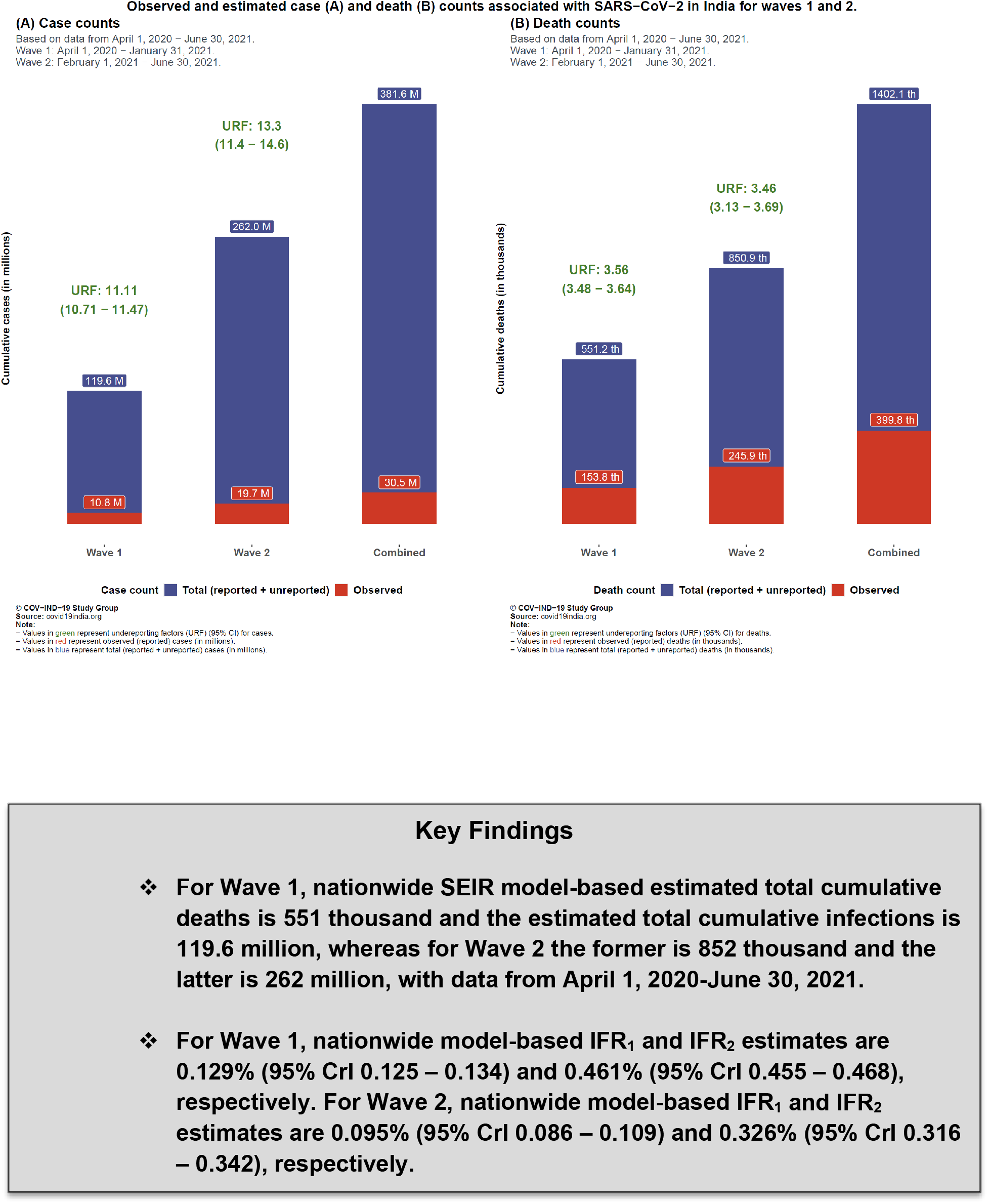
Comparison of observed and estimated case and death counts and corresponding underreporting factors from wave 1, wave 2 and both combined.

### Combined estimates across the waves

As shown in Figures 6 above, the total cases (reported and unreported) is estimated to be 381.6 million and the total deaths (reported and unreported) to be 1.4 million, across waves 1 and 2 for India. Subsequently, the combined infection fatality rate (IFR) across both waves is estimated to be 0.101% for IFR_1_, and 0.367% for IFR_2_ (as previously defined). State-level case and death underreporting factors for India are included in ***Supplementary Appendix N*** Tables 1-3 ***and*** Figure 3.

We also include a state and country level comparison of the estimated case underreporting factors across both waves to the estimated case under counting factors presented by ICSM in the 4^th^ nationwide seroprevalence survey (as of May 31, 2021) for India (*59*) (see ***Supplementary Appendix N*** Table 4). Briefly, the estimated case underreporting factors presented herein are lower at large than the case undercounting factors announced for the 4^th^ nationwide seroprevalence survey (*59*), with the exceptions of Andhra Pradesh, Chhattisgarh, Jammu and Kashmir, and Kerala where the case underreporting factors that we estimate are larger, as well as Maharashtra where the case underreporting factor is the same. As such, we present estimated case underreporting factors that are conservative with respect to the recent nationwide serosurvey under counting estimates. The true extent of the magnitude of infections absent in reported case counts is wagered to lie somewhere in between.

#### Key Findings

✤ **As of 30^th^ June 2021, nationwide URF (C) and URF (D) are estimated to be 12.5 and 3.5, respectively by the SEIR model.**
✤ **Nationwide SEIR model-based estimated total cumulative infections is 381.6 million and estimated total cumulative deaths is 1.4 million (as of 30^th^ June 2021), whereas reported deaths were 412 thousand and cases were at 30.4 million.**
✤ **Nationwide model-based cumulative IFR1 and IFR2 estimates are 0.101% (95% CI: 0.097 – 0.116) and 0.367% (95% CI: 0.358 – 0.383), respectively.**
✤ **The magnitude of these model-based estimates largely reconcile with the empirical findings. Across empirical data and model-based estimation, IFR2 is nearly 3.6 times the size of IFR_1_.**

## DISCUSSION AND CONCLUSIONS

### Neighboring Countries

Despite there being insufficient data and sparse eligible studies in the neighboring countries, a brief snapshot into the SARS-CoV-2 situation within each country shows that the case fatality rate (unadjusted for underreporting) in neighboring countries is at par with India, if not higher. In Table 5, we provide a comparison of nationwide CFRs, as of 31 July 2021, within each of the countries included in this review (i.e., India, Bangladesh, Nepal, Pakistan, and Sri Lanka). Of these five countries, Pakistan stands out as having the highest CFR at 2.37% with 1.02 million cumulative reported infections and 24 thousand cumulative reported deaths (31 July 2021). Whereas India in fact has the lowest CFR of these 5 countries at 1.36%, but less surprisingly given its large population has the highest reported death and confirmed infection toll of 430 thousand and 31.6 million (31 July 2021), respectively. We recognize and caution that the CFR is an imperfect measure of the true SARS-CoV-2 fatality rate and that levels of underreporting of deaths and cases vary by country (*8*). In sum, Table 5 also presents a CFR across the countries included in this systematic review which is 1.42% to summarize the observed mortality rate in this region. The CFR among the neighboring countries alone is 1.96% which indicates that the SARS-CoV-2 observed mortality rate of the neighboring countries appears to be similar to that of India, if not slightly higher (whilst cautioning the fog of undercounting).

**Table 5.** Comparison of nationwide CFRs among countries in review, as of July 2021.

| Country (Population)<sup>a</sup> | As of July 31, 2021 |  |  |
| --- | --- | --- | --- |
|  | Reported Deaths<sup>b</sup> | Reported Cases<sup>c</sup> | Case Fatality Rate (%) |
| Pakistan (216 million) | 24,266 | 1,024,861 | 2.37 |
| Bangladesh (163 million) | 23,988 | 1,249,484 | 1.92 |
| Sri Lanka (21 million) | 5,878 | 308,812 | 1.90 |
| Nepal (28 million) | 10,259 | 695,389 | 1.48 |
| India (1.36 billion) | 430,732 | 31,613,993 | 1.36 |
| Region excluding India (430 million) | 64,391 | 3,278,546 | 1.96 |
| Region including India (1.796 billion) | 495,123 | 34,892,539 | 1.42 |

Note: Case Fatality Rates (CFR) are computed as the Reported Deaths divided by Reported Cases. Regional CFRs are unweighted.
[a] Population estimates are projected for the year 2019 from World Bank
(https://data.worldbank.org/indicator/SP.POP.TOTL?locations=BD-NP-PK-LK-IN) (80).
[b] Reported Deaths were collected cumulatively 14 days after the date July 31, 2021 (i.e. August 14, 2021) from Worldometer COVID-19 Data (https://www.worldometers.info/coronavirus/) (81).
[c] Reported Cases were collected cumulatively for the date July 31, 2021 from Worldometer COVID-19 Data (https://www.worldometers.info/coronavirus/) (81).

### Summary on fatality rates in India

For India, there were enough eligible studies to perform a meta-analysis of infection fatality rates. A key result of the meta-analysis concerning the nationwide IFR estimates for India is that both the CFR (i.e., not accounting for infection nor death underreporting) and IFR_1_ (i.e., not accounting for death underreporting) have decreased over time, whereas IFR_2_ appears to have remained same or even increased in some studies over time. In other words, when failing to take into account case and/or death underreporting (i.e., CFR and IFR_1_), the SARS-CoV-2 mortality rate falsely appears to be decreasing. However, when correctly adjusting for both underreporting of infections and deaths (i.e., IFR_2_), we do not observe that less deaths are resulting from the same number of infections through wave 2 compared to wave 1. This aligns with the catastrophic collapse of the healthcare system observed at large in India during the second wave of the pandemic (*2*). In sum, use of either the CFR or IFR_1_ (not accounting for death underreporting) alone may be deceptively misleading, and IFR_2_ should also be utilized in public health programming and policy decision making.

The narrative review of general population SARS-CoV-2 infection fatality rate or seroprevalence studies brought forth some trends concerning virus spread and fatality. In India, dense urban areas and slum dwellings have been found to have markedly higher prevalence rates in comparison to non-slum or rural locations (e.g., 58.4% (95% CI: 56.8– 59.9) among slums versus 17.3% (95% CI: 16.0–18.7) for Mumbai (*58*) among non-slums, and 57.9% (95% CI: 53.4–62.3) among a slum in Bengaluru (*51*)). Trends in the infection fatality rates (IFRs) are less obvious among the albeit limited number of slum population-focused seroprevalence studies in this region. A study in Mumbai found the IFR of SARS-CoV-2 in slums to be one-third the size of that of non-slums (i.e. 0.076% among slums versus 0.263% among non-slums (*58*)). Similarly, another study on a slum in Bengaluru estimated the IFR to be 2.94 deaths per 10,000 infections (or 0.029%) (*51*).

When estimating regional and statewide IFR estimates for India, the latest available studies for certain regions was during the outset of the pandemic (e.g., Central India with both input studies around August 2020). Such variation in study time periods highlights the need for updated seroprevalence studies in select regions. The findings from this regional meta-analysis also underscore the geographical differences in data collection/research spread across India. A large number of the included studies in this systematic review and meta-analysis were in the South and North, with substantially less coverage in the Central and Northeast regions. Moreover, no eligible serosurveys were available for the region Northeast India (which includes the following states: Arunachal Pradesh, Assam, Manipur, Meghalaya, Mizoram, Nagaland, Sikkim and Tripura). This leads us to the limitations of the meta-analysis presented herein.

### Limitations of Meta-analysis

We were not able to incorporate Bangladesh, Nepal, Pakistan, nor Sri Lanka in the meta-analysis presented in this paper as no eligible studies were available for these countries (aside from Pakistan, which had 2 included seroprevalence surveys), as previously discussed in the *Results* section. There were insufficient age-as well as sex-disaggregated IFRs for India (and for the neighboring countries) to examine heterogeneity in IFR estimates by either demographic. Furthermore, although age and sex-disaggregated seroprevalence estimates were extracted from the included studies, disaggregated deaths and cases by these demographics are not available for India nor for its states, cities, nor districts, at the time of this review. Additionally, there were insufficient data on excess deaths for select states (e.g., Jammu and Kashmir and Puducherry) that precluded our ability to compute IFR_2_ for studies encompassed in these states and, as such, we were unable to include these studies in the meta-analysis for the regional IFR_2_. Additionally, we caution that several of the included studies in the quantitative synthesis are in the process of being peer-reviewed, and so multiple underlying datapoints have not yet been verified.

Lastly, we note that global studies that examine India along with multiple other countries (as part of a country level global analysis) but do not explicitly refer to India in the title/abstract would not be captured by our published search strategy. Therefore, a limitation of the meta-analysis is that these IFR estimates for India from global studies are not comprehensively reflected in the empirical results of IFRs and similarly in the excess deaths synthesis. For example, a recent study by Rahmandad et al. (2021) (*82*) that *appears* (exact estimates are not reported) to estimate for India a ratio of estimated to reported deaths on the log scale in the ballpark of 1.2-1.5 (i.e. 3.3-4.4 when exponentiated) and an IFR near 0.4-0.5% for India (as of 22 December 2020), as part of a global analysis of 92 nations, was not captured (no explicit reference to India in title/abstract). Hence, this is an example of model-based estimates not reflected in the empirical findings evidenced in this report.

### Recommendations moving forward

Based on the empirical and model-based findings evidenced in this report, we provide some high-level recommendations, primarily, as it concerns India. To reiterate, investigations aimed at estimating the true extent of COVID-19 attributed deaths in India have relied on a myriad of data sources (Table A). Such data sources do not come without their challenges (e.g., non-response bias and selection bias), and often lead to considerable variation in the ultimate results. For example, Ramachandran and Malani (2021) provide a higher estimate of excess deaths for India of 5.21 million (compared to their 3.36 million excess deaths estimate) for January 2020 to June 2021, when further considering households that do not respond consecutively (*30*). Given that no current all-cause mortality data exist for India at the time of this report with the latest release in 2010-2013, linking data sources in this manner is a common alternative. We encourage the release of timely and disaggregated data on SARS-CoV-2 cases and deaths, as are needed to assess stratified effects by age-sex-geography(*5*). Despite geographic heterogeneity, quality of case and death reporting remains of great concern for India and continues to mask the true fatality rate as well as impede ongoing epidemiological investigations. In the absence of all-cause and disaggregated COVID-19 mortality data, tools (e.g., a personal digital health identifier) (*5*) and fortified vital surveillance methods (e.g., monitoring inactive bank accounts) and practices (e.g., increased community engagement) (*5*) are needed to facilitate and validate the use of the previously mentioned data sources and others. Strengthening the nationwide vital surveillance system in these ways would enable timely and pointed interventions from public health and government officials to ultimately prevent further overload of healthcare systems and loss of life. To put the excess deaths synthesis into context, the range of death underreporting factor for India of 4.4-11.9 (based on 1.79-11.9 million excess deaths from (*21*) and (*23*), respectively) falls toward the higher end of the global death undercounting factor of 3.3 (95% CI: 2.1-4) estimated by the Economist (as of May 2021) (*83*). This indicates that the extent of death underreporting, as it pertains to SARS-CoV-2 remains particularly acute for India as compared to that of globally.

### Why death matters?

From an economic perspective, why is it necessary to do these mortality calculations? Death matters because life matters – prolonging life is a goal, not merely a means to generate economic value. The widely used human development index (HDI) of the World Bank takes a multi-dimensional approach to measuring welfare. Material wellbeing (GDP) is only one of its three components; the others involve knowledge (literacy rate) and life (life expectancy at birth).

Nevertheless, for the purpose of cost-benefit analysis and economic evaluation of public health measures, it is useful to be able to assign a monetary value to every life lost or saved. Measuring the value of a life by lost earnings or productivity is deeply flawed – it would, for example, have the absurd implication that the lives of non-working individuals have no value at all. The most commonly used approach to estimating the value of a statistical life (VSL) adopts the revealed preference or willingness-to-pay (WTP) principle. Hedonic wage regressions estimate the compensating differential (*84*) in market wages for jobs that involve a higher probability of fatal accidents – a measure of how much income the average worker is willing to sacrifice in order to reduce mortality risk. Sacrificed earnings are then aggregated over the lifetime using present discounted values and scaled up to a probability of 1 to arrive at VSL.

There are several estimates of VSL from developed countries^1^ but studies on developing countries including India are few and far between. For our purposes, we will use Majumder and Madheswaran (2018) (*85*) who estimate VSL for the average male industrial worker in India to be INR 44.69 million (US $0.61 million at current exchange rates)^2^. This may introduce upward as well as downward biases into our calculations. For example, people dying of COVID-19 are, on average, older than the industrial workforce, which implies the mortality cost presented below is an overestimate. On the other hand, wages are only one part of a worker’s economic contribution. A proper accounting of the social value of a life must also account for profits, rents and consumers’ surplus generated by the economic activity of that individual, as well as non-pecuniary benefits conferred on relatives, friends and acquaintances. These are not reflected in private WTP and are therefore a source of underestimating VSL. The calculations presented below are ballpark figures that will reflect reality closely if the above-mentioned biases largely cancel each other out.

Table 6 below monetizes the death toll of COVID-19 in India till June 2021. Based on reported death figures alone, the economic cost of Covid-induced mortality comes to around 9% of India’s annual GDP. If we use the model-based estimates or the lower end of the range of excess death estimates, we incur a notional cost of 30-40% of annual GDP. Note that the official decline in India’s GDP in the fiscal year 2020-21 is 7.7%, which is a gross underestimate of the true loss once the value of lives is taken into account. By various estimates, the fiscal outlay of the government in its financial rescue package was about 1% of GDP. This is a very small fraction of what is at stake in this pandemic. Total healthcare spending in 2018 by the central and state governments taken together was only 1.28% of GDP, which again illustrates the serious under-investment in mitigating health shocks like COVID-19.

**Table 6.**
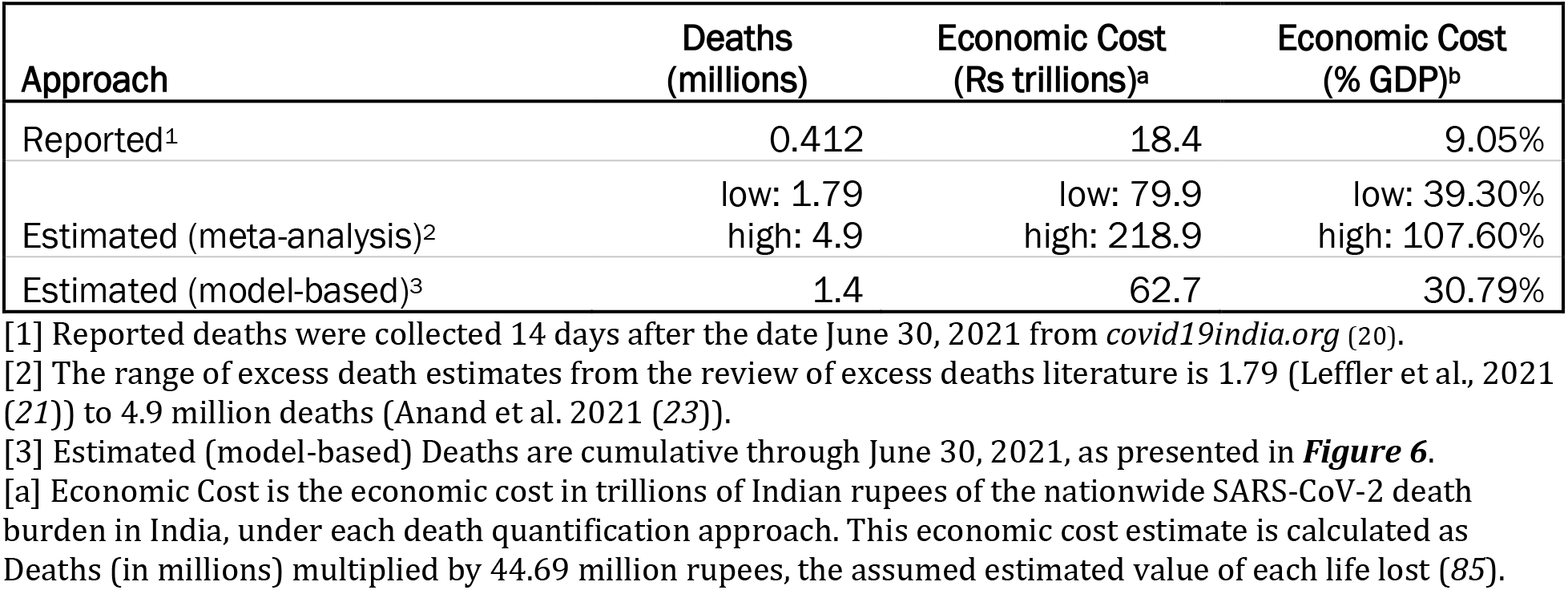
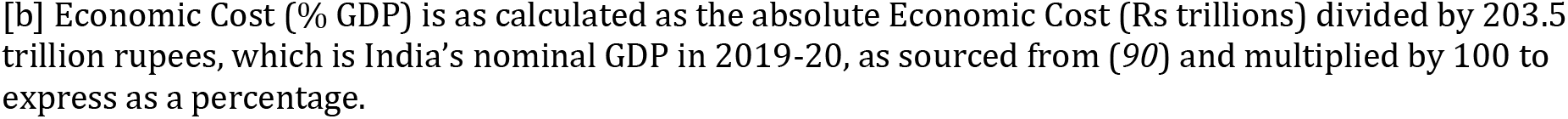
Summary of economic cost estimates attributed to SARS-CoV-2 death toll in India, as of June 2021.

Other than national estimates of mortality and IFR, there is considerable value to measuring infections and deaths across gender, caste, income levels, regions, etc. These can be useful guides for policymakers, telling them where scarce healthcare resources and aid should be concentrated. The within-country heterogeneity in both the epidemiological and economic impact of the disease has been noted in studies from all parts of the world. Unfortunately, absence of disaggregated data has tied the hands of researchers in India. Nevertheless, our meta-analysis uncovers substantial regional variations in IFR_2_, from a low of 0.191 – 0.205% in the South to a high of 0.477% in the North. These differences are plausibly created by socio-economic factors (like differences in quality of governance and healthcare) rather than biological ones and provide policymakers the information necessary for welfare arbitrage and efficient resource allocation.

In closing, premature mortality is not only an indicator of health but also of economic cost and productivity, as discussed above and as has been evidenced for other health conditions in India (*91*). Furthermore, unforeseen death within a family or community may disturb the livelihood and income generation structure, thereby pervading to the broader welfare of impacted individuals, although reverse causation is present (*92*). Aside from the health lens, premature mortality and infectious disease mortality’s consequences for economic stability and ensuing welfare policy further support the need for continued attention to assessing the mortality burden of COVID-19 within India.

## Supporting information

Supplemental RiskofBias

## Data Availability

Data files and code used to produce the results herein are available on GitHub at the below repository.

https://github.com/lzimmermann4/india_c19_ifrs

## Acknowledgments

The authors thank the librarians from the University of Michigan Taubman Health Sciences Library for their instruction on developing the search strategy for this rapid review.

## Author Contributions

Conceptualization: LZ, BM

Methodology: LZ, BM

Investigation: LZ, SB, SP, RK, RB, PG, BM

Supervision: BM

Writing – original draft: LZ, BM

Writing – review & editing: LZ, SB, SP, RK, RB, PG, BM

## Supplementary Appendices

## Appendix A. PRISMA checklist *[1]*

| Section and Topic | Item # | Checklist item | Location where item is reported |
| --- | --- | --- | --- |
| **TITLE** |  |  |  |
| Title | 1 | Identify the report as a systematic review. | Title Page |
| **ABSTRACT** |  |  |  |
| Abstract | 2 | Background: Provide an explicit statement of the main objective(s) or question(s) the review addresses. Methods: Specify the inclusion and exclusion criteria for the review. Specify the information sources (e.g. databases, registers) used to identify studies and the date when each was last searched. Specify the methods used to assess risk of bias in the included studies. Specify the methods used to present and synthesise results. Results: Give the total number of included studies and participants and summarise relevant characteristics of studies. Present results for main outcomes, preferably indicating the number of included studies and participants for each. If meta-analysis was done, report the summary estimate and confidence/credible interval. If comparing groups, indicate the direction of the effect (i.e. which group is favoured). Discussion: Provide a brief summary of the limitations of the evidence included in the review (e.g. study risk of bias, inconsistency and imprecision). Provide a general interpretation of the results and important implications. Other: Specify the primary source of funding for the review. Provide the register name and registration number. | Title Page |
| **INTRODUCTION** |  |  |  |
| Rationale | 3 | Describe the rationale for the review in the context of existing knowledge. | 3-6 |
| Objectives | 4 | Provide an explicit statement of the objective(s) or question(s) the review addresses. | 6 |
| **METHODS** |  |  |  |
| Eligibility criteria | 5 | Specify the inclusion and exclusion criteria for the review and how studies were grouped for the syntheses. | 11-12 |
| Information sources | 6 | Specify all databases, registers, websites, organisations, reference lists and other sources searched or consulted to identify studies. Specify the date when each source was last searched or consulted. | 10-11, App B |
| Search strategy | 7 | Present the full search strategies for all databases, registers and websites, including any filters and limits used. | App B |
| Selection process | 8 | Specify the methods used to decide whether a study met the inclusion criteria of the review, including how many reviewers screened each record and each report retrieved, whether they worked independently, and if applicable, details of automation tools used in the process. | 13, App B |
| Data collection process | 9 | Specify the methods used to collect data from reports, including how many reviewers collected data from each report, whether they worked independently, any processes for obtaining or confirming data from study investigators, and if applicable, details of automation tools used in the process. | 13, App B |
| Data items | 10a | List and define all outcomes for which data were sought. Specify whether all results that were compatible with each outcome domain in each study were sought (e.g. for all measures, time points, analyses), and if not, the methods used to decide which results to collect. | 7-10, 13 |
|  | 10b | List and define all other variables for which data were sought (e.g. participant and intervention characteristics, funding sources). Describe any assumptions made about any missing or unclear information. | 7-10, 13 |
| Study risk of bias assessment | 11 | Specify the methods used to assess risk of bias in the included studies, including details of the tool(s) used, how many reviewers assessed each study and whether they worked independently, and if applicable, details of automation tools used in the process. | 15 |
| Effect measures | 12 | Specify for each outcome the effect measure(s) (e.g. risk ratio, mean difference) used in the synthesis or presentation of results. | 14, App F |
| Synthesis methods | 13a | Describe the processes used to decide which studies were eligible for each synthesis (e.g. tabulating the study intervention characteristics and comparing against the planned groups for each synthesis (item #5)). | App F, App G |
|  | 13b | Describe any methods required to prepare the data for presentation or synthesis, such as handling of missing summary statistics, or data conversions. | 14, App F, App G |
|  | 13c | Describe any methods used to tabulate or visually display results of individual studies and syntheses. | 13-15, App F |
|  | 13d | Describe any methods used to synthesize results and provide a rationale for the choice(s). If meta-analysis was performed, describe the model(s), method(s) to identify the presence and extent of statistical heterogeneity, and software package(s) used. | 13-15, App F |
|  | 13e | Describe any methods used to explore possible causes of heterogeneity among study results (e.g. subgroup analysis, meta-regression). | 13-15 |
|  | 13f | Describe any sensitivity analyses conducted to assess robustness of the synthesized results. | 15 |
| Reporting bias assessment | 14 | Describe any methods used to assess risk of bias due to missing results in a synthesis (arising from reporting biases). | 15 |
| Certainty assessment | 15 | Describe any methods used to assess certainty (or confidence) in the body of evidence for an outcome. | 10, 15 |
| **RESULTS** |  |  |  |
| Study selection | 16a | Describe the results of the search and selection process, from the number of records identified in the search to the number of studies included in the review, ideally using a flow diagram. | 17-19 |
|  | 16b | Cite studies that might appear to meet the inclusion criteria, but which were excluded, and explain why they were excluded. | 22, App D, App E, App G |
| Study characteristics | 17 | Cite each included study and present its characteristics. | 19-22 |
| Risk of bias in studies | 18 | Present assessments of risk of bias for each included study. | App I |
| Results of individual studies | 19 | For all outcomes, present, for each study: (a) summary statistics for each group (where appropriate) and (b) an effect estimate and its precision (e.g. confidence/credible interval), ideally using structured tables or plots. | 20, App C |
| Results of syntheses | 20a | For each synthesis, briefly summarise the characteristics and risk of bias among contributing studies. | 37, App I |
|  | 20b | Present results of all statistical syntheses conducted. If meta-analysis was done, present for each the summary estimate and its precision (e.g. confidence/credible interval) and measures of statistical heterogeneity. If comparing groups, describe the direction of the effect. | 29-37 |
|  | 20c | Present results of all investigations of possible causes of heterogeneity among study results. | 33-36 |
|  | 20d | Present results of all sensitivity analyses conducted to assess the robustness of the synthesized results. | 37-38, App H |
| Reporting biases | 21 | Present assessments of risk of bias due to missing results (arising from reporting biases) for each synthesis assessed. | 37, App J |
| Certainty of evidence | 22 | Present assessments of certainty (or confidence) in the body of evidence for each outcome assessed. | 37, App J |
| **DISCUSSION** |  |  |  |
| Discussion | 23a | Provide a general interpretation of the results in the context of other evidence. | 42-45 |
|  | 23b | Discuss any limitations of the evidence included in the review. | 45-46 |
|  | 23c | Discuss any limitations of the review processes used. | 45-46 |
|  | 23d | Discuss implications of the results for practice, policy, and future research. | 46-50 |
| **OTHER INFORMATION** |  |  |  |
| Registration and protocol | 24a | Provide registration information for the review, including register name and registration number, or state that the review was not registered. | NA |
|  | 24b | Indicate where the review protocol can be accessed, or state that a protocol was not prepared. | NA |
|  | 24c | Describe and explain any amendments to information provided at registration or in the protocol. | NA |
| Support | 25 | Describe sources of financial or non-financial support for the review, and the role of the funders or sponsors in the review. | Title Page |
| Competing interests | 26 | Declare any competing interests of review authors. | Title Page |
| Availability of data, code and other materials | 27 | Report which of the following are publicly available and where they can be found: template data collection forms; data extracted from included studies; data used for all analyses; analytic code; any other materials used in the review. | 38, App C |

NA = not applicable.

## Appendix B. Systematic review search procedure

To conduct the present systematic review, we systematically collected publications and preprints concerning infection fatality rates (IFRs) from SARS-CoV-2 in India and neighboring countries of Bangladesh, Nepal, Pakistan, and Sri Lanka. To identify relevant papers, we searched four databases: PubMed, Embase, Global Index Medicus, and isearch for preprints (encompassing bioRxiv, medRxiv, and SSRN). The search was conducted on July 3, 2021 and, as such, the results reflect published studies and preprints available from January 1, 2020 to July 3, 2021. Results were further verified through August 15, 2021 through reviewing media reports, government press releases, and manual search of preprints and publications. Data were extracted from the online search engines into the reference manager Zotero, deduplicated, and imported into Excel for screening.

The title/abstract screening, the full-text screening, and data abstraction were independently performed by two screeners to verify which studies met inclusion and exclusion criteria and to very data collected. When the two screeners disagreed on the marking for a citation, the screeners reached a consensus on whether to advance the citation to the next level of screening. Below we publish the full search strategies for each database.

**PubMed (National Library of Medicine)**

**<u>Date searched:</u>** 7/3/2021

**<u>Number of results:</u>** 2,940

**<u>Date filter:</u>** January 1, 2020 to [blank]

**<u>Other filters applied:</u>** None

1.

covid-19[tw] OR COVID19[tw] OR SARS-CoV-2[tw] OR SARS-CoV2[tw] OR severe acute respiratory syndrome coronavirus 2[tw] OR 2019-nCoV[tw] OR 2019nCoV[tw] OR coronavirus[tw] OR coronavirus[mh] OR covid-19[mh]

2.

india[text word] OR india[mesh] OR indian[text word] OR pakistan[text word] OR pakistani[text word] OR pakistan[mesh] OR bangladesh[text word] OR bangladeshi[text word] OR bangladesh[mesh] OR nepal[text word] OR nepal[mesh] OR “sri lanka”[text word] OR “sri lankan”[text word] OR “sri lanka”[mesh]

3.

IFR[text word] OR infection*[text word] OR CFR[text word] OR case*[text word] OR transmission*[text word] OR mortalit*[text word] OR mortality[mesh] OR fatalit*[text word] OR lethalit*[text word] OR death*[text word] OR burden[text word] OR underreporting[text word] OR “under-reporting”[text word] OR seroprevalence[text word] OR serosurvey[text word] OR serology[text word] OR serology[mesh] OR seroconversion[text word] OR seroconversion[mesh] OR “serosurveillance”[text word] OR Seroepidemiologic studies[mesh] OR seroepid*[text word] OR seropositiv*[text word] OR antibod*[text word] OR

antibodies[mesh] OR surveillance[text word] OR SIR[text word] OR SEIR[text word] OR “susceptible-exposed-infected-removed”[text word] OR “susceptible-infected-removed”[text word]

(1 AND 2 AND 3)

**Embase (Elsevier)**

**<u>Date searched:</u>** 7/3/2021

**<u>Number of results:</u>** 1,119

**<u>Date filter:</u>** 2020 to 2021

**<u>Other filters applied:</u>** Embase only and not Medline (as Medline is included in PubMed)

1.

covid-19:ti,ab,kw OR COVID19:ti,ab,kw OR SARS-CoV-2:ti,ab,kw OR SARS-CoV2:ti,ab,kw OR “severe acute respiratory syndrome coronavirus 2”:ti,ab,kw OR 2019-nCoV:ti,ab,kw OR 2019nCoV:ti,ab,kw OR coronavirus:ti,ab,kw OR ‘Coronavirinae’/exp OR ‘coronavirus disease 2019’/exp

2.

india:ti,ab,kw OR ‘india’/exp OR indian:ti,ab,kw OR pakistan:ti,ab,kw OR pakistani:ti,ab,kw OR ‘pakistan’/exp OR bangladesh:ti,ab,kw OR bangladeshi:ti,ab,kw OR ‘bangladesh’/de OR nepal:ti,ab,kw OR ‘nepal’/de OR “sri lanka”:ti,ab,kw OR “sri lankan”:ti,ab,kw OR ‘sri lanka’/de

3.

IFR:ti,ab,kw OR infection*:ti,ab,kw OR CFR:ti,ab,kw OR case*:ti,ab,kw OR transmission*:ti,ab,kw OR mortalit*:ti,ab,kw OR ‘mortality’/exp OR fatalit*:ti,ab,kw OR lethalit*:ti,ab,kw OR death*:ti,ab,kw OR burden:ti,ab,kw OR underreporting:ti,ab,kw OR under-reporting:ti,ab,kw OR seroprevalence:ti,ab,kw OR serosurv*:ti,ab,kw OR serology:ti,ab,kw OR ‘serology’/exp OR seroconversion:ti,ab,kw OR ‘seroconversion’/de OR ‘Seroepidemiology’/exp OR seroepid*:ti,ab,kw OR seropositiv*:ti,ab,kw OR antibod*:ti,ab,kw OR ‘antibody’/exp OR surveillance:ti,ab,kw OR SIR:ti,ab,kw OR SEIR:ti,ab,kw OR susceptible-exposed-infected-removed:ti,ab,kw OR susceptible-infected-removed:ti,ab,kw

(1 AND 2 AND 3)

**isearch (National Library of Medicine)**

**<u>Date searched:</u>** 7/3/2021

**<u>Number of results:</u>** 1,078

**<u>Date filter:</u>** January 1, 2020 to [blank]

**<u>Other filters applied:</u>** filtered to facets bioRxiv, medRxiv, SSRN; searched through title and abstract.

Note: Since isearch is curated to include COVID-19 related studies only, we translated the PubMed concept blocks 2 and 3.

(india OR indian OR pakistan OR pakistani OR bangladesh OR bangladeshi OR nepal OR “sri lanka” OR “sri lankan”) AND (IFR OR infection* OR CFR OR case* OR transmission* OR mortalit* OR fatalit* OR lethalit* OR death* OR burden OR underreporting OR “under-reporting” OR seroprevalence OR serosurvey OR serology OR seroconversion OR “serosurveillance” OR seroepid* OR seropositiv* OR antibod* OR surveillance OR SIR OR SEIR OR “susceptible-exposed-infected-removed” OR “susceptible-infected-removed”)

**Global Index Medicus--SEAR & EMR (World Health Organization)**

*Note:* We searched in IMSEAR (Index Medicus for the South-East Asia Region) for India, Bangladesh, Nepal, and Sri Lanka, and we searched in IMEMR (Index Medicus for the Eastern Mediterranean Region) for Pakistan.

**For IMSEAR:**

**<u>Date searched:</u>** 7/3/2021

**<u>Number of results:</u>** 35

**<u>Date filter:</u>** 2020 to [blank]

**<u>Other filters applied:</u>** Index filtered to “IMSEAR (South-East Asia)”; searched through title, abstract, subject.

(tw:(covid-19) OR tw:(COVID19) OR tw:(SARS-CoV-2) OR tw:(SARS-CoV2) OR tw:(“severe acute respiratory syndrome coronavirus 2”) OR tw:(2019-nCoV) OR tw:(2019nCoV) OR tw:(coronavirus)) AND (tw:(india) OR tw:(indian) OR tw:(bangladesh) OR tw:(bangladeshi) OR tw:(nepal) OR tw:(“sri lanka”) OR tw:(“sri lankan”)) AND (tw:(IFR) OR tw:(infection*) OR tw:(CFR) OR tw:(case*) OR tw:(transmission*) OR tw:(mortalit*) OR tw:(fatalit*) OR tw:(lethalit*) OR tw:(death*) OR tw:(burden) OR tw:(underreporting) OR tw:(“under-reporting”) OR tw:(seroprevalence) OR tw:(serosurvey) OR tw:(serology) OR tw:(seroconversion) OR tw:(“serosurveillance”) OR tw:(seroepid*) OR tw:(seropositiv*) OR tw:(antibod*) OR tw:(surveillance) OR tw:(SIR) OR tw:(SEIR) OR tw:(“susceptible-exposed-infected-removed”) OR tw:(“susceptible-infected-removed”))

**For IMEMR:**

**<u>Date searched:</u>** 7/3/2021

**<u>Number of results:</u>** 2

**<u>Date filter:</u>** 2020 to [blank]

**<u>Other filters applied:</u>** Index filtered to “IMEMR (Eastern Mediterranean)”; searched through title, abstract, subject.

(tw:(covid-19) OR tw:(COVID19) OR tw:(SARS-CoV-2) OR tw:(SARS-CoV2) OR tw:(“severe acute respiratory syndrome coronavirus 2”) OR tw:(2019-nCoV) OR tw:(2019nCoV) OR tw:(coronavirus)) AND (tw:(pakistan) OR tw:(pakistani)) AND (tw:(IFR) OR tw:(infection*) OR tw:(CFR) OR tw:(case*) OR tw:(transmission*) OR tw:(mortalit*) OR tw:(fatalit*) OR tw:(lethalit*) OR tw:(death*) OR tw:(burden) OR tw:(underreporting) OR tw:(“under-reporting”) OR tw:(seroprevalence) OR tw:(serosurvey) OR tw:(serology) OR tw:(seroconversion) OR tw:(“serosurveillance”) OR tw:(seroepid*) OR tw:(seropositiv*) OR tw:(antibod*) OR tw:(surveillance) OR tw:(SIR) OR tw:(SEIR) OR tw:(“susceptible-exposed-infected-removed”) OR tw:(“susceptible-infected-removed”))

## Appendix C. Summary of included articles

**Table 1.**
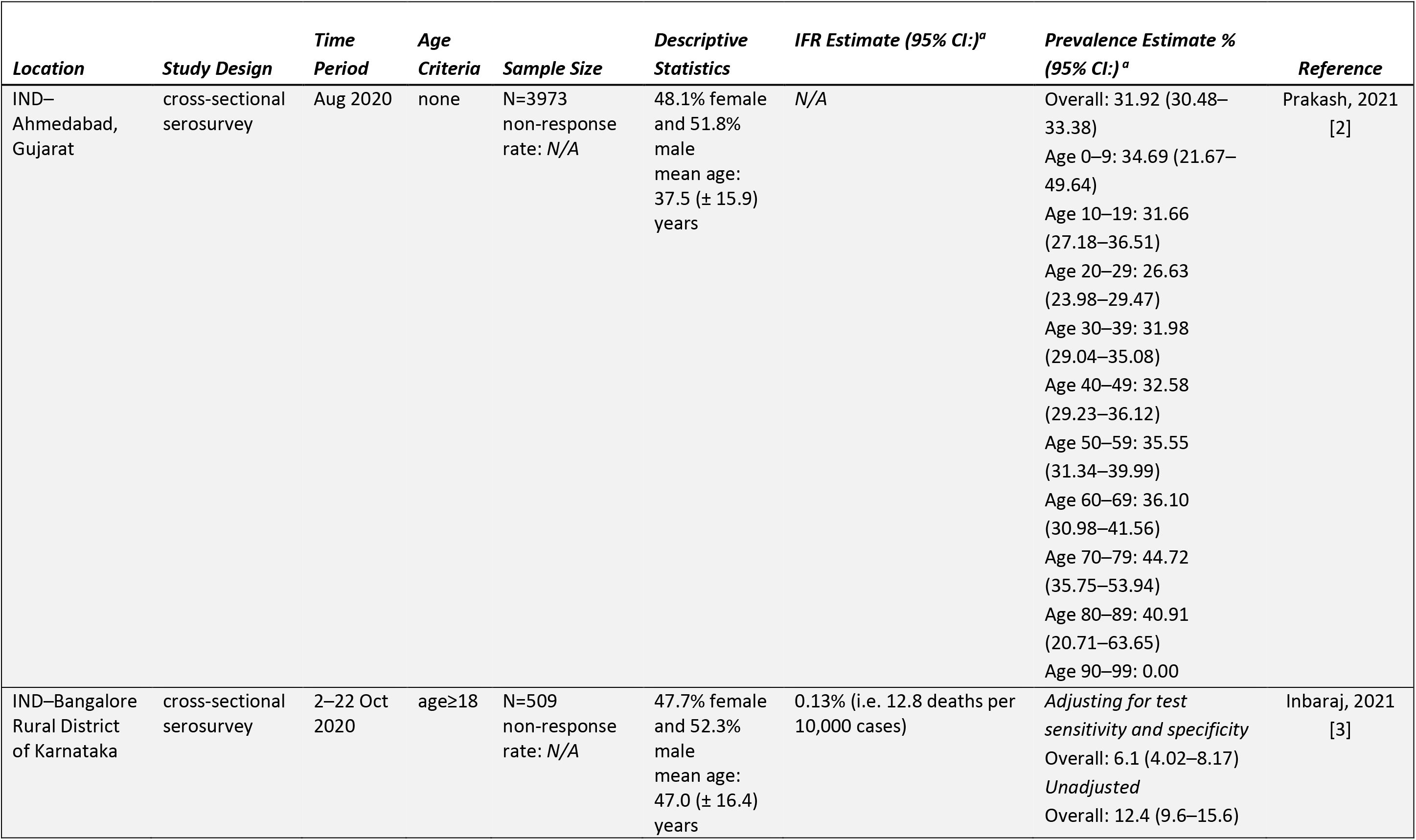
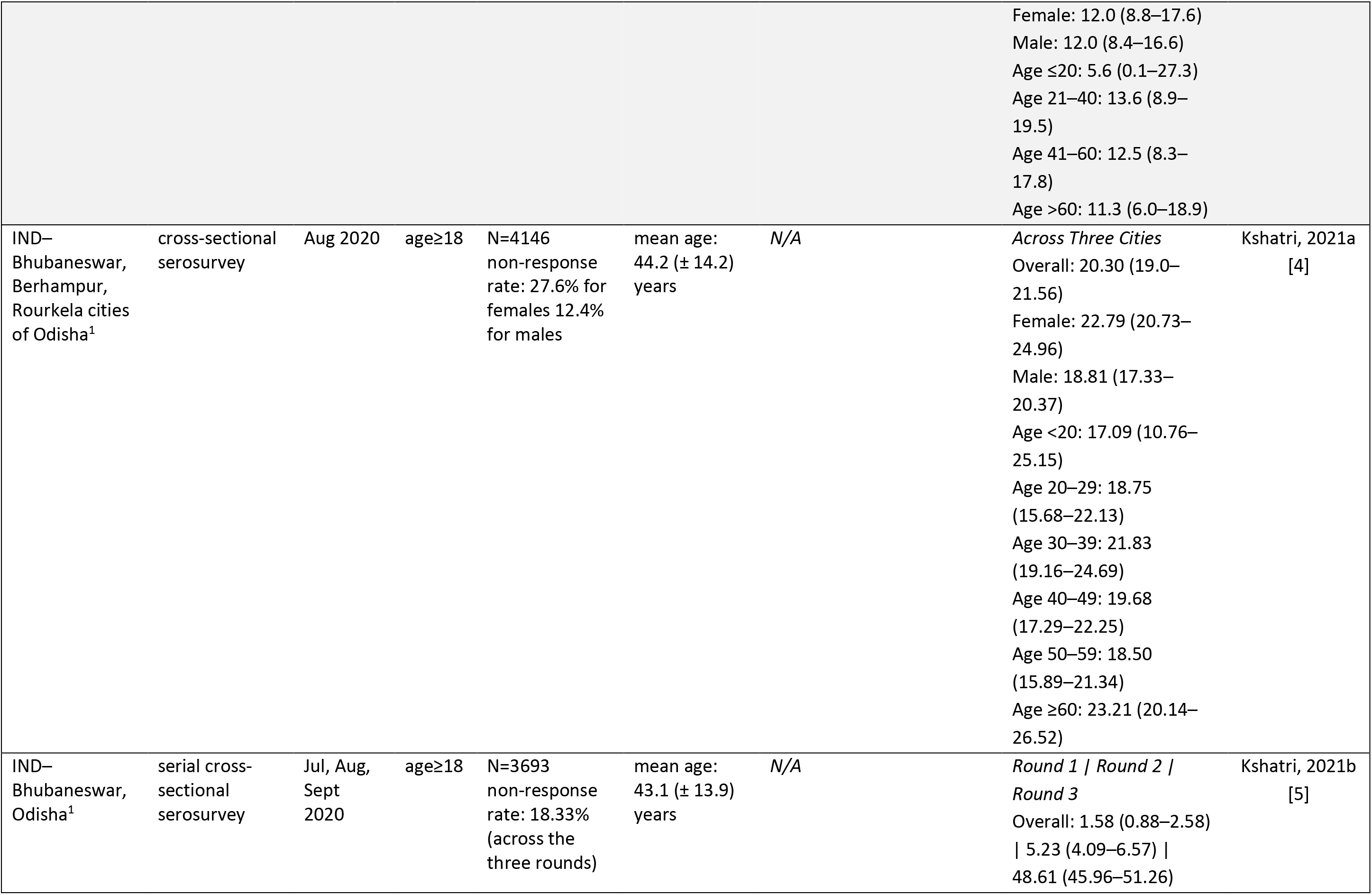
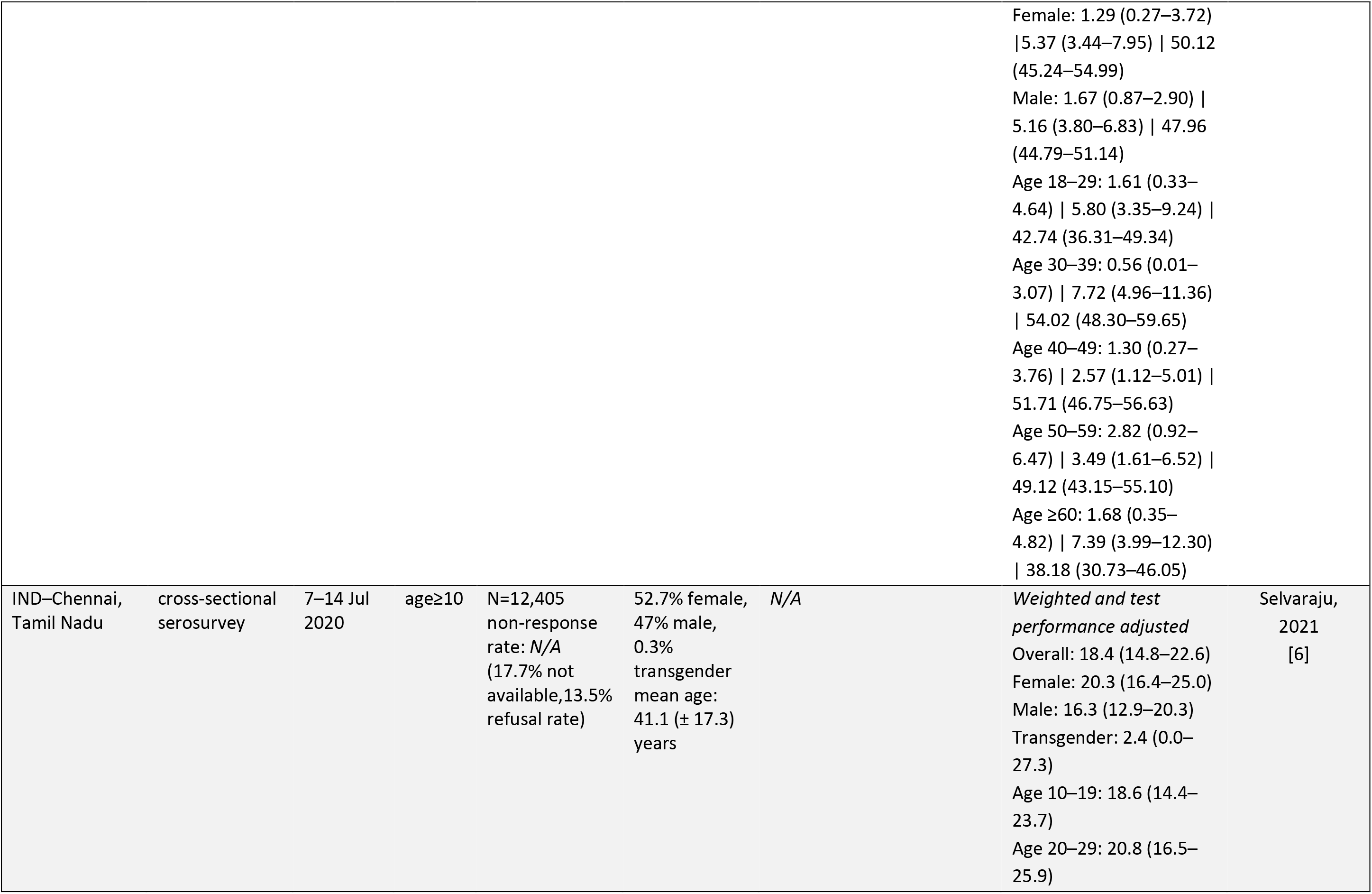
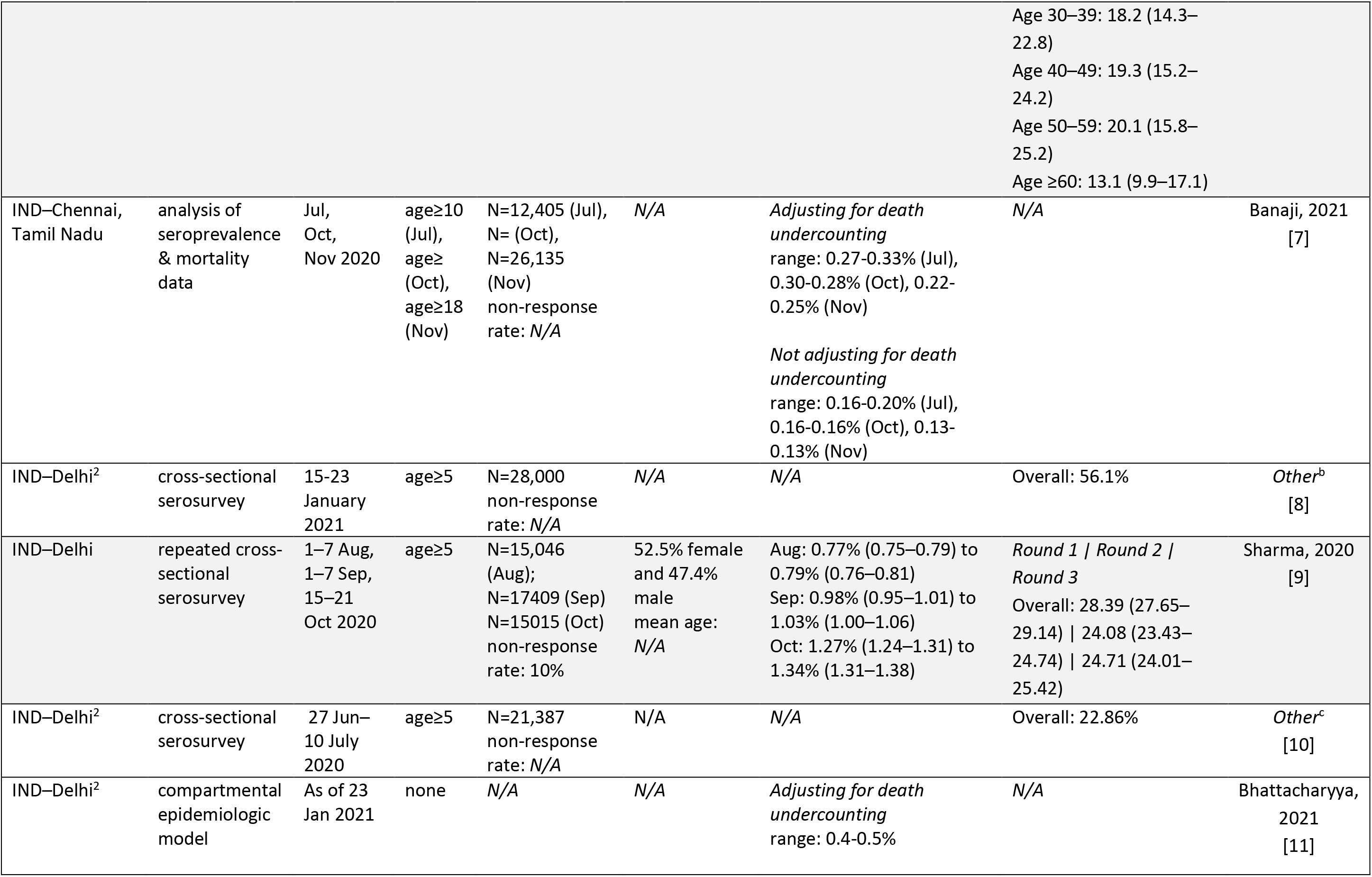
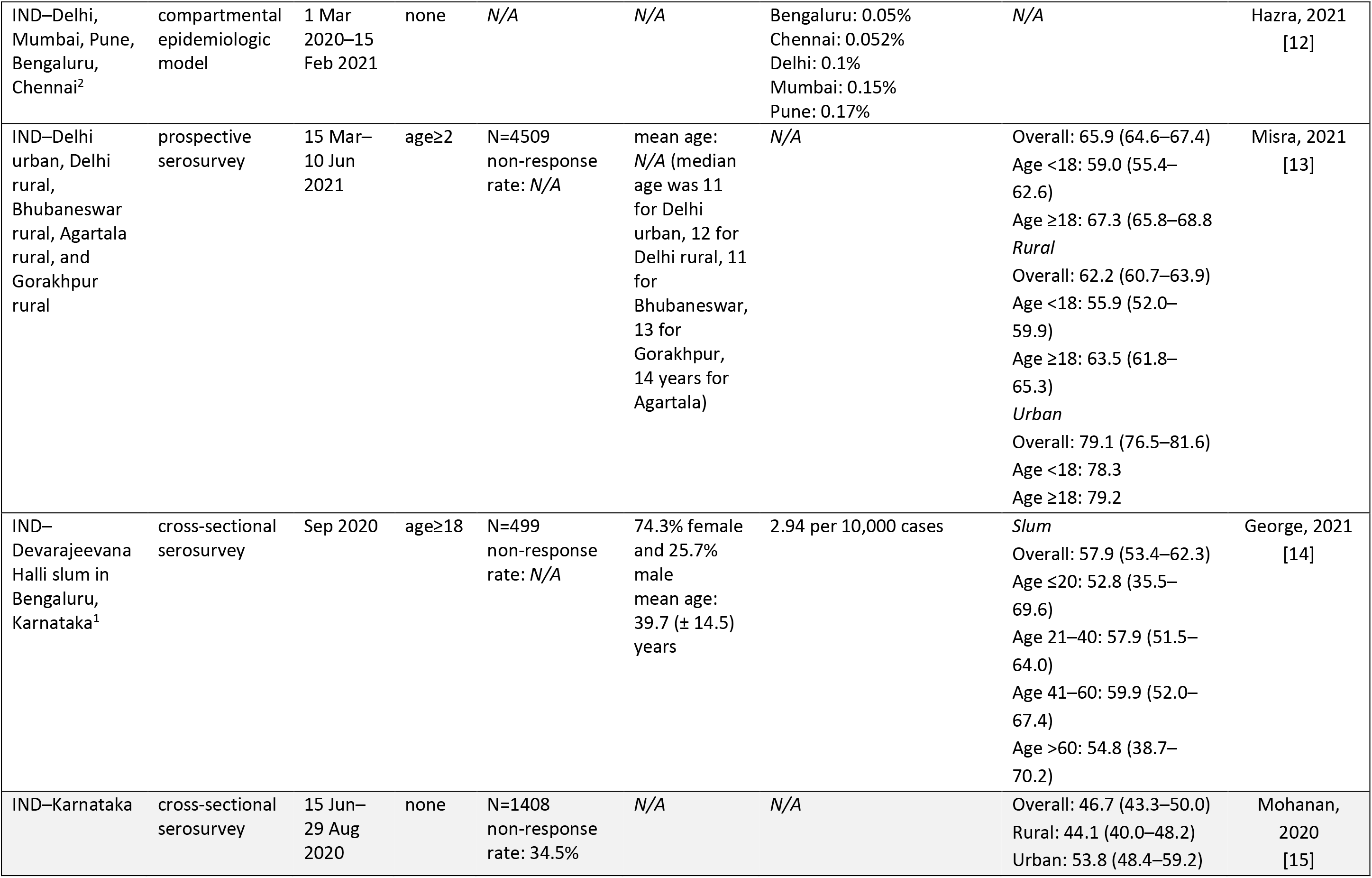
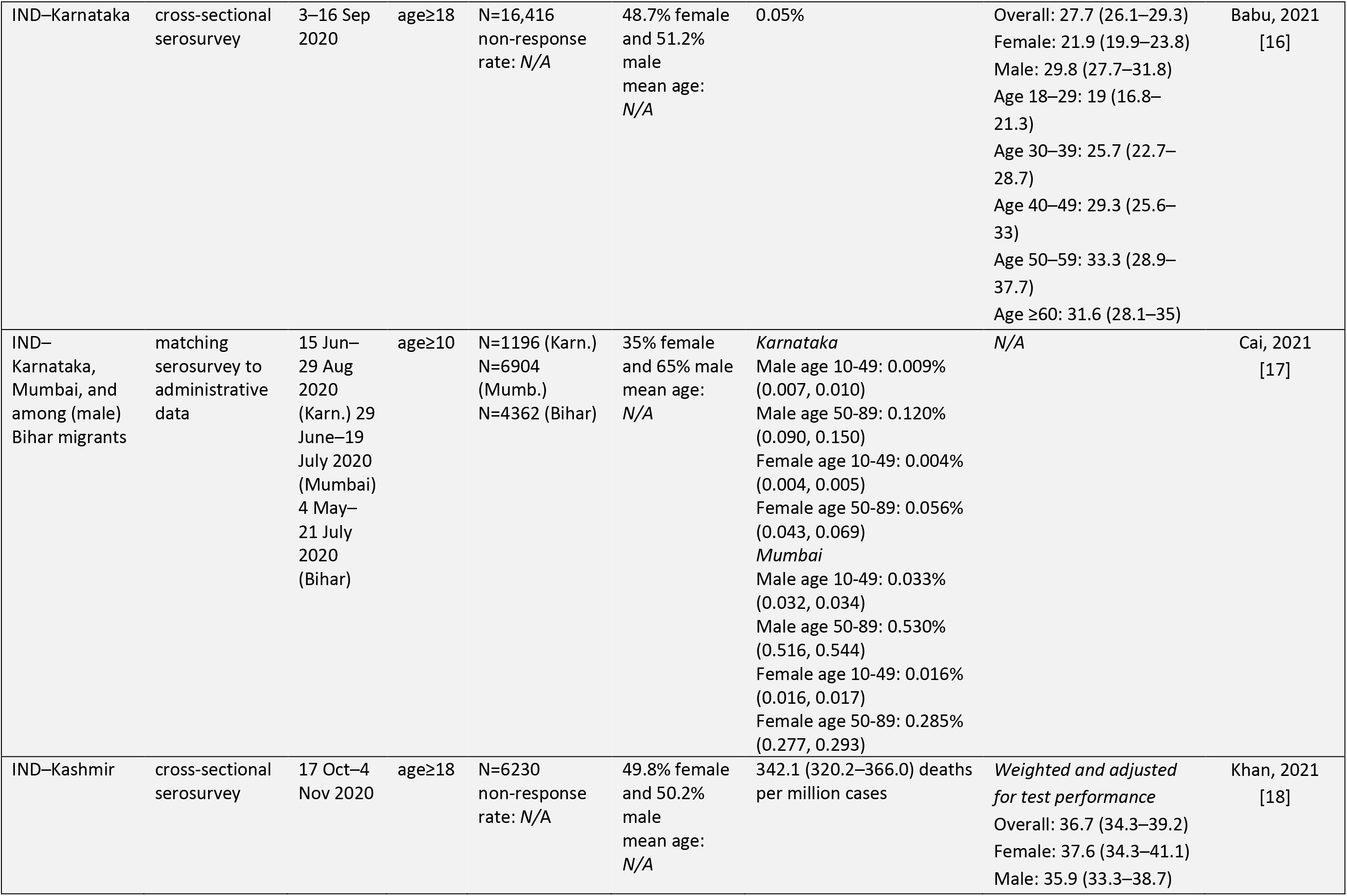
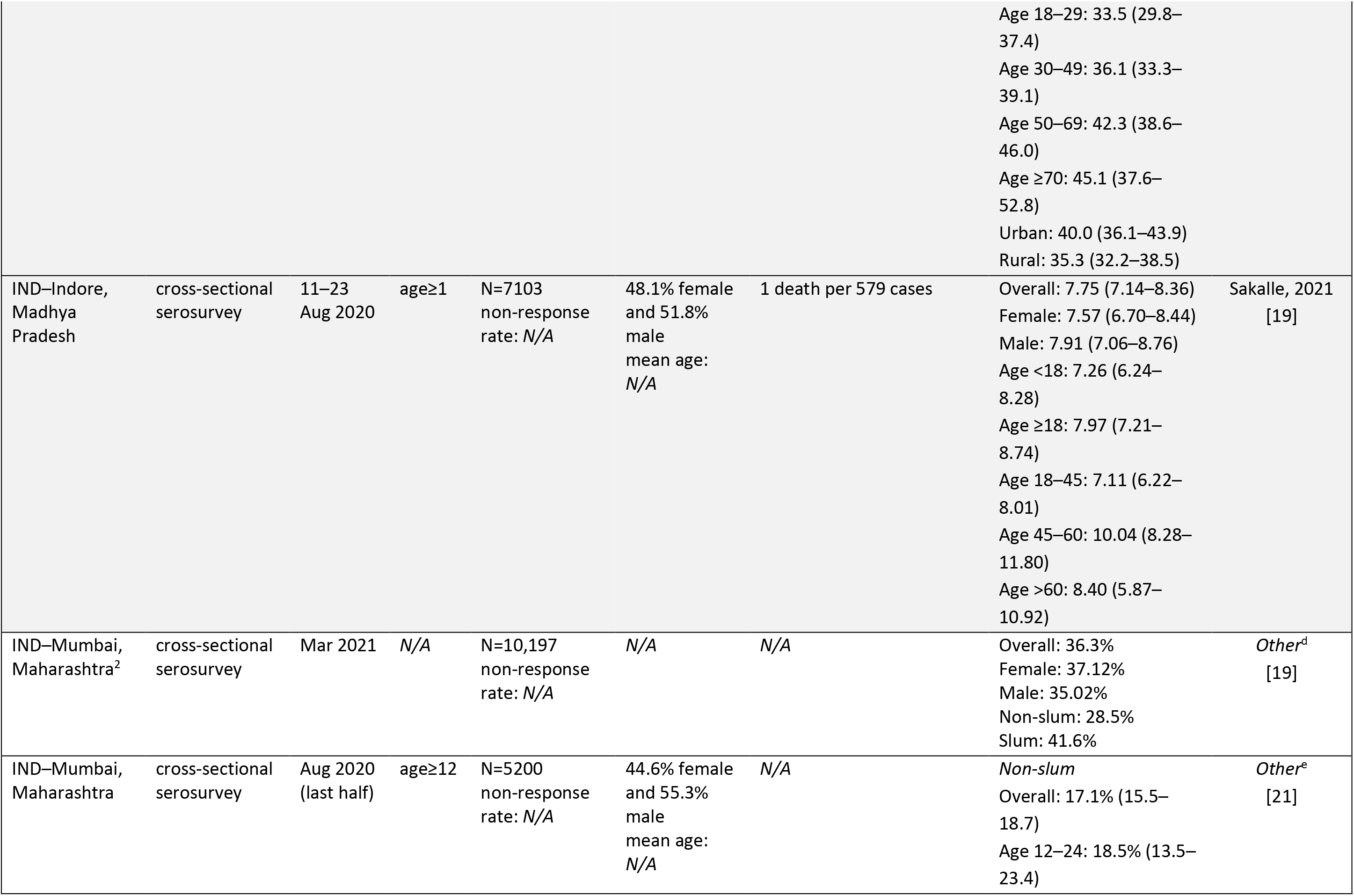
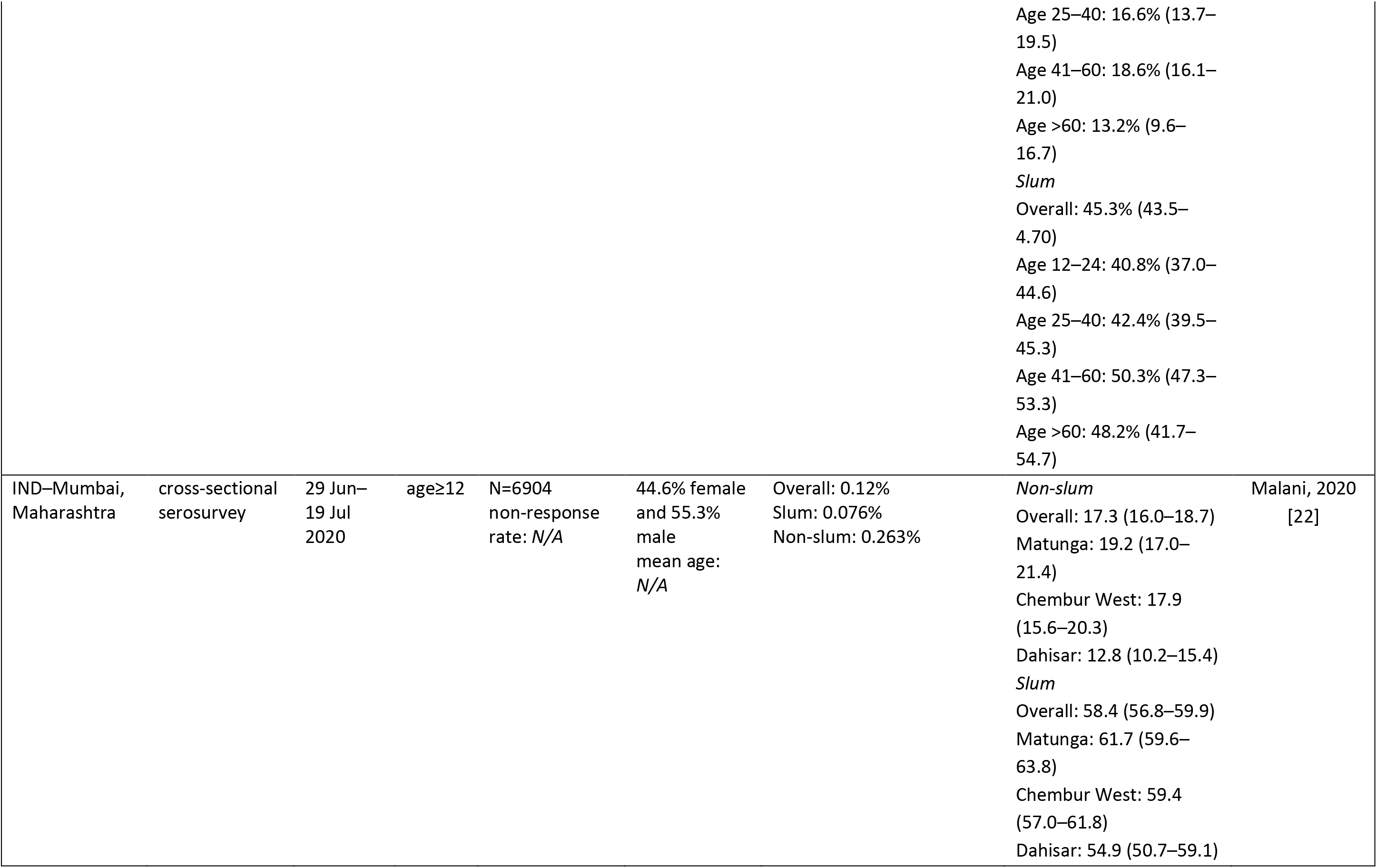
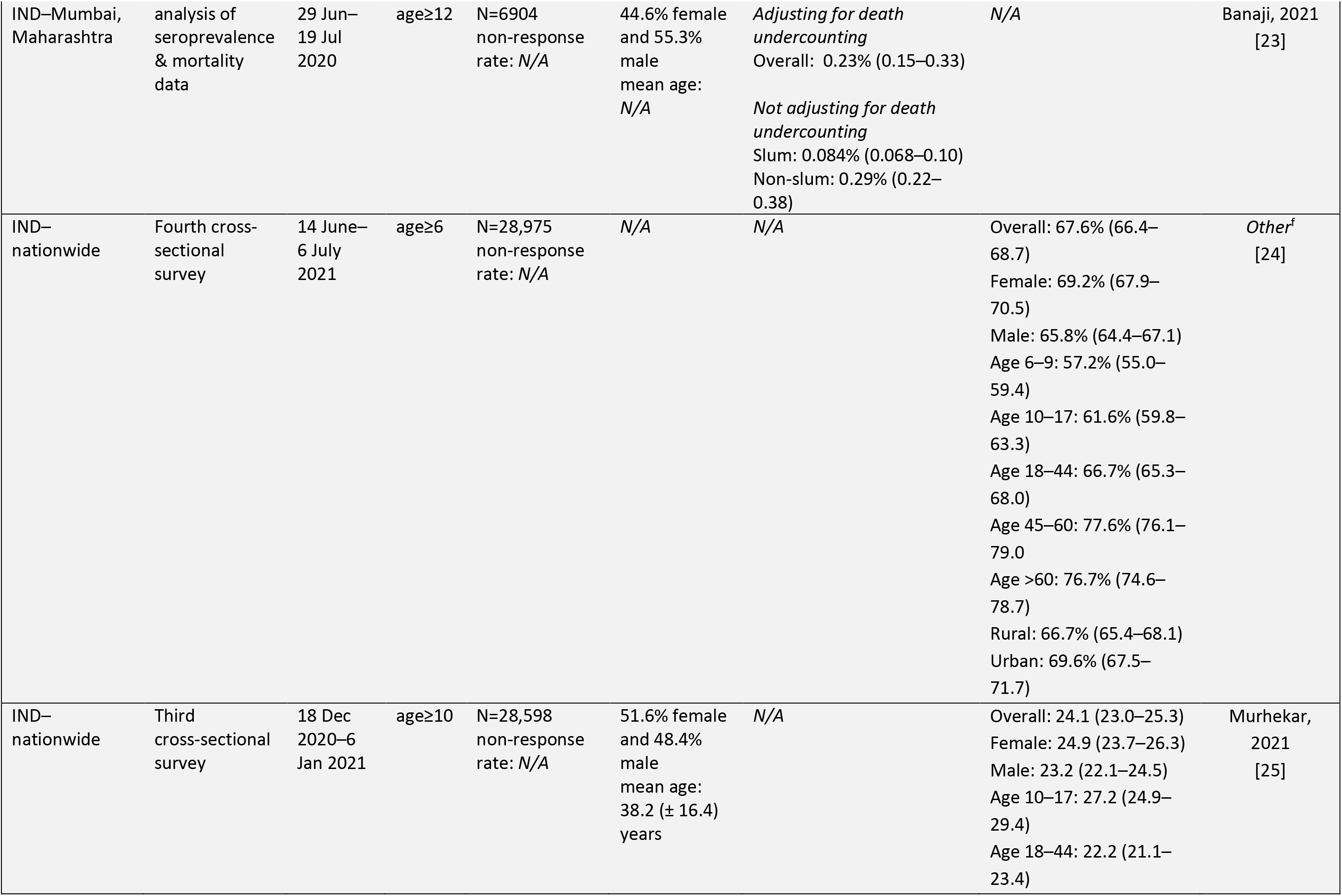
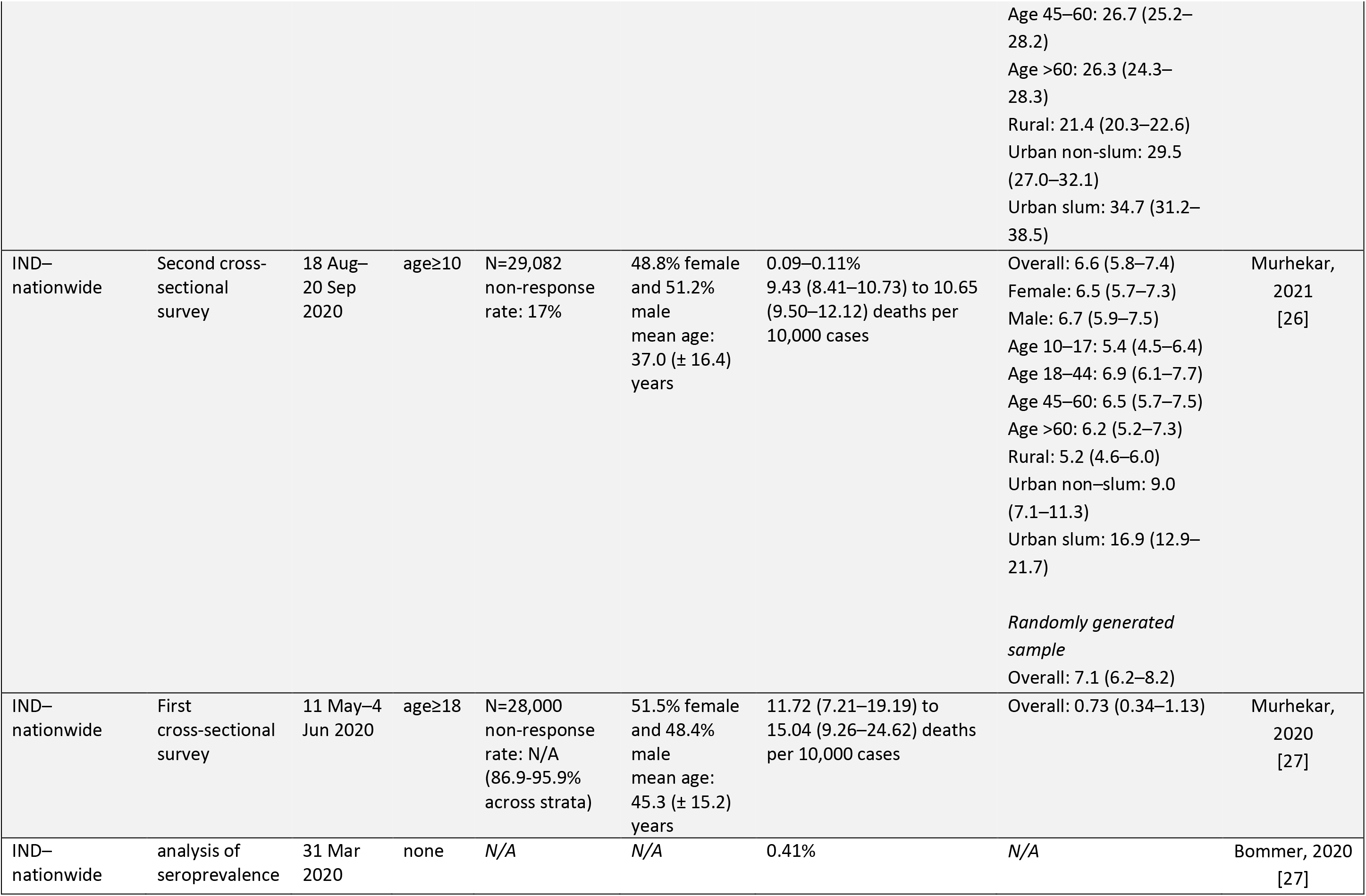
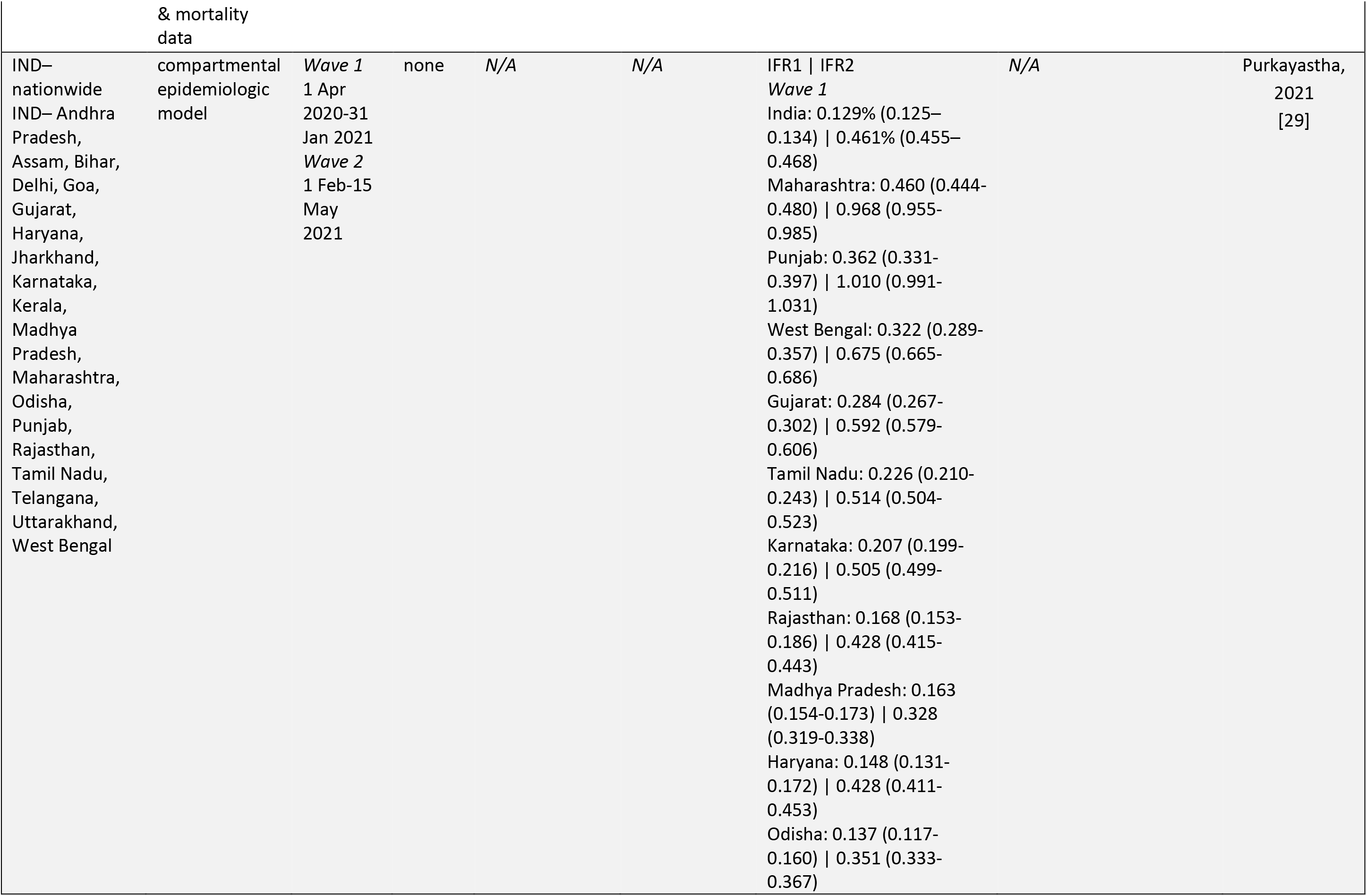
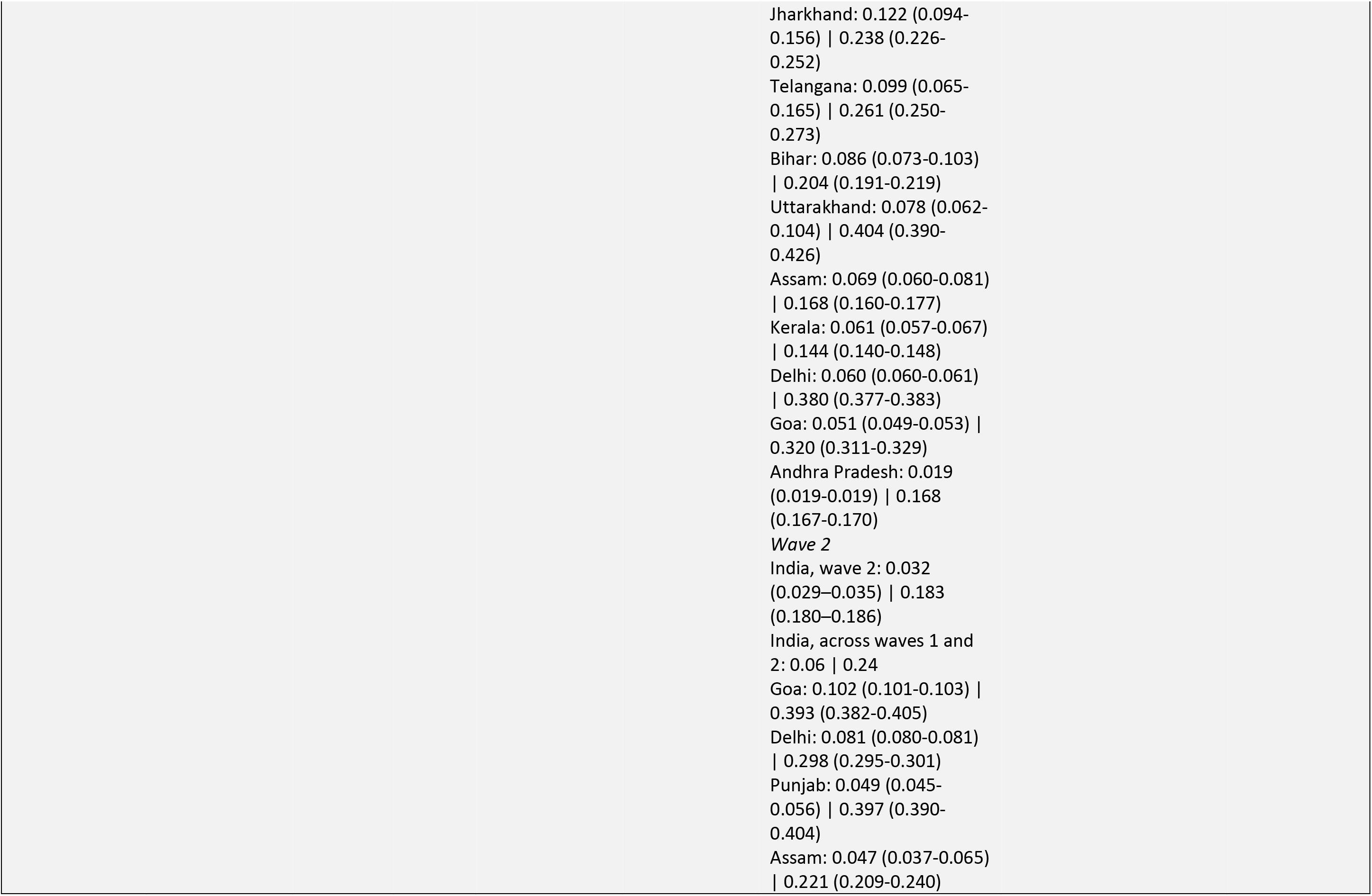
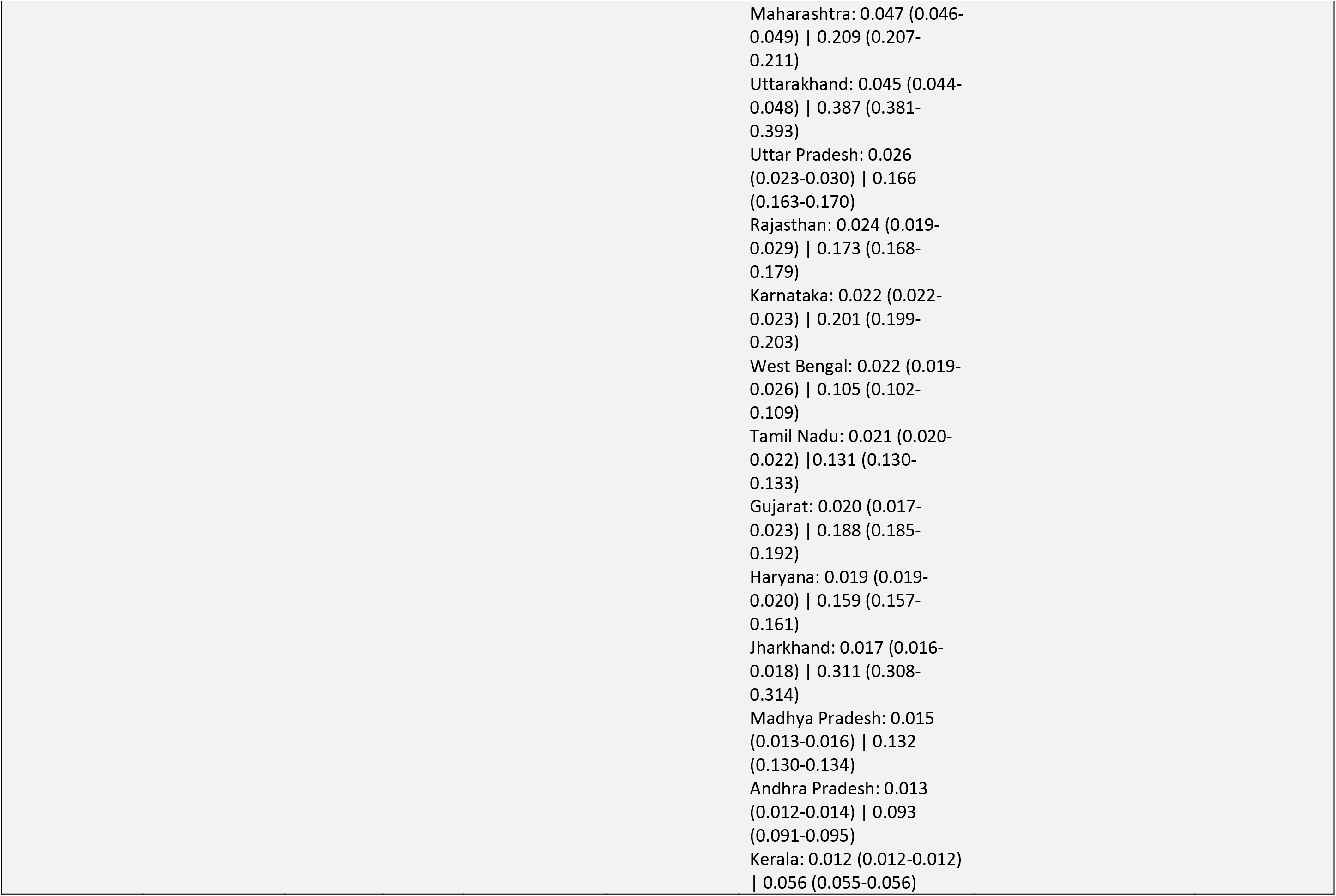
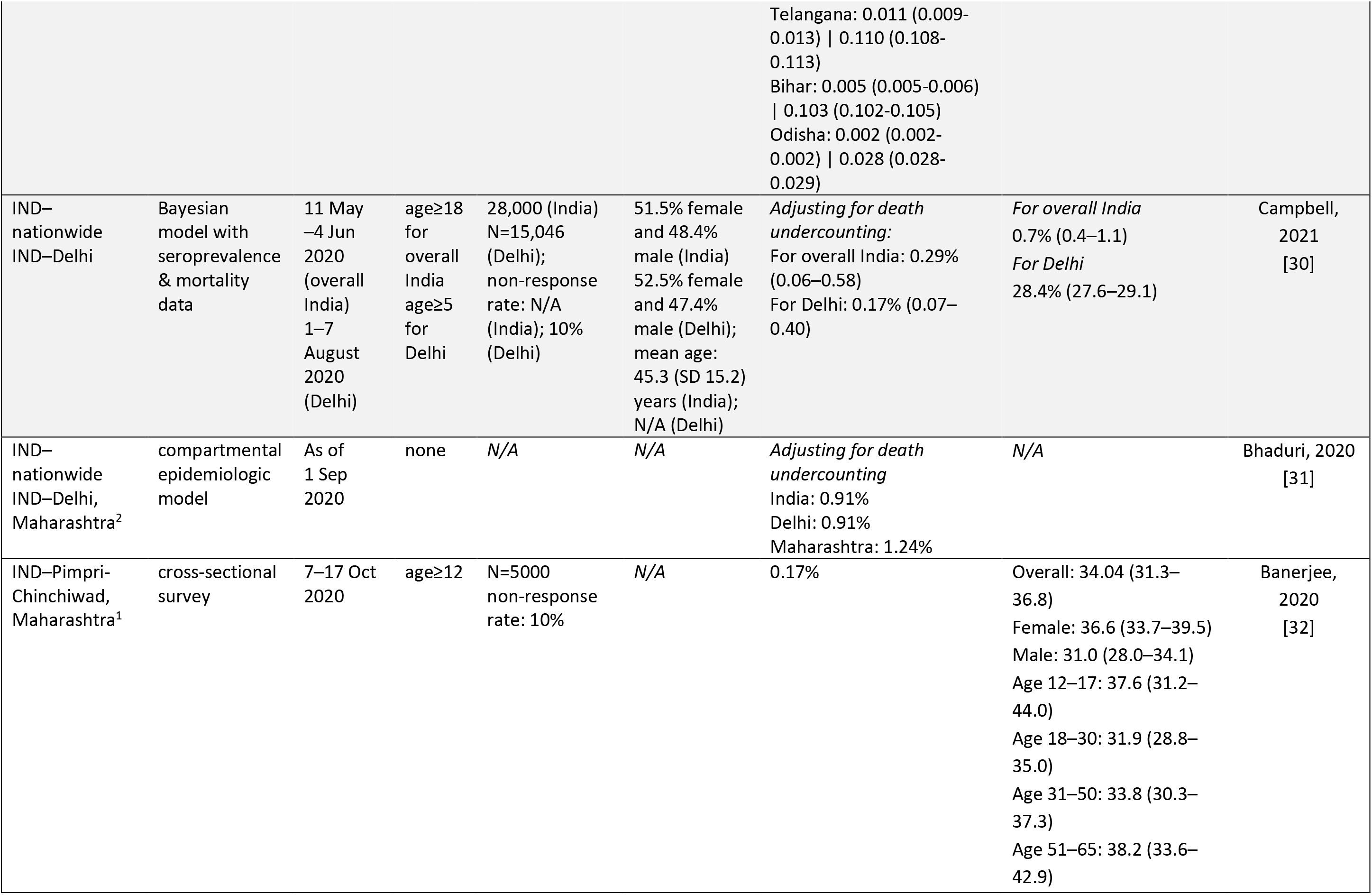
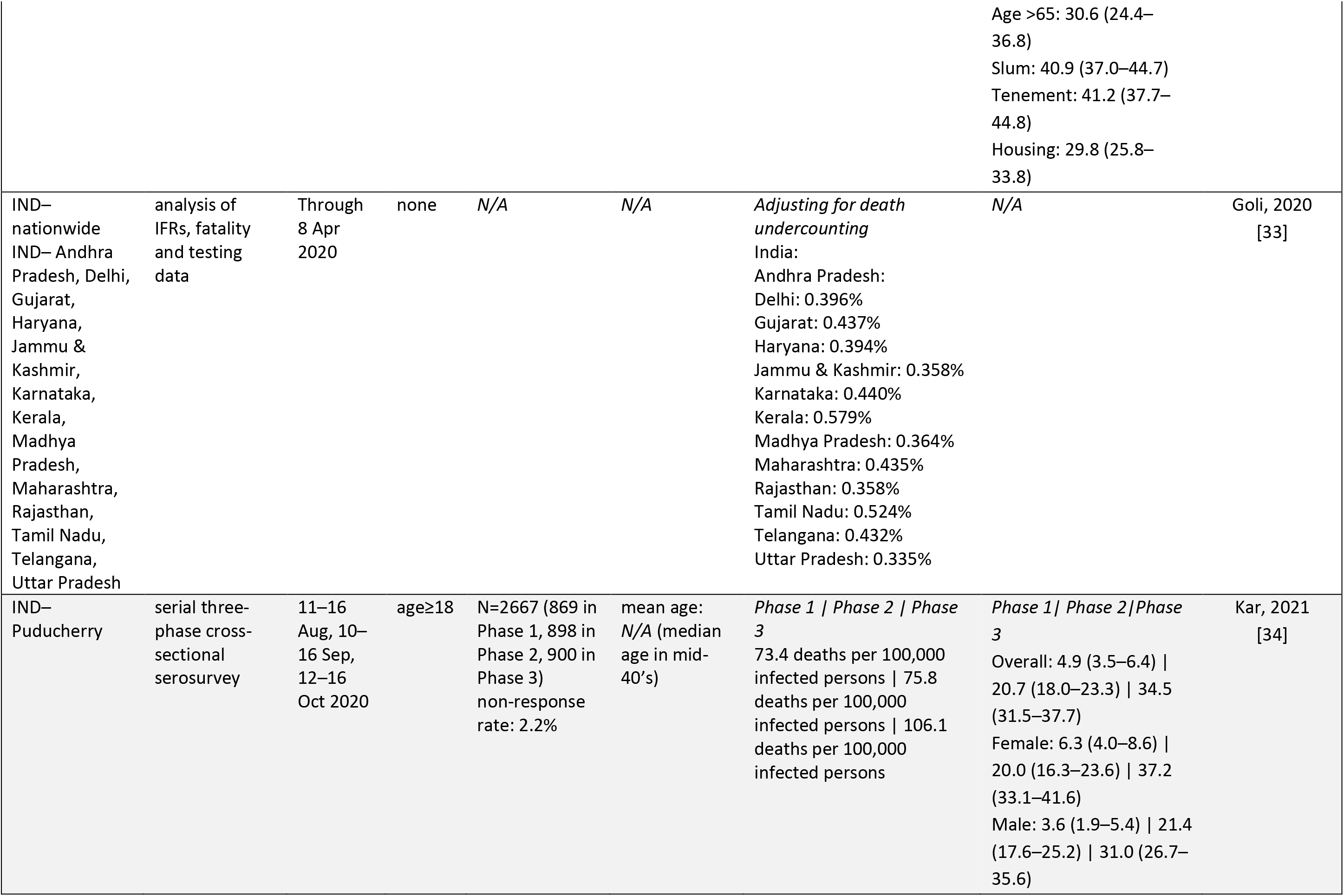
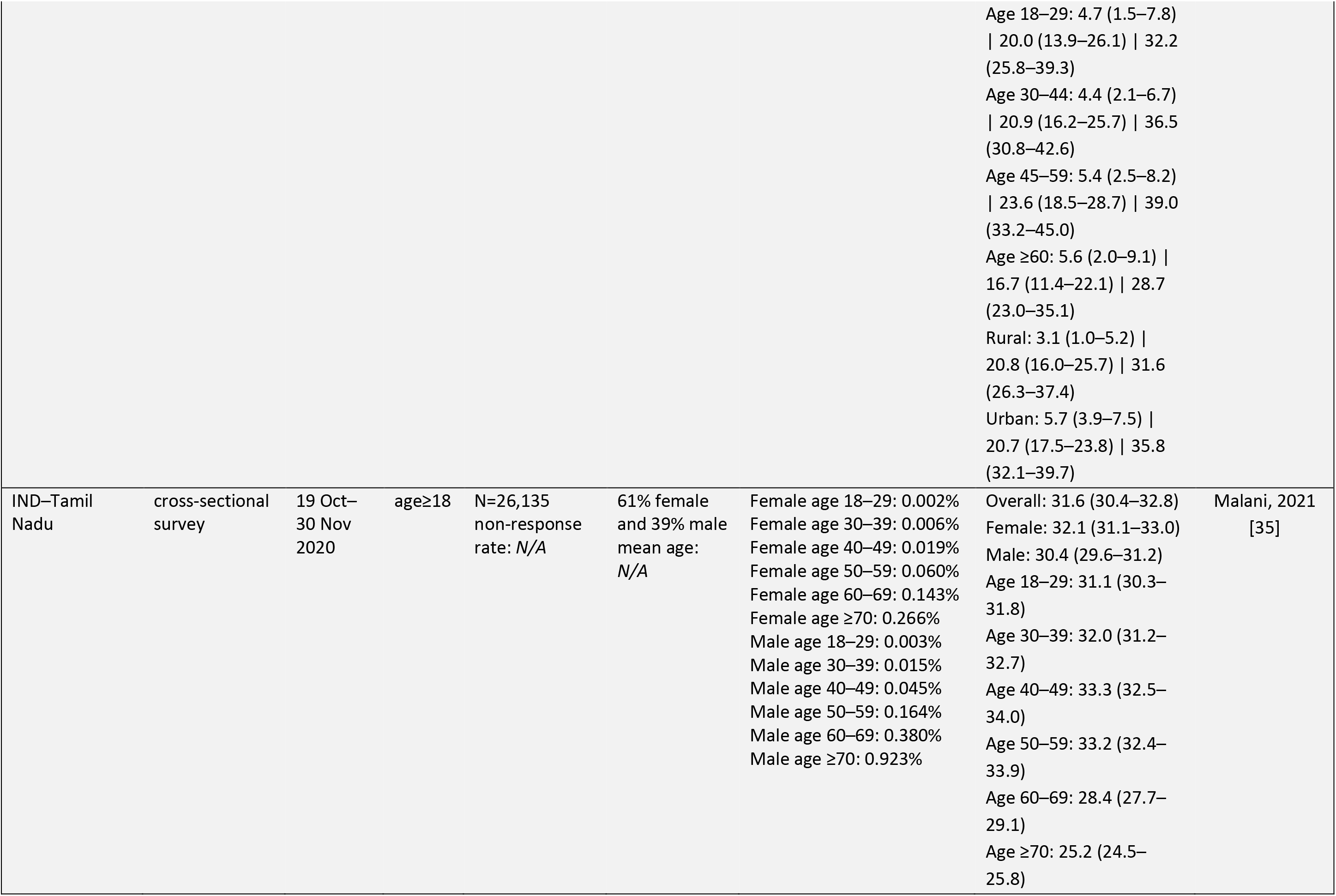
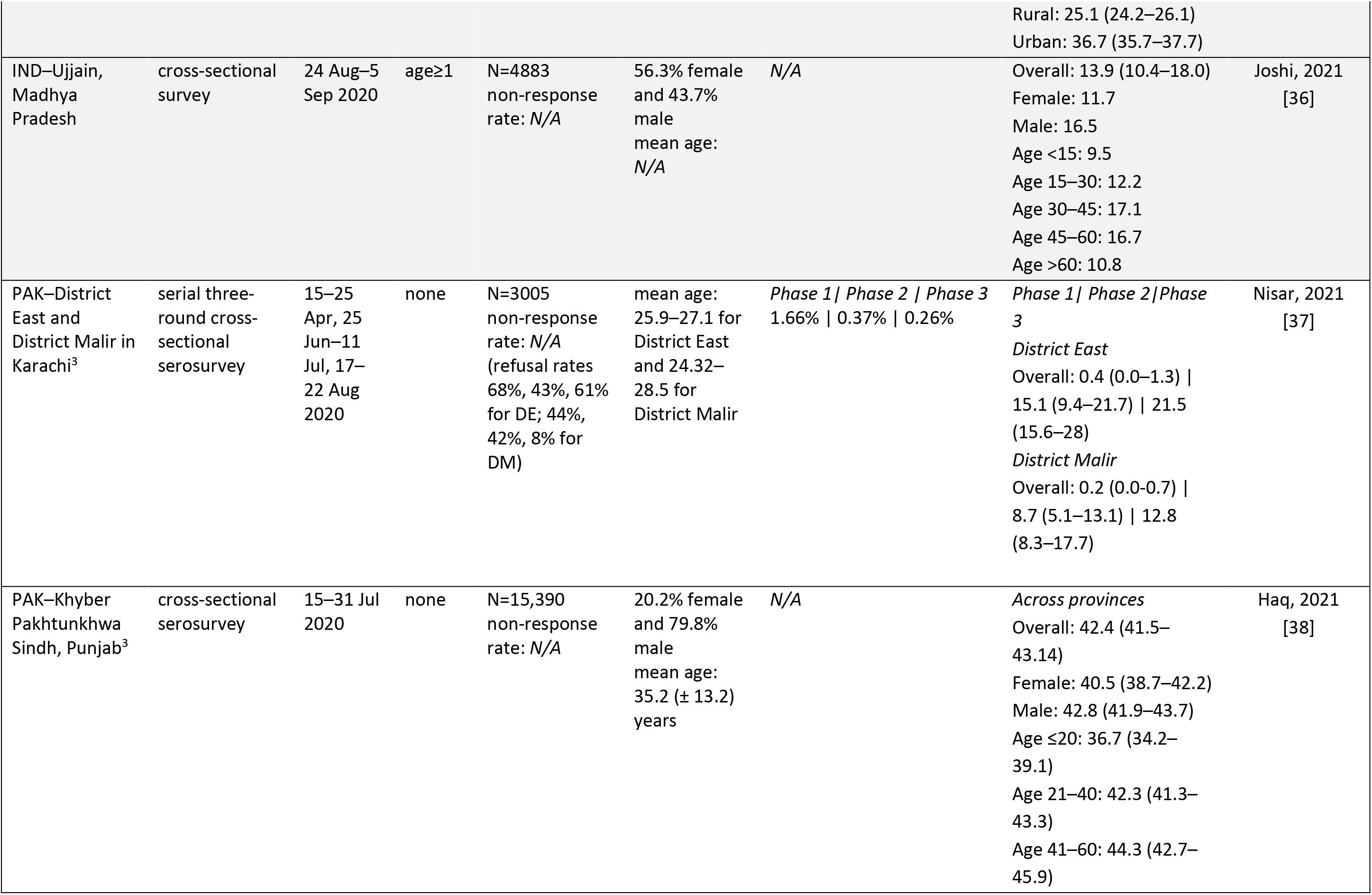
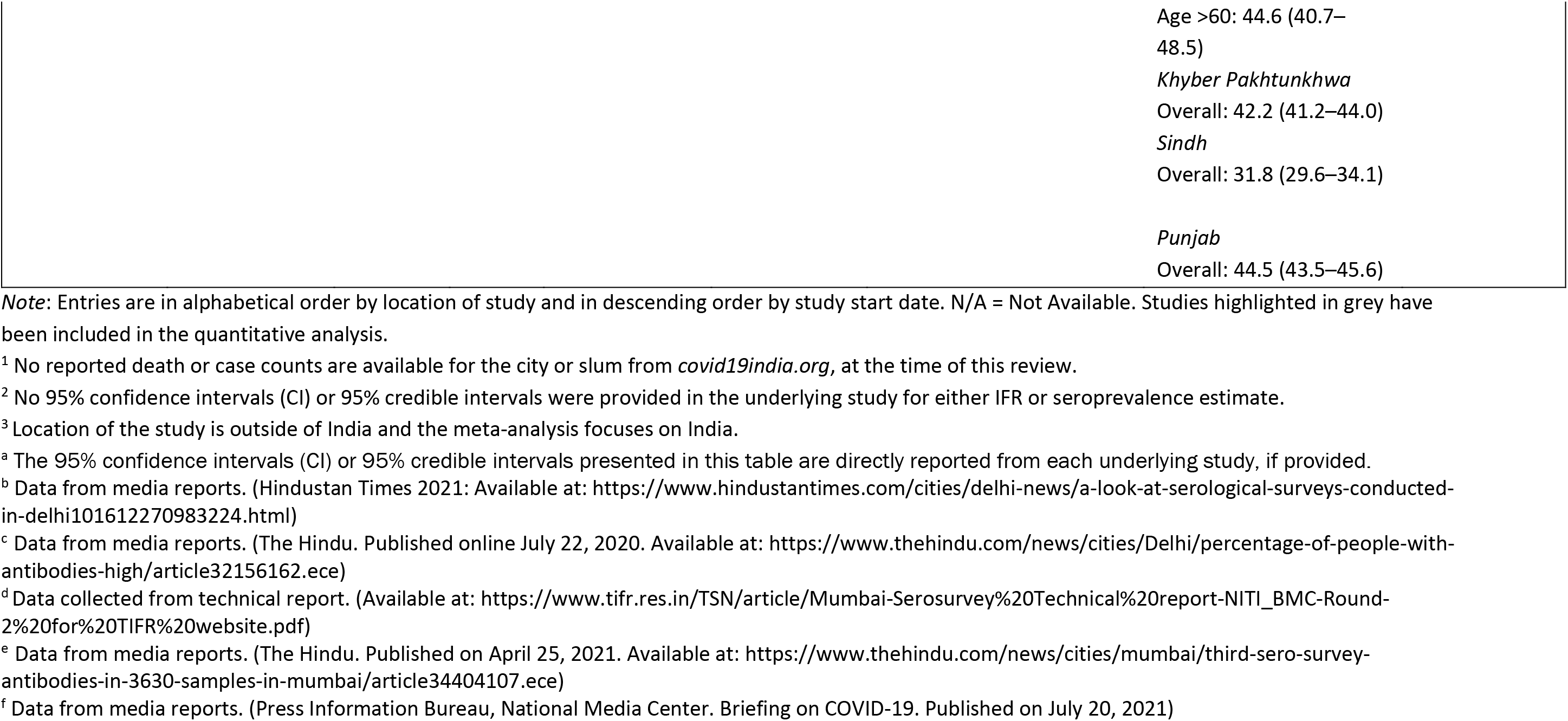
Summary of data abstraction from included studies.

## Appendix D. List of excluded articles

**Table 1.** List of excluded articles from qualitative review and reason for exclusion.

| First Author, Ref | Location (Country–Location) | Reason for Exclusion |
| --- | --- | --- |
| Ahamad, [39] | Bangladesh | Provides no relevant seroprevalence or IFR |
| Ahamad, [40] | Bangladesh | Provides no relevant seroprevalence or IFR |
| Al-Bari, [41] | Bangladesh | Provides no relevant seroprevalence or IFR |
| Barnwal, [42] | Bangladesh | Provides no relevant seroprevalence or IFR |
| Dey, [43] | Bangladesh | Provides no relevant seroprevalence or IFR |
| Hasan, [44] | Bangladesh | Active recruitment of participants |
| Islam, [45] | Bangladesh | Provides no relevant seroprevalence or IFR |
| Islam, [46] | Bangladesh | Provides no relevant seroprevalence or IFR |
| Khan, [47] | Bangladesh | Provides no relevant seroprevalence or IFR |
| Mukaddes, [48] | Bangladesh | Provides no relevant seroprevalence or IFR |
| Rahman, [49] | Bangladesh | Provides no relevant seroprevalence or IFR |
| Siam, [50] | Bangladesh | Provides no relevant seroprevalence or IFR |
| Russell, [51] | Bangladesh and others | Provides forecasted estimates |
| Rana, [52] | Bangladesh–Southern Bangladesh | Focuses on asymptomatic individuals |
| Adapa, [53] | India | Provides no relevant seroprevalence or IFR |
| Adapa, [54] | India | Provides no relevant seroprevalence or IFR |
| Al Arydah, [55] | India | Provides no relevant seroprevalence or IFR |
| Asirvatham, [56] | India | Provides no relevant seroprevalence or IFR |
| Cai, [57] | India | Previous version |
| Chatterjee, [58] | India | Provides no relevant seroprevalence or IFR |
| Chatterjee, [59] | India | Provides forecasted estimates |
| Frost, [60] | India | Provides forecasted estimates |
| Gonzalez, [61] | India | Provides no relevant seroprevalence or IFR |
| Gupta, [62] | India | Focuses on tested individuals |
| Gupta, [63] | India | Provides no relevant seroprevalence or IFR |
| Jahan, [64] | India | Provides no relevant seroprevalence or IFR |
| Kumar, [65] | India | Provides no relevant seroprevalence or IFR |
| Kumar, [66] | India | Provides no relevant seroprevalence or IFR |
| Menon, [67] | India | Provides no relevant seroprevalence or IFR |
| Mohanty, [68] | India | Provides no relevant seroprevalence or IFR |
| Mukhopadhyay, [69] | India | Provides no relevant seroprevalence or IFR |
| Naushin, [70] | India | Focuses on laboratory workers |
| Neve, [71] | India | Provides forecasted estimates |
| Parai, [72] | India | Focuses on healthcare workers |
| Radha, [73] | India | Provides no relevant seroprevalence or IFR |
| Ranjan, [74] | India | Provides no relevant seroprevalence or IFR |
| Jayesh, [75] | India | Provides no relevant seroprevalence or IFR |
| Singh, [76] | India | Focuses on high contact workers |
| Srivastav, [77] | India | Provides no relevant seroprevalence or IFR |
| Tamrakar, [78] | India | Provides no relevant seroprevalence or IFR |
| Unnikrishnan, [79] | India | Provides no relevant seroprevalence or IFR |
| Venkatesan, [80] | India | Conducted at a hospital/healthcare clinic/ICU |
| Wang, [81] | India | Provides forecasted estimates |
| Yadav, [82] | India | Focuses on patient cohort |
| Ansari, [83] | India | Provides no relevant seroprevalence or IFR |
| Abraham, [84] | India | Provides no relevant seroprevalence or IFR |
| Giri, [85] | India and hotspot regions | Provides no relevant seroprevalence or IFR |
| Shah, [86] | India and others | Provides no relevant seroprevalence or IFR |
| Wong, [87] | India and others | Examines pediatric participants |
| Zaveri, [88] | India and others | Provides no relevant seroprevalence or IFR |
| Bhattacharyya, [89] | India and states | Previous version |
| Chauhan, [90] | India and states | Provides no relevant seroprevalence or IFR |
| Meghana, [91] | India and states | Provides forecasted estimates |
| Mukherjee, [92] | India and states | Previous version |
| Purkayastha, [93] | India and states | Previous version |
| Patel, [94] | India, Bangladesh, and Pakistan | Provides no relevant seroprevalence or IFR |
| Velumani, [95] | India—12 cities across India | Focuses on tested individuals |
| Gupta, [96] | India—52 districts and 20 states across India | Focuses on SARI patient cohort |
| Prakash, [97] | India—Ahmedabad, Gujarat | Focuses on healthcare workers |
| Mahto, [98] | India—Bihar | Focuses on healthcare workers |
| Malani, [99] | India—Bihar | Focuses on working individuals |
| Madhusudan, [100] | India—Chennai, Tamil Nadu | Focuses on healthcare workers |
| Pons Salort, [101] | India—Delhi | Provides forecasted estimates |
| Siddiqui, [102] | India—Delhi | Conducted at a hospital/healthcare clinic/ICU |
| Thiruvengadam, [103] | India—Delhi | Focuses on patient cohort |
| Kaushal, [104] | India—Dharavi Slum in Mumbai, Maharashtra | Provides no relevant seroprevalence or IFR |
| Mishra, [105] | India—Eastern India | Focuses on healthcare workers |
| Kataria, [106] | India—Gurugram, Haryana | Focuses on healthcare workers |
| Ranjan, [107] | India—Karnataka | Provides no relevant seroprevalence or IFR |
| Khan, [108] | India—Kashmir | Focuses on healthcare workers |
| Kumar, [109] | India—Kerala | Focuses on healthcare workers |
| Kaur, [110] | India—Majha, Punjab | Focuses on patient cohort |
| Goenka, [111] | India—metropolitan city | Focuses on healthcare workers |
| Kumar, [112] | India—Mumbai, Maharashtra | Focuses on healthcare workers |
| Mahajan, [113] | India–Mumbai, Maharashtra | Focuses on healthcare workers |
| Mahajan, [114] | India–Mumbai, Maharashtra | Focuses on healthcare workers |
| Singhal, [115] | India–Mumbai, Maharashtra | Focuses on healthcare workers |
| Tanna, [116] | India–Nagpur, Maharashtra | Focuses on patient cohort |
| Sharma, [117] | India–New Delhi | Focuses on healthcare workers |
| Gupta, [118] | India–Northern India | Focuses on healthcare workers |
| Khan, [119, p. 2] | India–District Srinagar | Focuses on patient cohort |
| Satpati, [120] | India–Paschim Medinipur, West Bengal | Focuses on asymptomatic individuals |
| Mahto, [121] | India–Patna, Bihar | Focuses on healthcare workers |
| Bogam, [122] | India–Pune, Maharashtra | Provides no relevant seroprevalence or IFR |
| Ghose, [123] | India–Pune, Maharashtra | Focuses on asymptomatic individuals and high incidence sub-wards |
| Monteiro, [124] | India–Pune, Maharashtra | Provides no relevant seroprevalence or IFR |
| Sharma, [125] | India–Punjab | Provides no relevant seroprevalence or IFR |
| Vignesh, [126] | India–Tamil Nadu | Provides no relevant seroprevalence or IFR |
| Laxminarayan, [127] | India–Tamil Nadu and Andhra Pradesh | Provides no relevant seroprevalence or IFR |
| Panchamia, [128] | India–three states in Western India | Focuses on elderly homes |
| Suresh, [129] | India–Uttar Pradesh | Conducted at a hospital/healthcare clinic/ICU |
| Banerjee, [130] | India–West Bengal | Conducted at a hospital/healthcare clinic/ICU |
| Basnet, [131] | Nepal | Provides no relevant seroprevalence or IFR |
| Dhimal, [132] | Nepal | Provides no relevant seroprevalence or IFR |
| Pathak, [133] | Nepal | Provides no relevant seroprevalence or IFR |
| Sharma, [134] | Nepal | Conducted at a hospital/healthcare clinic/ICU |
| Abbas, [135] | Pakistan | Focuses on healthcare workers |
| Chaudhry, [136] | Pakistan | Provides no relevant seroprevalence or IFR |
| Din, [137, p. 19] | Pakistan | Provides no relevant seroprevalence or IFR |
| Peter, [138] | Pakistan | Provides forecasted estimates |
| Waqar, [139] | Pakistan | Focuses on symptomatic individuals |
| Zaidi, [140] | Pakistan | Focuses on working individuals |
| Naiyar, [141] | Pakistan–Gujrat, Punjab | Conducted at a hospital/healthcare clinic/ICU |
| Younas, [142] | Pakistan–Karachi, Sindh | Conducted among blood donors |
| Ali, [143] | Pakistan–Khyber Pakhtunkhwa | Focuses on patient cohort |
| Haq, [144] | Pakistan–Khyber Pakhtunkhwa | Focuses on healthcare workers |
| Javed, [145] | Pakistan–multiple cities across Pakistan | Focuses on working individuals |
| Nisar, [146, p. 2] | Pakistan–Peshawar | Conducted among blood donors |
| Jeewandara, [147] | Sri Lanka–Colombo, Western Province | Focuses on asymptomatic individuals |

## Appendix E. Summary of excluded articles

**Table 1.** Summary of excluded articles by country, type of study, and reason

| Exclusion Reason<sup>a</sup> | Type of Study | Number of Excluded Studies |  |  |  |  |
| --- | --- | --- | --- | --- | --- | --- |
|  |  | India | Bangladesh | Nepal | Pakistan | Sri Lanka |
| Hospital, urgent or tertiary care | Serosurvey | 6 | -- | -- | 2 | -- |
|  | Other | 6 | -- | 1 | -- | -- |
| Healthcare workers | Serosurvey | 16 | -- | -- | 2 | -- |
|  | Other | -- | -- | -- | -- | -- |
| Workers<sup>b</sup> | Serosurvey | 1 | -- | -- | 2 | -- |
|  | Other | 1 | -- | -- | -- | -- |
| Asymptomatic or Symptomatic | Serosurvey | 2 | 1 | -- | 1 | 1 |
|  | Other | -- | -- | -- | -- | -- |
| Blood donors | Serosurvey | -- | -- | -- | 2 | -- |
|  | Other | -- | -- | -- | -- | -- |
| Testing center | Serosurvey | 1 | -- | -- | -- | -- |
|  | Other | 1 | -- | -- | -- | -- |
| Active recruitment | Serosurvey | -- | -- | -- | -- | -- |
|  | Other | -- | 1 | -- | -- | -- |
| Elderly care persons | Serosurvey | -- | -- | -- | -- | -- |
|  | Other | 1 | -- | -- | -- | -- |
| No relevant measures | Serosurvey | -- | -- | -- | -- | -- |
|  | Other | 43 | 13 | 3 | 4 | -- |
| **Total Excluded Studies<sup>c</sup>:** |  | **78** | **15** | **4** | **13** | **1** |

(a) Studies excluded in the full-text screening are included in this table.
(b) Study population is limited to one or more working occupations (e.g., industrial workers, street vendors, industrial workers and street vendors, etc.).
(c) The summation of *Total Excluded Studies* across countries does not equal 109 studies because there are studies that examine multiple of the focus countries (e.g., India and Bangladesh, etc.). Phrased otherwise, the column counts in this table are not mutually exclusive.

## Appendix F. Meta-analysis methodology

The aims of the meta-analysis are two-fold (1) estimate a nationwide IFR_1_ and IFR_2_ with lower and upper bounds based on nationwide excess deaths, and (2) estimate regional IFR_1_ and IFR_2_ with lower and upper bounds based on state/city/district-specific excess deaths.

### Data collection and preparation

A description of the data collection and preparation of the datafile for the meta-analysis is provided in the ***Methods*** section. Here we elaborate on aspects that require further explanation and clarification.

For included studies with a pre-calculated infection fatality rate, the IFR_1_ and/or IFR_2_, along with the 95% confidence interval, are directly extracted from the included study. For studies that provide a pre-calculated IFR_1_ but no IFR_2_ (i.e., do not further account for death underreporting), we compute the IFR_2_ through multiplying the numerator of the pre-calculated IFR_1_ by the appropriate range of excess deaths estimates. For studies that report a pre-calculated IFR_2_ but no IFR_1_ (i.e., do not provide preliminary infection fatality estimate without accounting for death reporting), we compute the IFR_1_, using the seroprevalence estimate quoted within the included study and following the same steps described below to compute IFR_1_.

For studies without a pre-calculated IFR_1_ and/or IFR_2_, IFR_1_ and/or IFR_2_ is computed, as given in the below formulas

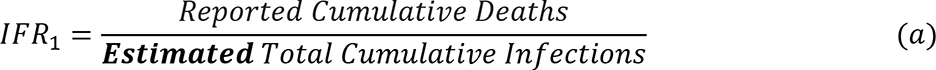

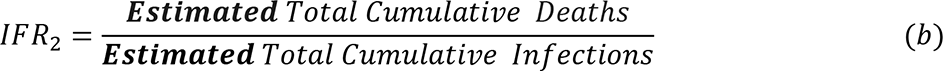

Where

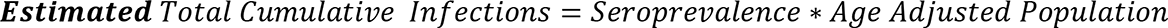

and

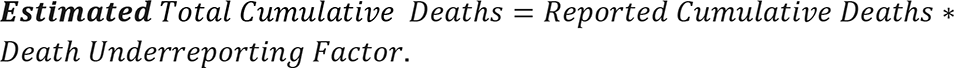

For the denominator in formulas (*a*) and (*b*), the seroprevalence estimate corresponding to the general study population, as well as the 95% confidence interval, are directly extracted from the included study. We retrieved seroprevalence estimates that were adjusted (within the serosurvey design) for the test performance and weighted to be representative of the study’s general population (typically, for the demographics age and sex, among others, such as rural versus urban), if available. For studies that did not both weight and account for test performance, we extracted the solely weighted or solely test performance adjusted seroprevalence estimate, as provided. For studies that did not report either a weighted or a test performance adjusted seroprevalence estimate, we retrieved the provided crude seroprevalence estimates. The age-adjusted population estimate is calculated as the 2019 projected population estimate on the 2011 census website multiplied by the proportion of the population above the age-cutoff of the included study (e.g., proportion of the population of Karnataka aged ≥ 18 years), as obtained from the age composition for the study area from the 2011 census. As noted in the ***Results*** section, for select cities and districts, no 2019 projected population estimate was available and as such we use the 2011 census population estimates for the following cities and districts: Ahmedabad, Chennai, Bangalore Rural District, Indore, Ujjain. We note that this may lead to an overestimation of IFR_1_ nor IFR_2_ from these studies, and in turn slightly inflated pooled estimates for the regions containing these study locations.

For the numerator in formulas (*a*) and (*b*), COVID-19 reported cumulative deaths are sourced from *<u>covid19india.org</u>* and collected 14 days after the study end date to account for delay in death from SARS-CoV-2 symptom onset. In practice, reported (i.e., observed) deaths for the target population are obtained some specified number of days after the end date of the serological study that varies between studies (e.g., from 2 days [35] to 21 days [14] [18], among others). Levin et al., 2021 [148] perform a simulation-based sensitivity analysis to derive an appropriate fatality delay, and propose and adopt in their systematic review of age-specific infection fatality rates a fatality delay of 4 weeks after the midpoint of the serosurvey. As previously discussed, reported deaths were not available for select cities or districts in *<u>covid19india.org</u>* and so we were not able to compute IFR_1_ nor IFR_2_ for the cities or districts-level studies Berhampur, Bhubaneswar, Pimpri-Chinchiwad, Rourkela and Devarajeevana Halli slum in Bengaluru, and thereby were not able to include these studies in the quantitative meta-analysis (as listed in Appendix G).

For the numerator in formula (*b*), the death under reporting factor is either directly extracted from media reports and excess deaths studies available at the time of this report (as are listed in the ***Methods*** section) or calculated using excess cumulative deaths estimates provided within these sources. For the latter, URF (D) is computed as the provided excess deaths estimate divided by the COVID-19 reported deaths 14 days after the end date of the study end date, as previously reasoned. As previously noted, excess deaths were not available for the following states at the time of this study and so we were unable to compute IFR_2_, as well as include in the regional analysis, the following states and corresponding studies: Jammu and Kashmir (ref) and Puducherry (ref).

Being that the infection fatality rate (IFR) measure is understood to be a rate and that upon inspection its distribution was heavily right skewed, a log transformation is applied to the sampling data to approximate a normal distribution.

For the 95% confidence interval for *IFR*_1_, first we obtain the standard error for seroprevalence from the directly provided 95% confidence interval from each included study as

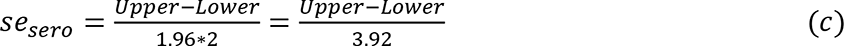

Now that we have the standard error for seroprevalence, the standard error for the log of IFR_1_ can be obtained as IFR_1_ relies on *Seroprevalence* as detailed in formula (*a*) above. First, notice that

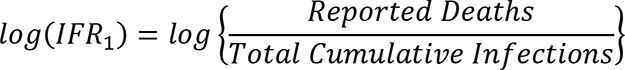

By rules of logarithmic operations, it follows that

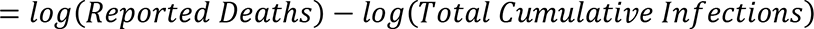

By the definition in formula (*a*) and assuming *Reported Deaths* do not contribute to variability and are thereby fixed, it follows that

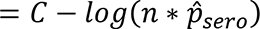

where *n* is the study sample size and 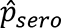 is the seroprevalence estimate from the included study.

By rules of logarithmic operations, we have that

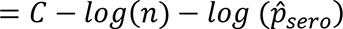

Assuming *n* does not contribute to variability and fixing at some constant

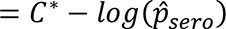

Therefore, 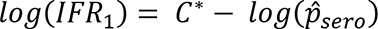. Let us consider the variance of *log (IFR_1_)*.

Substituting in for *log (IFR_1_)* from above, we have

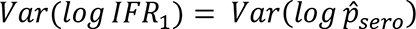

From the Taylor Series expansion, it follows that

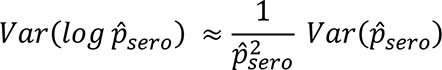

Then, the standard error for the log of IFR_1_ is given by

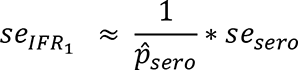

where *se_sero_* is defined as in formula (*c*) above and 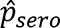 is the directly provided seroprevalence estimate.

Then, letting 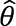 denote the estimate of *log (IFR_1_)*, the asymptotic approximate 95% confidence interval for *log (IFR_1_)* is as follows:

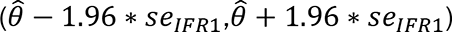

The resulting confidence intervals are then exponentiated to back-transform from the logarithmic scale.

For the 95% confidence interval for *IFR_2_*, as the upper and lower bounds of the 95% confidence interval for the associated *URF* (*D*) are often not available (i.e., the range of uncertainty associated with excess deaths estimates is not consistently available), the asymptotic approximate 95% confidence interval for *IFR_2_* is obtained by multiplying the 95% confidence interval for *IFR_1_* by the associated *UR*F (*D*).

### Meta-analysis framework

A random effects model is used with the DerSimonian-Laird (DL) estimator for *τ*^2^ (also denoted as *tau*^2^), the variance of the true effect sizes. The DL estimator, 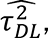 is given by

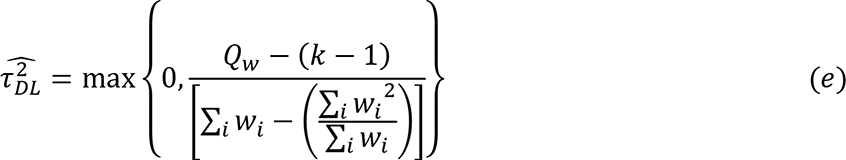

where *Q_W_* denotes the appropriate test statistic with *k-1* denoting the degrees of freedom and *w*_*i*_ denotes the sampling weight for the *C*^*t*ℎ^ included study datapoint.

The inverse variance approach is then used to obtain the pooled estimates (nationwide, regional, and state-specific within India). This means that the weighting in the random effects model is the inverse of the sampling variance, as follows

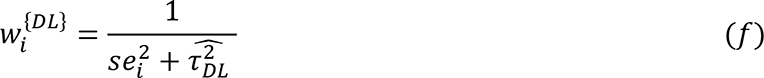

where i denotes the *i*^*t*ℎ^ included study datapoint, 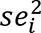 is the standard error from the *i*^*t*ℎ^ included study estimate, and 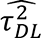 is the DL estimated random effects variance component, as defined in (*e*) above.

Using a random effects model with inverse variance method and DL estimator, the estimate for the pooled effect size is then given as follows:

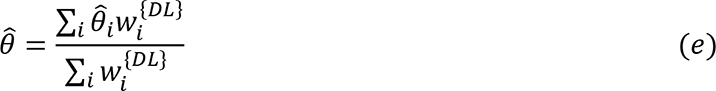

As previously mentioned, a log transformation is applied to the sampling data to approximate a normal distribution. In other words, we log transform both IFR_1_ and IFR_2_ in the meta-analysis and appropriately back-transform the resulting point estimates and standard errors by exponentiating the log-transformed values. Hence, 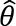 in (*e*) above in this context is 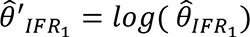 and, similarly, 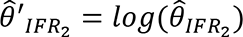.

Then, to estimate the nationwide pooled infection fatality rates (IFR_1_ and IFR_2_) for India, countrywide IFR estimates (pre-calculated or computed) among included studies (as verified through August 15, 2021) are pooled, as provided in (*e*) above using the random effects framework detailed. The nationwide pooled infection fatality estimate includes the computed IFRs from each of the four nationwide seroprevalence surveys conducted consecutively for India, as stratifying by time periods (i.e., the nationwide first and second waves of SARS-CoV-2 in India) is of particular interest.

To estimate the regional pooled infection fatality rates (IFR_1_ and IFR_2_) in India, IFR estimates (pre-calculated or computed) from included studies within a state are pooled, as provided in (*e*) above with the same random effects approach outlined. Then, the regional IFR is estimated as the pooled IFR across pooled state level IFRs, as provided in (*e*) except where *i* now denotes the *i*^*t*ℎ^ state. Since the regional analysis does not involve stratifying by waves (and thereby time points), for serial (or repeated) serosurveys for which multiple seroprevalence estimates are provided for a given study location at various time points, the most recent estimate is considered and included in the regional analysis.

Using the *meta* package in R, pooled effect sizes are estimated, as well as 95% confidence intervals, following the methodological framework above.

## Appendix G. Summary of excluded articles from quantitative summary

**Table 1.** Summary of excluded articles from quantitative summary and reason for exclusion.

| Reference | Location (Country–Location) | Reason for Exclusion |
| --- | --- | --- |
| Kshatri, 2021a<br>[4] | India–Bhubaneswar | No fatality data available |
| Kshatri, 2021b<br>[5] | India–Bhubaneswar, Berhampur, Rourkela cities of Odisha | No fatality data available |
| Banaji, 2021<br>[7] | India–Chennai, Tamil Nadu | Provides no 95% confidence intervals |
| Bhattacharyya, 2021<br>[11] | India–Delhi | Provides no 95% confidence intervals |
| *Other*<br>[8] | India–Delhi | Provides no 95% confidence intervals |
| *Other*<br>[10] | India–Delhi | Provides no 95% confidence intervals |
| Hazra, 2021<br>[12] | India–Delhi, Mumbai, Pune, Bengaluru, Chennai | Provides no 95% confidence intervals |
| Misra, 2021<br>[13] | India–Delhi urban, Delhi rural, Bhubaneswar rural, Agartala rural, and Gorakhpur rural | No fatality data available |
| George, 2021<br>[14] | India–Devarajeevana Halli slum in Bengaluru, Karnataka | Provides seroprevalence in slum |
| *Other*<br>[19] | India–Mumbai, Maharashtra | Provides no 95% confidence intervals |
| *Other*<br>[21] | India–Mumbai, Maharashtra | Provides seroprevalence in slum and non-slum |
| Malani, 2020<br>[22] | India–Mumbai, Maharashtra | Provides seroprevalence in slum and non-slum |
| Bommer, 2020<br>[27] | India–nationwide | Provides no 95% confidence intervals |
| Bhaduri, 2020<br>[31] | India–nationwide, Delhi, Maharashtra | Provides no 95% confidence intervals |
| Banerjee, 2020<br>[32] | India–Pimpri-Chinchwad, Maharashtra | No fatality data available |
| Goli, 2020<br>[33] | India–nationwide, Andhra Pradesh, Delhi, Gujarat, Haryana, Jammu & Kashmir, Karnataka, Kerala, Madhya Pradesh, Maharashtra, Rajasthan, Tamil Nadu, Telangana, Uttar Pradesh | Provides no 95% confidence intervals |
| Nisar, 2021<br>[37] | Pakistan–District East and District Malir in Karachi | Focuses on location outside of India |
| Haq, 2021<br>[38] | Pakistan–Khyber Pakhtunkhwa Sindh, Punjab | Focuses on location outside of India |

## Appendix H. Summary of seroprevalence estimates within India

Below is a forest plot with the seroprevalence estimates used to compute IFR_1_ and in turn IFR_2_ (except for studies for which IFR_2_ was pre-calculated) in the meta-analysis of nationwide and regional IFRs.

**Figure 1.**
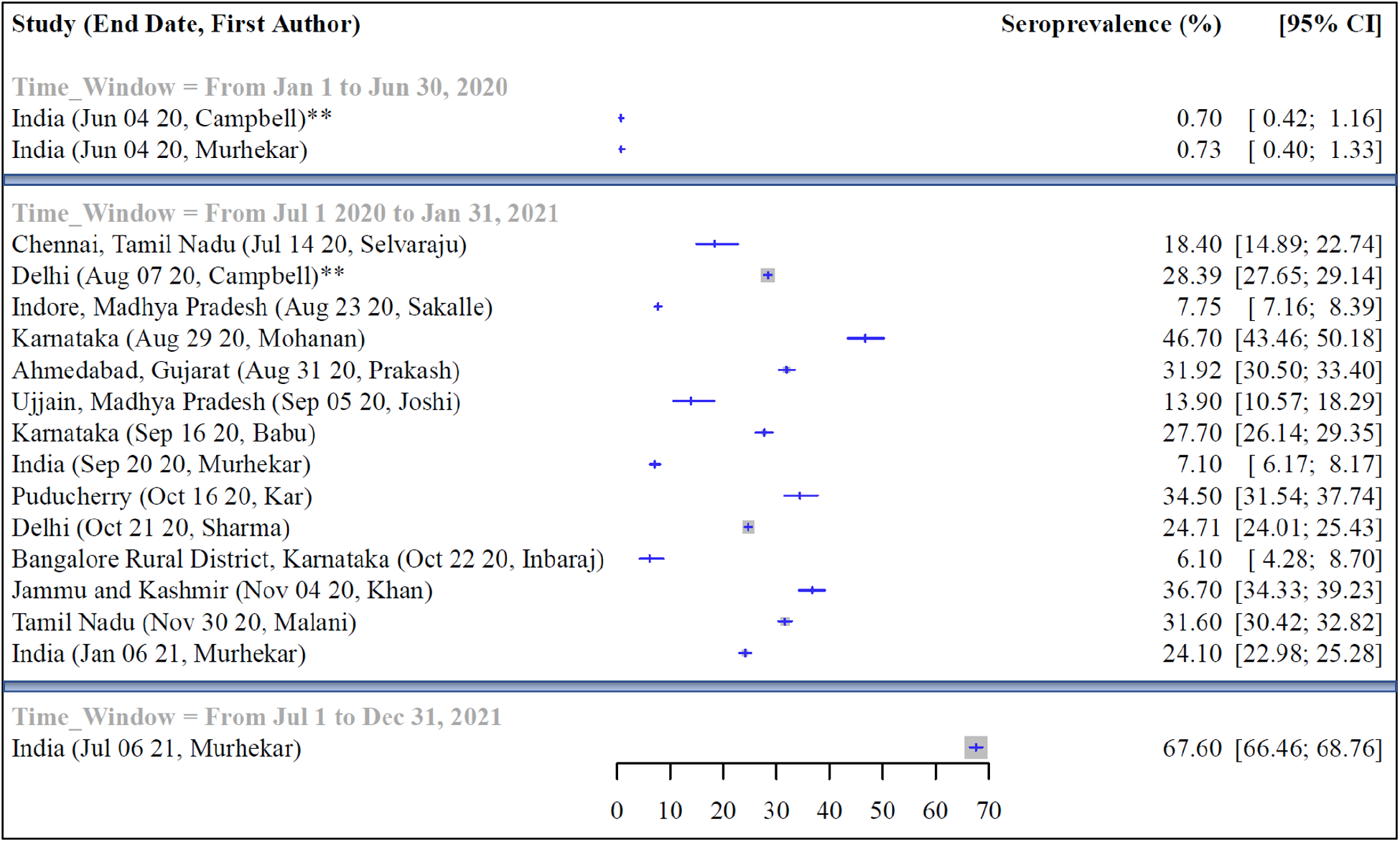
Forest plot of SARS-CoV-2 seroprevalence estimates for India utilized in meta-analysis of IFR_1_ and IFR_2_.

## Appendix I. Risk of bias assessment across included articles

See supplementary file *Supplementary_RiskofBias.xlsx* for results from the risk of bias assessment among the included studies in the meta-analysis, using the Joanna Briggs Institute (JBI) tool. Responses to each question in the JBI approach, as well as the cumulative score and rank of risk of bias, are detailed for each of the 19 studies (i.e. 15 serosurvey studies and 4 other study designs).

## Appendix J. Assessment of publication bias

To formally test for funnel plot asymmetry, the Egger’s test (i.e., linear regression) is performed with a resulting p-value of 0.0501. Seeing as the significance level of the funnel plot intercept in the Egger’s test nearly meets the benchmark of 0.05, we further conduct the Begg’s test (i.e., rank correlation test) and with a p-value<0.0001, conclude that the funnel plot is asymmetric. As discussed within the ***Results*** section, despite the results of these diagnostic tests, we do not suspect that publication bias is the driving factor behind the observed asymmetry. Firstly, this is because the bulk of the included studies are seroprevalence studies, which are inherently large studies with rigorous study designs, thereby tending toward high precision (i.e., low standard errors). Secondly, there may be heterogeneity in the true effect size between the included studies for reasons such as geographic variation that may be attributing to the largely horizontal dispersion of the standard errors in Figure 6 (the funnel plot) in the ***Results*** section, and funnel plots assume a single true effect size.

## Appendix K. SEIR-fansy model framework

*(Note: Explanation below is unchanged from Supplementary Materials in previous submission^1^.)*

### Introduction

Here we are using the SEIR-fansy model^1,2^ and software package^3^ which uses a compartmental model accounting for false negative rates and preferential diagnostic testing for SARS-CoV-2 infections. The SEIR-fansy model can be represented by the compartmental model in Figure S1.

1. Purkayastha S, Kundu R, Bhaduri R, Barker D, Kleinsasser M, Debashree R, Mukherjee B. Estimating the wave 1 and wave 2 infection fatality rates from SARS-CoV-2 in India*. BMC Res Notes*. 14, 262 (2021). doi:10.1186/s13104-021-05652-2.

2. Bhaduri R, Kundu R, Purkayastha S, Kleinsasser M, Beesley LJ, Mukherjee B. *Extending the Susceptible-Exposed-Infected-Removed (SEIR) model to handle the high false negative rate and symptom-based administration of Covid-19 diagnostic tests: SEIR-fansy.* medRxiv [Preprint]. 2020 Sep 25:2020.09.24.20200238. doi: 10.1101/2020.09.24.20200238. PMID: 32995829; PMCID: PMC7523173.

3. Ritwik Bhaduri, Ritoban Kundu, Soumik Purkayastha, Lauren Beesley and Bhramar Mukherjee (2020). *SEIRfansy: Extended Susceptible-Exposed-Infected-Recovery Model.* R package version 1.1.0. https://CRAN.R-project.org/package=SEIRfansy

**Figure S1:**
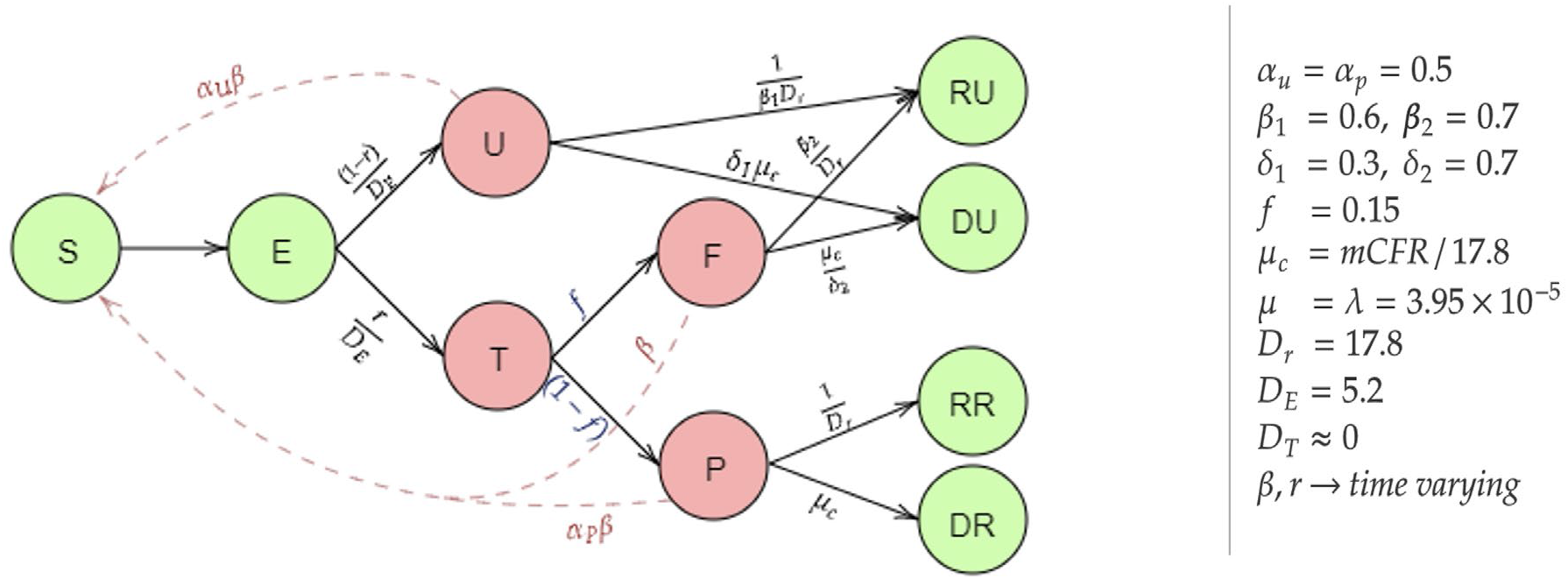
Schematic diagram for the SEIR-fansy model with imperfect testing and misclassification.

### Mathematical framework

The following differential equations summarize the transmission dynamics being modeled.

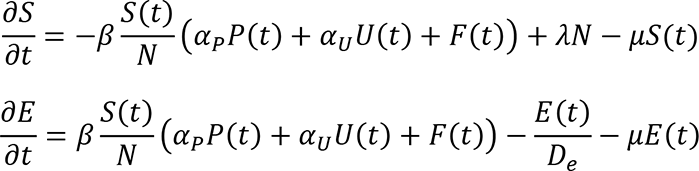

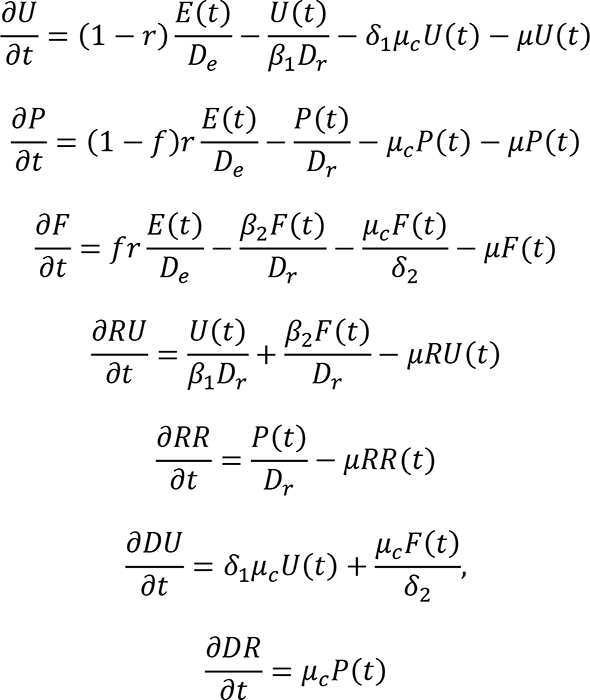

Using the Next Generation Matrix Method (28), we have calculated the basic reproduction number

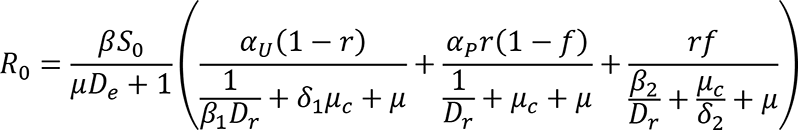

where *S*_0_ = λ/*μ* = 1 since we have assumed that natural birth and death rates are equal within this short period of time. In this setting, both *β* and *R* are time-varying parameters which are estimated using the Metropolis-Hastings MCMC method. To estimate the parameters, we at first need to solve the differential equations, which is difficult to perform in this continuous-time setting. It is also worth noting that we do not require the values of the variables for each time point. Instead, we only need their values at discrete time steps, i.e., for each day. Thus, we approximate the above set of differential equations by a set of recurrence relations. For any compartment *X*, the instantaneous rate of change with respect to time *t* (given by 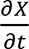) is approximated by the difference between the counts of that compartment on the (*t* + 1)^*t*ℎ^ day and the *t*^*t*ℎ^ day, that is *X*(*t* + 1) − *X*(*t*). Starting with an initial value for each of the compartments on the Day 1 and using the discrete-time recurrence relations, we can then obtain the solutions of interest. Some examples of these discrete-time recurrence relations are presented below.

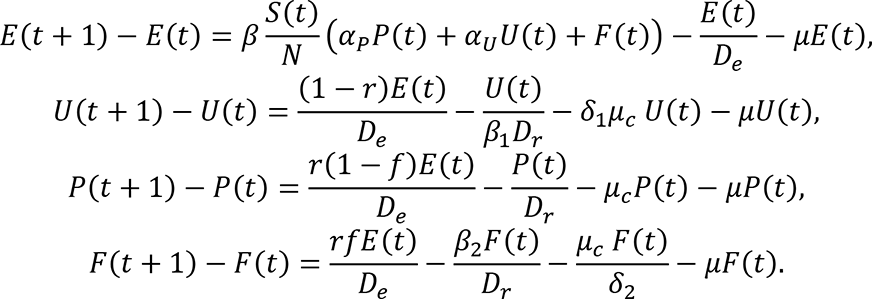

The rest of the differential equations can each be similarly approximated by a discrete-time recurrence relation.

### Likelihood assumptions and estimation

We use Bayesian estimation techniques and Markov chain Monte Carlo (MCMC) methods (namely, Metropolis-Hastings method with Gaussian proposal distribution) for estimating the parameters. First, we approximated the above set of differential equations using a discrete time approximation using daily differences. So, after we started with an initial value for each of the compartments on the day 1, using the discrete time recurrence relations we can find the counts for each of the compartments on the next days. To proceed with the MCMC-based estimation, we specify the likelihood explicitly. We assume (conditional on the parameters) the number of new confirmed cases on day *RR* depend only on the number of exposed individuals on the previous day. Specifically, we use multinomial modeling to incorporate the data on recovered and deceased cases as well. The joint conditional distribution is

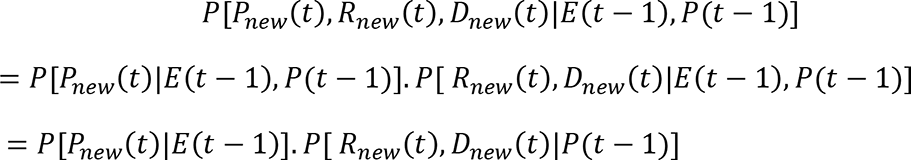

A multinomial distribution-like structure is then defined,

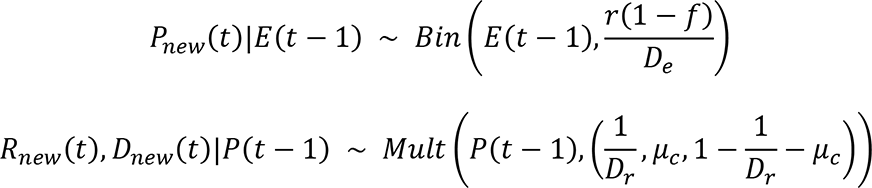

*Note:* the expected values of *E*(*t* − 1) and *P*(*t* − 1) are obtained by solving the discrete time differential equations as described earlier.

### Prior assumptions and MCMC

For the parameter *r*, we assume a *U*(0,1) prior, while for *β*, we assume an improper non-informative flat prior with the set of positive real numbers as support. After specifying the likelihood and the prior distributions of the parameters, we draw samples from the posterior distribution of the parameters using the Metropolis-Hastings algorithm with a Gaussian proposal distribution. We run the algorithm for 200,000 iterations with a burn-in period of 100,000. Finally, the mean of the parameters in each of the iterations are obtained as the final estimates of *β* and *r* for the different time periods. To obtain confidence intervals of various estimates we predict the number of individuals in each compartment given a set of parameters which are drawn using MCMC. This is done for 100,000 iterations. Using these values, we obtain the 95% Bayesian Credible Intervals of the estimates (such as infection fatality rates and underreporting factors)

### Estimation of parameters of interest

Our main parameters of interest here are Underreporting factors for cases and deaths and Infection Fatality rate. Underreporting factors (URF) for cases and deaths are defined as follows:

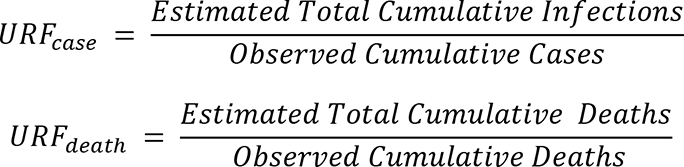

Here, total cumulative cases refers to all Cumulative cases including both reported and unreported cases. Similarly total cumulative deaths includes both reported and unreported deaths. Since we are unable to observe unreported cases or deaths we estimate total cumulative cases and deaths as follows:

1. Total Cumulative cases at time *t* = *P(t)+U(t)+F(t)+RR(t)+RU(t)+DR(t)+DU(t)*
2. Total Cumulative deaths at time *t* = *DR(t)+DU(t)*

To estimate the true fatality rate of COVID-19, we calculate 2 different infection fatality rates IFR1 and IFR2 as defined in formulas (*a*) and (*b*) respectively, in Supplementary Appendix F.

We also calculate the Case fatality rate as defined in the ***Methods*** section.

Now *Cumulative Deaths* follows a *Bin*(*Observerd cumulative cases, CFR_true_*) distribution, with the estimate of *CFR_true_* given by CFR, making CFR is a binomial proportion. Let 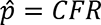 and *n* = *Observerd cumulative cases*. So the asymptotic approximate 95% confidence interval is given by

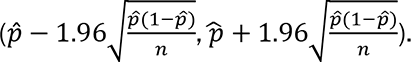

The estimates of infection fatality rates and underreporting factors are based on Bayesian credible intervals constructed from the exact posterior draws, as described before.

## Appendix L. Data source and model-based results

The data has been sourced from *covid19india.org*. We used daily case-recovery-death count data from April 1, 2020 to January 31, 2021 for wave 1 and from February 1, 2021 – June 30, 2021 for wave 2. The predicted number of reported and total cases and deaths for January 31, 2021 (wave 1) and June 30 (for wave 2 and waves 1 and 2 combined) are shown in Tables S1, S2, and S3 respectively.

The mean estimates and the 95% CrI’s of underreporting factors for cases and deaths on January 31, 2021 are shown in Figure S2. Relevant wave 2 values are presented in Figure S3.

**Table T1:**
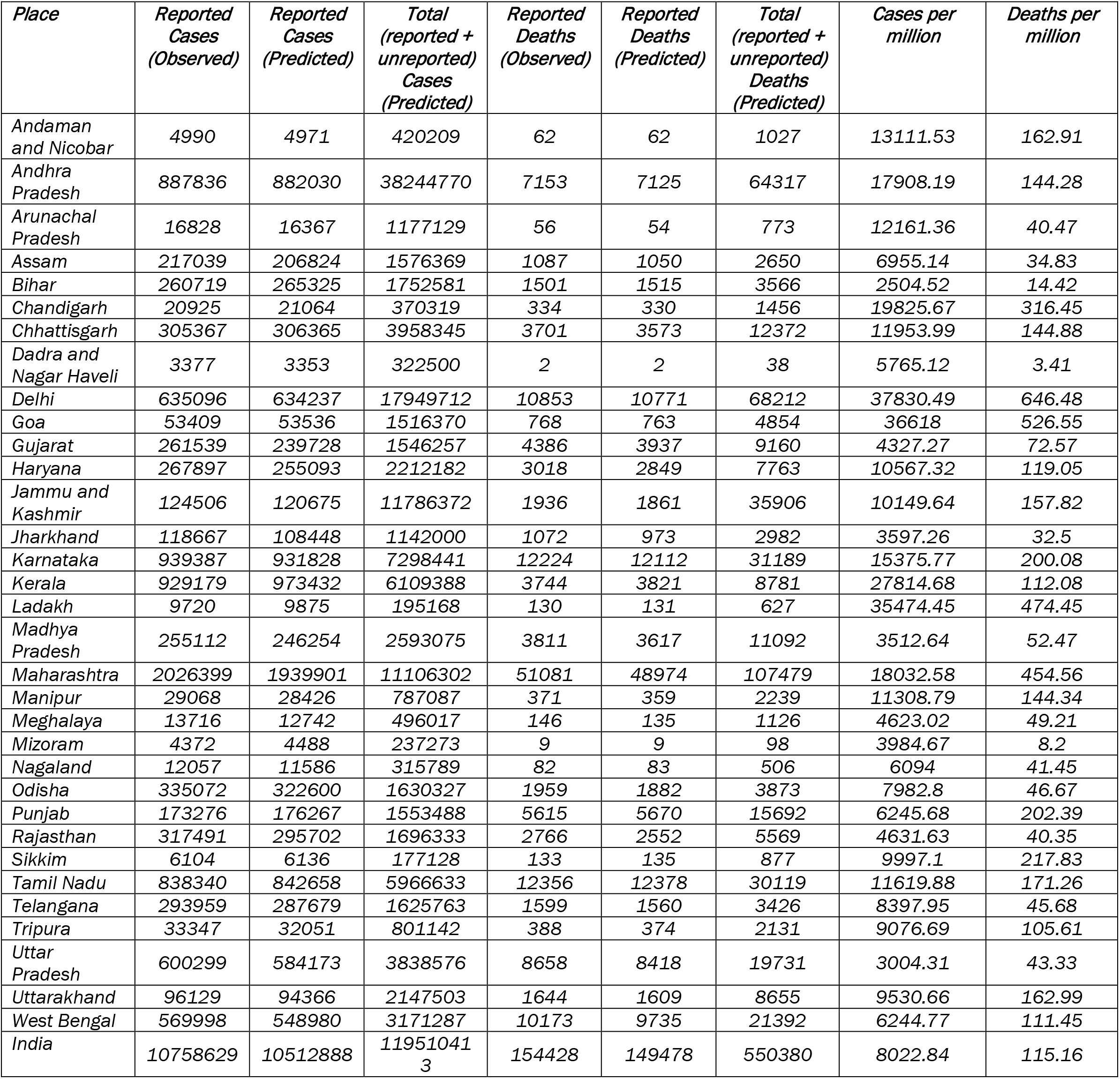
Summary of the different metrics for the states and the nation for wave 1, on 31st January 2021

| **Place** | **Reported**<br>**Cases**<br>**(Observed)** | **Reported**<br>**Cases**<br>**(Predicted)** | **Total**<br>**(reported +**<br>**unreported)**<br>**Cases**<br>**(Predicted)** | **Reported**<br>**Deaths**<br>**(Observed)** | **Reported**<br>**Deaths**<br>**(Predicted)** | **Total**<br>**(reported +**<br>**unreported)**<br>**Deaths**<br>**(Predicted)** | **Cases per**<br>**million** | **Deaths per**<br>**million** |
| --- | --- | --- | --- | --- | --- | --- | --- | --- |
| Andaman and Nicobar | 4990 | 4971 | 420209 | 62 | 62 | 1027 | 13111.53 | 162.91 |
| Andhra Pradesh | 887836 | 882030 | 38244770 | 7153 | 7125 | 64317 | 17908.19 | 144.28 |
| Arunachal Pradesh | 16828 | 16367 | 1177129 | 56 | 54 | 773 | 12161.36 | 40.47 |
| Assam | 217039 | 206824 | 1576369 | 1087 | 1050 | 2650 | 6955.14 | 34.83 |
| Bihar | 260719 | 265325 | 1752581 | 1501 | 1515 | 3566 | 2504.52 | 14.42 |
| Chandigarh | 20925 | 21064 | 370319 | 334 | 330 | 1456 | 19825.67 | 316.45 |
| Chhattisgarh | 305367 | 306365 | 3958345 | 3701 | 3573 | 12372 | 11953.99 | 144.88 |
| Dadra and Nagar Haveli | 3377 | 3353 | 322500 | 2 | 2 | 38 | 5765.12 | 3.41 |
| Delhi | 635096 | 634237 | 17949712 | 10853 | 10771 | 68212 | 37830.49 | 646.48 |
| Goa | 53409 | 53536 | 1516370 | 768 | 763 | 4854 | 36618 | 526.55 |
| Gujarat | 261539 | 239728 | 1546257 | 4386 | 3937 | 9160 | 4327.27 | 72.57 |
| Haryana | 267897 | 255093 | 2212182 | 3018 | 2849 | 7763 | 10567.32 | 119.05 |
| Jammu and Kashmir | 124506 | 120675 | 11786372 | 1936 | 1861 | 35906 | 10149.64 | 157.82 |
| Jharkhand | 118667 | 108448 | 1142000 | 1072 | 973 | 2982 | 3597.26 | 32.5 |
| Karnataka | 939387 | 931828 | 7298441 | 12224 | 12112 | 31189 | 15375.77 | 200.08 |
| Kerala | 929179 | 973432 | 6109388 | 3744 | 3821 | 8781 | 27814.68 | 112.08 |
| Ladakh | 9720 | 9875 | 195168 | 130 | 131 | 627 | 35474.45 | 474.45 |
| Madhya Pradesh | 255112 | 246254 | 2593075 | 3811 | 3617 | 11092 | 3512.64 | 52.47 |
| Maharashtra | 2026399 | 1939901 | 11106302 | 51081 | 48974 | 107479 | 18032.58 | 454.56 |
| Manipur | 29068 | 28426 | 787087 | 371 | 359 | 2239 | 11308.79 | 144.34 |
| Meghalaya | 13716 | 12742 | 496017 | 146 | 135 | 1126 | 4623.02 | 49.21 |
| Mizoram | 4372 | 4488 | 237273 | 9 | 9 | 98 | 3984.67 | 8.2 |
| Nagaland | 12057 | 11586 | 315789 | 82 | 83 | 506 | 6094 | 41.45 |
| Odisha | 335072 | 322600 | 1630327 | 1959 | 1882 | 3873 | 7982.8 | 46.67 |
| Punjab | 173276 | 176267 | 1553488 | 5615 | 5670 | 15692 | 6245.68 | 202.39 |
| Rajasthan | 317491 | 295702 | 1696333 | 2766 | 2552 | 5569 | 4631.63 | 40.35 |
| Sikkim | 6104 | 6136 | 177128 | 133 | 135 | 877 | 9997.1 | 217.83 |
| Tamil Nadu | 838340 | 842658 | 5966633 | 12356 | 12378 | 30119 | 11619.88 | 171.26 |
| Telangana | 293959 | 287679 | 1625763 | 1599 | 1560 | 3426 | 8397.95 | 45.68 |
| Tripura | 33347 | 32051 | 801142 | 388 | 374 | 2131 | 9076.69 | 105.61 |
| Uttar Pradesh | 600299 | 584173 | 3838576 | 8658 | 8418 | 19731 | 3004.31 | 43.33 |
| Uttarakhand | 96129 | 94366 | 2147503 | 1644 | 1609 | 8655 | 9530.66 | 162.99 |
| West Bengal | 569998 | 548980 | 3171287 | 10173 | 9735 | 21392 | 6244.77 | 111.45 |
| India | 10758629 | 10512888 | 119510413 | 154428 | 149478 | 550380 | 8022.84 | 115.16 |

**Table T2:** Summary of the different metrics for the states and the nation for wave 2, on 30th June 2021

| *Place* | *Reported Cases*<br>*(Observed)* | *Reported Cases*<br>*(Predicted)* | *Total*<br>*(reported +*<br>*unreported)*<br>*Cases*<br>*(Predicted)* | *Reported Deaths*<br>*(Observed)* | *Reported Deaths*<br>*(Predicted)* | *Total*<br>*(reported +*<br>*unreported)*<br>*Deaths*<br>*(Predicted)* | *Cases per*<br>*million* | *Deaths per*<br>*million* |
| --- | --- | --- | --- | --- | --- | --- | --- | --- |
| Andaman and Nicobar | 2475 | 2538 | 22006 | 66 | 63 | 178 | 6503.21 | 173.42 |
| Andhra Pradesh | 1005454 | 1052130 | 24433883 | 5590 | 5404 | 30424 | 20280.61 | 112.75 |
| Arunachal Pradesh | 19341 | 21105 | 168634 | 120 | 115 | 308 | 13977.47 | 86.72 |
| Assam | 293999 | 320108 | 10416260 | 3494 | 3439 | 25964 | 9421.36 | 111.97 |
| Bihar | 461307 | 459699 | 9984295 | 8089 | 7695 | 40041 | 4431.41 | 77.7 |
| Chandigarh | 40730 | 41573 | 581721 | 474 | 468 | 1746 | 38590.17 | 449.1 |
| Chhattisgarh | 689201 | 703661 | 12440190 | 9739 | 9717 | 42812 | 26979.67 | 381.25 |
| Dadra and Nagar Haveli | 7047 | 7415 | 132289 | 2 | 2 | 9 | 12030.44 | 3.41 |
| Delhi | 799064 | 806934 | 14348458 | 14125 | 14016 | 62005 | 47597.5 | 841.38 |
| Goa | 113451 | 116695 | 1343287 | 2292 | 2244 | 7401 | 77783.68 | 1571.43 |
| Gujarat | 561769 | 593952 | 9133281 | 5674 | 5834 | 23185 | 9294.7 | 93.88 |
| Haryana | 500735 | 522462 | 10517838 | 6424 | 6479 | 31671 | 19751.72 | 253.4 |
| Jammu and Kashmir | 191410 | 197500 | 7943113 | 2391 | 2315 | 20502 | 15603.61 | 194.91 |
| Jharkhand | 226971 | 236331 | 4274772 | 4040 | 4079 | 18394 | 6880.38 | 122.47 |
| Karnataka | 1907238 | 1962929 | 37788085 | 22914 | 22533 | 107790 | 31217.43 | 375.05 |
| Kerala | 2004396 | 2083298 | 23997385 | 9599 | 9266 | 30606 | 60000.97 | 287.34 |
| Ladakh | 10366 | 10916 | 117421 | 72 | 72 | 224 | 37832.12 | 262.77 |
| Madhya Pradesh | 534581 | 549186 | 8512735 | 5169 | 5160 | 20615 | 7360.66 | 71.17 |
| Maharashtra | 4042252 | 4073053 | 59223522 | 71088 | 70007 | 268306 | 35971.31 | 632.6 |
| Manipur | 41220 | 46295 | 481010 | 791 | 772 | 2480 | 16036.48 | 307.74 |
| Meghalaya | 36326 | 38828 | 1512205 | 698 | 674 | 6122 | 12243.8 | 235.26 |
| Mizoram | 16120 | 17044 | 306787 | 84 | 86 | 427 | 14691.86 | 76.56 |
| Nagaland | 13206 | 13915 | 383920 | 410 | 423 | 2782 | 6674.75 | 207.23 |
| Odisha | 577736 | 594332 | 20182943 | 2157 | 1989 | 15246 | 13764.07 | 51.39 |
| Punjab | 422429 | 421946 | 10421537 | 10456 | 9990 | 57838 | 15226.32 | 376.88 |
| Rajasthan | 634910 | 633034 | 16637039 | 6157 | 5919 | 35662 | 9262.21 | 89.82 |
| Sikkim | 14575 | 15696 | 218046 | 175 | 182 | 703 | 23870.86 | 286.61 |
| Tamil Nadu | 1645335 | 1726816 | 40903538 | 20358 | 19214 | 109652 | 22805.3 | 282.17 |
| Telangana | 329792 | 333956 | 8438699 | 2068 | 2005 | 11775 | 9421.64 | 59.08 |
| Tripura | 32869 | 35134 | 1214075 | 291 | 271 | 2236 | 8946.58 | 79.21 |
| Uttar Pradesh | 1105782 | 1132551 | 25351545 | 13939 | 13930 | 69963 | 5534.1 | 69.76 |
| Uttarakhand | 244199 | 253662 | 4885358 | 5676 | 5771 | 27475 | 24210.98 | 562.74 |
| West Bengal | 931107 | 958952 | 24747185 | 7556 | 7329 | 43904 | 10200.99 | 82.78 |
| India | 19690200 | 19890297 | 2.61E+08 | 245824 | 236813 | 850612 | 14683.22 | 183.31 |

**Table T3:** Summary of the different metrics for the states and the nation for waves 1 and 2 combined, on 30th June 2021

| *Place* | *Reported Cases (Observed)* | *Reported Cases (Predicted)* | *Total (reported + unreported) Cases (Predicted)* | *Reported Deaths (Observed)* | *Reported Deaths (Predicted)* | *Total (reported + unreported) Deaths (Predicted)* | *Cases per million* | *Deaths per million* |
| --- | --- | --- | --- | --- | --- | --- | --- | --- |
| Andaman and Nicobar | 6589 | 7509 | 442215 | 88 | 125 | 1205 | 19614.74 | 336.33 |
| Andhra Pradesh | 1435447 | 1934160 | 62678653 | 9372 | 12529 | 94741 | 38188.8 | 257.03 |
| Arunachal Pradesh | 21803 | 37472 | 1345763 | 81 | 169 | 1081 | 26138.83 | 127.19 |
| Assam | 328526 | 526932 | 11992629 | 2184 | 4489 | 28614 | 16376.5 | 146.8 |
| Bihar | 651888 | 725024 | 11736876 | 3831 | 9210 | 43607 | 6935.93 | 92.12 |
| Chandigarh | 55352 | 62637 | 952040 | 635 | 798 | 3202 | 58415.84 | 765.55 |
| Chhattisgarh | 912468 | 1010026 | 16398535 | 11734 | 13290 | 55184 | 38933.66 | 526.13 |
| Dadra and Nagar Haveli | 9515 | 10768 | 454789 | 4 | 4 | 47 | 17795.56 | 6.82 |
| Delhi | 1393747 | 1441171 | 32298170 | 21504 | 24787 | 130217 | 85427.99 | 1487.86 |
| Goa | 135851 | 170231 | 2859657 | 2099 | 3007 | 12255 | 114401.7 | 2097.98 |
| Gujarat | 752544 | 833680 | 10679538 | 9114 | 9771 | 32345 | 13621.97 | 166.45 |
| Haryana | 694384 | 777555 | 12730020 | 6685 | 9328 | 39434 | 30319.04 | 372.45 |
| Jammu and Kashmir | 244553 | 318175 | 19729485 | 3147 | 4176 | 56408 | 25753.25 | 352.73 |
| Jharkhand | 315475 | 344779 | 5416772 | 4479 | 5052 | 21376 | 10477.64 | 154.97 |
| Karnataka | 2203361 | 2894757 | 45086526 | 21841 | 34645 | 138979 | 46593.2 | 575.13 |
| Kerala | 2147727 | 3056730 | 30106773 | 6427 | 13087 | 39387 | 87815.65 | 399.42 |
| Ladakh | 16439 | 20791 | 312589 | 165 | 203 | 851 | 73306.57 | 737.22 |
| Madhya Pradesh | 731319 | 795440 | 11105810 | 6988 | 8777 | 31707 | 10873.3 | 123.64 |
| Maharashtra | 5378150 | 6012954 | 70329824 | 81475 | 118981 | 375785 | 54003.89 | 1087.16 |
| Manipur | 39722 | 74721 | 1268097 | 578 | 1131 | 4719 | 27345.27 | 452.08 |
| Meghalaya | 23284 | 51570 | 2008222 | 320 | 809 | 7248 | 16866.82 | 284.47 |
| Mizoram | 8678 | 21532 | 544060 | 24 | 95 | 525 | 18676.53 | 84.76 |
| Nagaland | 18039 | 25501 | 699709 | 203 | 506 | 3288 | 12768.75 | 248.68 |
| Odisha | 612220 | 916932 | 21813270 | 2366 | 3871 | 19119 | 21746.87 | 98.06 |
| Punjab | 497663 | 598213 | 11975025 | 11891 | 15660 | 73530 | 21472 | 579.27 |
| Rajasthan | 859576 | 928736 | 18333372 | 6777 | 8471 | 41231 | 13893.84 | 130.17 |
| Sikkim | 11424 | 21832 | 395174 | 205 | 317 | 1580 | 33867.96 | 504.44 |
| Tamil Nadu | 1598092 | 2569474 | 46870171 | 17669 | 31592 | 139771 | 34425.18 | 453.43 |
| Telangana | 528216 | 621635 | 10064462 | 2949 | 3565 | 15201 | 17819.59 | 104.76 |
| Tripura | 40813 | 67185 | 2015217 | 428 | 645 | 4367 | 18023.27 | 184.82 |
| Uttar Pradesh | 1619541 | 1716724 | 29190121 | 17546 | 22348 | 89694 | 8538.41 | 113.09 |
| Uttarakhand | 287279 | 348028 | 7032861 | 4811 | 7380 | 36130 | 33741.64 | 725.73 |
| West Bengal | 1133393 | 1507932 | 27918472 | 13281 | 17064 | 65296 | 16445.76 | 194.23 |
| India | 24963227 | 30403185 | 3.81E+08 | 273779 | 386291 | 1400992 | 22706.06 | 298.47 |

**Table T4:** Parameter values and descriptions for the SEIR-fansy model.

| Parameter | Value | Description |
| --- | --- | --- |
|  | *Time-varying* | Rate of infectious transmission by infected, tested individuals with false negative results. |
| p | 0.5 | Ratio of rate of spread of infection by tested positive patients to that by false negatives. $p < 1$ represents the scenario where individuals who test positive are infecting susceptible individuals at a lower rate than infected individuals with false negative test results. |
| u | 0.5 | Scaling factor for the rate of spread of infection by untested individuals. u is assumed to be $< 1$ as U mostly consists of asymptomatic or mildly symptomatic cases who are known to spread the disease at a much lower rate than those with higher levels of symptoms. |
| De | 5.2 | Incubation period (in days). |
| Dr | 17.8 | Means number of days until recovery for infected individuals. |
| Dt | 0 | Mean number of days for the test result to come after a person is tested. Under the assumption of instantaneous test results, this is taken to be zero. |
| c | 0.0562 | Death rate attributable to COVID-19 which is equivalent to inverse of the average number of days for death starting from the onset of disease times the probability of death of an infected individual. |
| $\lambda, \mu$ | $3.95 \times 10^{-5}$ | Natural birth and death rates (assumed to be equal). |
| r | *Time-varying* | Probability of being tested for infectious individuals. |
| f | 0.15 | Probability of a false negative RT-PCR diagnostic test result. |
| 1, 12 | 0.6 (1)<br>0.7 (2) | Scaling factors for rate of recovery for undetected and false negative individuals respectively. Both 1 and 2 are assumed to be less than 1. It is assumed that the recovery rate is slower than the detected ones for the False Negative ones because they are not getting any hospital treatments. The condition of Untested individuals is not so severe as they consist of mostly asymptomatic people. So, they are assumed to recover faster than the Current Positive Ones. |
| 1, 12 | 0.3 (1)<br>0.7 (2) | Scaling factors for death rate for undetected and false negative individuals respectively. Both 1 and 2 are assumed to be less than 1. Same as before, the death rate for False Negative ones are assumed to be higher than the Current detected Positive as they are not receiving proper treatment. While, for the Untested ones, the death rate is taken to be lesser because they are mostly asymptomatic. So, their probability of dying is much less. |

**Table T5:** Comparison of 4th nationwide serosurvey to combined estimates across waves 1 and 2.

|  | 4th Nationwide Serosurvey, 31 May 2021<sup>1</sup> |  |  |  | Model-Based Estimates, 30 June 2021 |  |  |  |
| --- | --- | --- | --- | --- | --- | --- | --- | --- |
| States | Seroprevalence (%) | Estimated Cases (in Lakhs) | Reported Cases (in Lakhs) | Under Counting Factor | Estimated Cases | Reported Cases | Under Reporting Factor | Difference (Under Counting factor - Under Reporting factor) |
| Bihar | 75.9 | 947 | 7 | 134 | 11733035 | 725024 | 16 | 118 |
| Uttar Pradesh | 71 | 1689 | 17 | 100 | 29700720 | 1716724 | 17 | 83 |
| Madhya Pradesh | 79 | 674 | 8 | 86 | 11248345 | 795440 | 14 | 72 |
| Jharkhand | 61.2 | 236 | 3 | 70 | 5494715 | 344779 | 16 | 54 |
| Rajasthan | 76.2 | 617 | 9 | 66 | 18167796 | 928736 | 20 | 46 |
| Gujarat | 75.3 | 481 | 8 | 59 | 11073822 | 833680 | 13 | 46 |
| West Bengal | 60.9 | 607 | 14 | 44 | 28541606 | 1507932 | 19 | 25 |
| Assam | 50.3 | 179 | 4 | 44 | 12843468 | 526932 | 24 | 20 |
| Telangana | 63.1 | 243 | 6 | 42 | 10136279 | 621635 | 16 | 26 |
| Odisha | 68.1 | 316 | 8 | 41 | 22332360 | 916932 | 24 | 17 |
| Punjab | 66.5 | 200 | 6 | 35 | 11989926 | 598213 | 20 | 15 |
| Jammu & Kashmir | 63 | 86 | 3 | 30 | 19619546 | 318175 | 62 | -32 |
| Tamil Nadu | 69.2 | 539 | 21 | 26 | 48926546 | 2569474 | 19 | 7 |
| Uttarakhand | 73.1 | 82 | 3 | 25 | 7182789 | 348028 | 21 | 4 |
| Himachal Pradesh | 62 | 46 | 2 | 24 | N/A | N/A | N/A | N/A |
| Chhattisgarh | 74.6 | 220 | 10 | 23 | 459405 | 10768 | 43 | -20 |
| Haryana | 60.1 | 169 | 8 | 22 | 13080661 | 777555 | 17 | 5 |
| Andhra Pradesh | 70.2 | 378 | 17 | 22 | 63562840 | 1934160 | 33 | -11 |
| Karnataka | 69.8 | 472 | 26 | 18 | 46131203 | 2894757 | 16 | 2 |
| Maharashtra | 58 | 714 | 57 | 12 | 70307014 | 6012954 | 12 | 0 |
| Kerala | 44.4 | 159 | 25 | 6 | 31342383 | 3056730 | 10 | -4 |
| India | 67.6 | 9265 | 282 | 33 | 380801295 | 30403185 | 13 | 20 |

N/A = Not available.
[1] Press Information Bureau, National Media Center. Briefing on COVID-19. July 20, 2021.

**Supplementary Figure 2.**
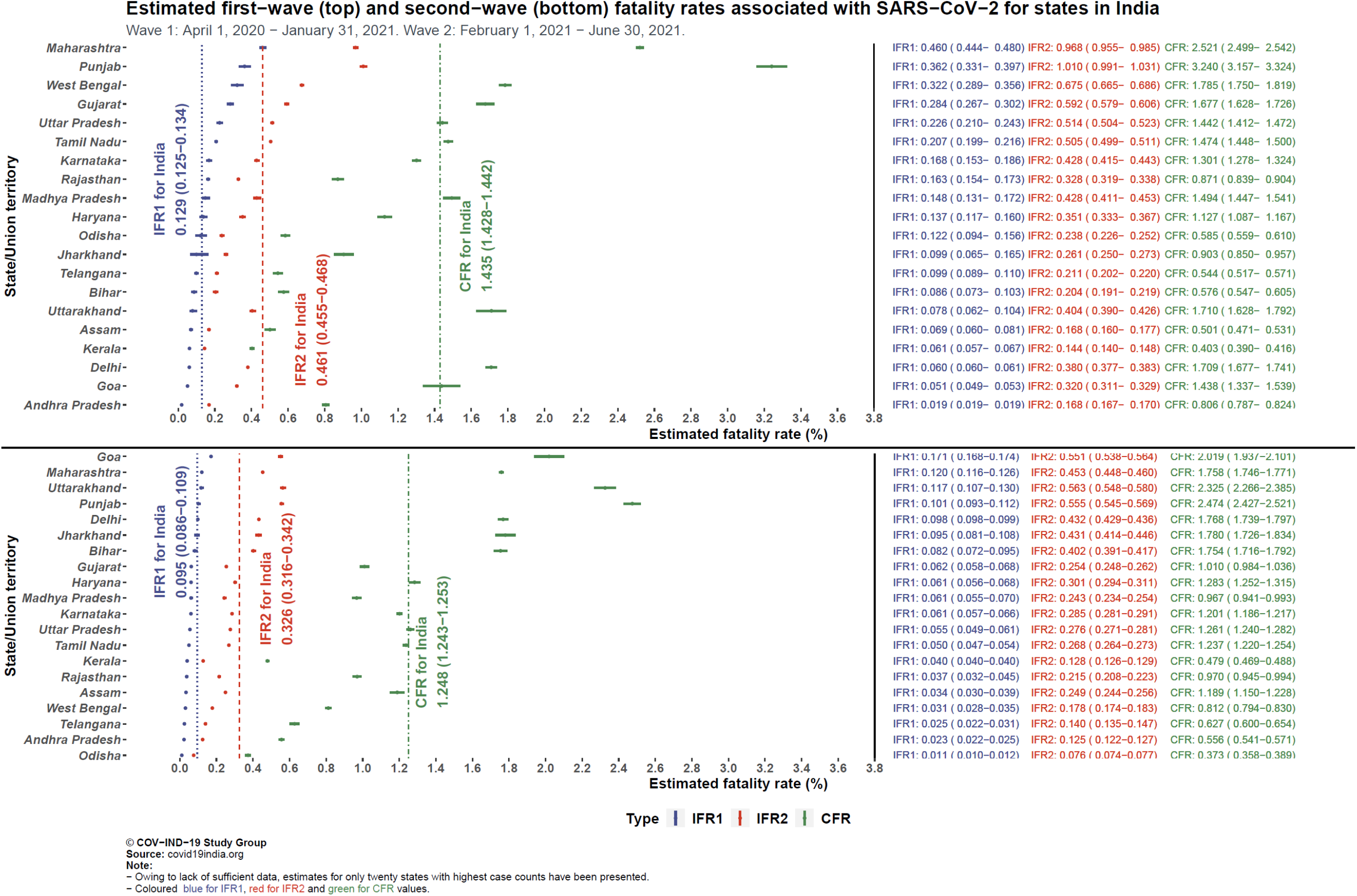
Forest plot of wave 1 and wave 2 infection fatality rates (IFR) and case fatality ratios (CFR) for SARS-CoV-2 in various states in India, where IFR_1_ includes reported deaths and IFR_2_ further includes the estimate of unreported deaths.

**Supplementary Figure 3:**
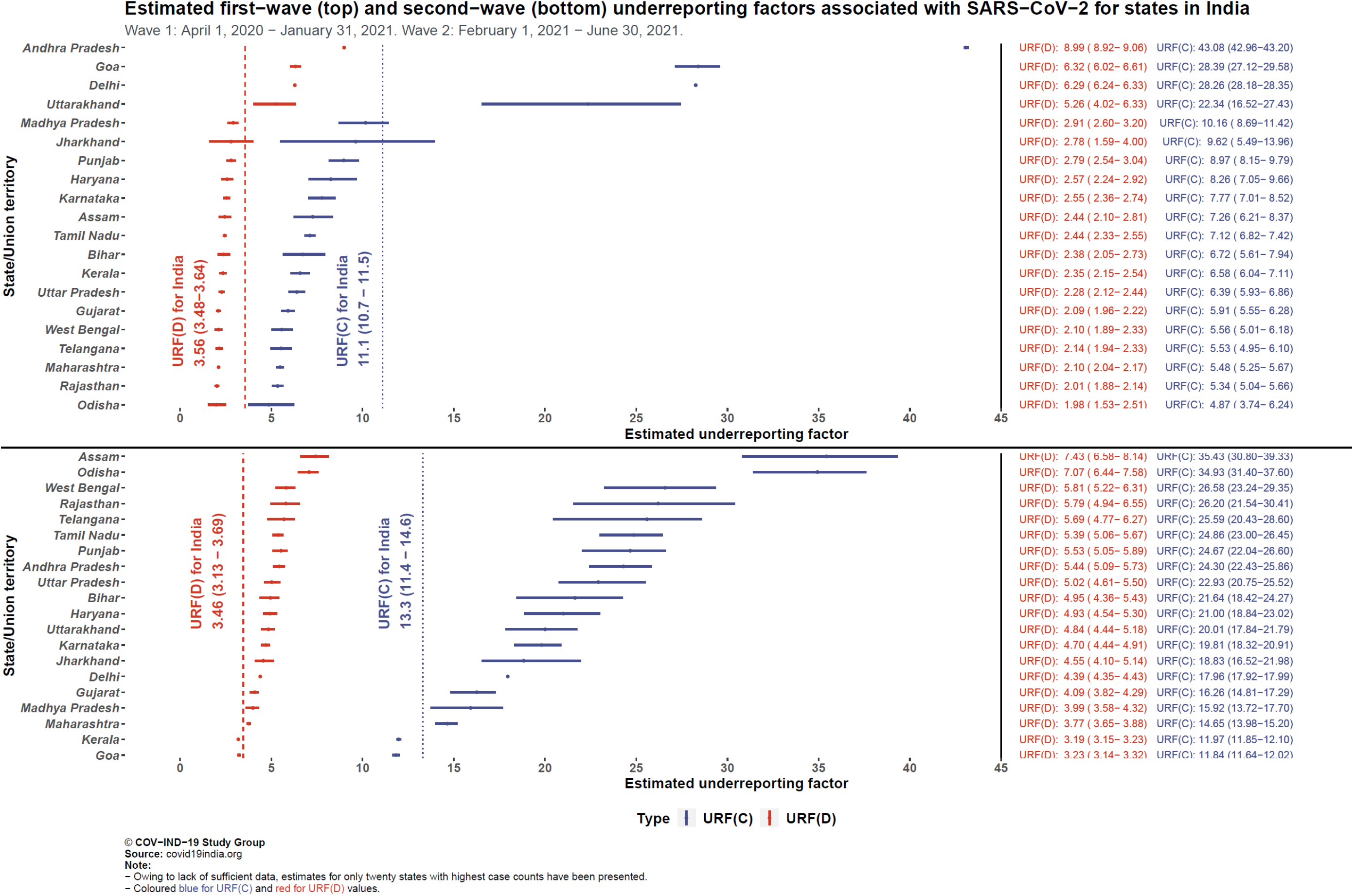
Estimated first and second wave underreporting factors for cases and deaths associated with SARS-CoV-2 for states in India.

## Footnotes

1 The paucity of data in poorer countries has prompted some economists to adopt the benefit transfer method (BMT) – VSL estimates from richer countries are taken off-the-shelf and linearly scaled down to produce an estimate for poorer countries using the per capita GDP ratio. As Majumder and Madheswaran (2018) (*85*) note, this is likely to generate a significant underestimate since available data suggest the income elasticity of the demand for safety is considerably less than one, i.e., the poor are willing to pay a much higher proportion of their incomes to increase the chance of staying alive, in similar spirit to Engel’s Law for food.

2 Other estimates of VSL in India are as follows: INR 14 - 19 million (*86*), INR 6.4 - 15 million (*87*), INR 14.8 – 15.4 million (*88*), INR 10.28 million (*89*).

